# Comparing DOACs with warfarin in AF patients with chronic kidney disease or valvular disease: A systematic review and meta-analysis

**DOI:** 10.1101/2024.01.13.24301121

**Authors:** Aileen Liang, Cathy Wang, Alla E Iansavichene, Alejandro Lazo-Langner

## Abstract

**Objective:** To analyze the safety and efficacy of different direct oral anticoagulant agents (DOACs) compared to warfarin in patients with concomitant atrial fibrillation (AF) and valvular disease or concomitant AF and chronic kidney disease (CKD).

**Methods:** We conducted literature searches in MEDLINE, Embase, and EBM Reviews to examine randomized-controlled trials (RCTs) and non-RCTs that included the aforementioned patient populations treated with warfarin or DOAC (rivaroxaban, dabigatran, apixaban, or edoxaban) and assessed outcomes of bleeding, stroke, or systemic/arterial thromboembolism. Meta-analysis was performed for eligible studies using the Mantel-Haenszel method random-effects model.

**Results:** 3,172 studies were screened and 154 studies were selected after two levels of screening. Meta-analysis showed that, in patients with concomitant AF and CKD, DOAC was associated with reduced bleeding in non-RCTs (OR 0.65, 95% Cl [0.49, 0.86], p=0.003), particularly in more severe CKD (eGFR < 60mL/min/1.73m^2^). Apixaban in particular was associated with reduced bleeding (OR 0.52, 95% Cl [0.44, 0.63], p<0.00001) and stroke incidence (OR 0.60, 95% Cl [0.41, 0.87], p=0.007). In patients with concomitant AF and valvular disease, DOAC was associated with reduced bleeding (OR 0.75, 95% CI [0.57, 0.97], p=0.03) and stroke incidence (OR 0.66, 95% CI [0.47, 0.93], p=0.02) in non-RCTs.

**Conclusion:** Our study studied populations that are typically excluded from large-scale anticoagulation studies and our findings suggest that DOACs may be superior to warfarin both in the prevention of thromboembolic event and in the reduction of bleeding risks in patients with concomitant CKD or valvular disease.

## 1. Introduction

In recent years, the use of direct oral anticoagulants (DOACs) such as apixaban, dabigatran, edoxaban, and rivaroxaban have become first-line choice for most anticoagulant indications. DOACs have been shown to be superior to the previous standard of care drug warfarin in treatment of non-valvular atrial fibrillation (AF) in several large-scale trials and is preferred for reasons including having fewer drug and food interactions and eliminating the need for coagulation test monitoring.^1–3^ Namely, RE-LY, ROCKET-AF, ARISTOTLE, and ENGAGE AF-TIMI 48 are four large-scale trials that have been paramount in solidifying the role of DOACs in treatment of atrial fibrillation and crucial to the establishment of many current guidelines^1,2,4,5^.

The RE-LY trial enrolled 18,113 participants and compared dabigatran with warfarin to show that dabigatran was noninferior to warfarin in terms of primary efficacy outcomes of stroke and systemic embolism and was also superior to warfarin in primary safety outcome of bleeding depends on the dosage. The ROCKET-AF trial enrolled 14,264 participants in comparing rivaroxaban to warfarin, demonstrating that rivaroxaban was noninferior to warfarin for the prevention of stroke or systemic embolism, and in addition led to fewer occurrences of intracranial and fatal bleeding. ARISTOTLE is another trial with 18,201 participants that compared apixaban to warfarin to show apixaban was superior to warfarin in preventing stroke or systemic embolism, caused less bleeding, and resulted in lower mortality. Lastly, ENGAGE AF-TIMI 48 had 21,105 participants in their comparison of edoxaban to warfarin, and showed that edoxaban was noninferior to warfarin with respect to the prevention of stroke or systemic embolism and were associated with significantly lower rates of bleeding and death from cardiovascular causes.

All these large-scale studies have excluded patients with concomitant moderate to severe mitral stenosis, patients with bioprosthetic heart valves, as well as patients with severe renal insufficiency (clearance <25 mL or <30 mL). As a result, although DOACs have been established for use in non-valvular AF in patients without comorbidities, it is less clear how efficient and safe DOACs are in these select populations.

Patients with concomitant AF and CKD or concomitant AF and valvular disease are high-risk populations that require extensive consideration for anticoagulation therapies. The present study aimed to review the outcomes of available literature to analyze the safety and efficacy of DOACs compared to warfarin in patients with concomitant AF and valvular disease or concomitant AF and CKD. To further characterize the comparative benefits and risks of DOACs, our review included subgroup analyses of specific DOACs as well as patients with different stages of CKD when applicable.

## 2. Methods

This study was a systematic review and meta-analysis and was developed in accordance with the Preferred Reporting Items for Systematic Reviews and Meta-Analysis guidelines. The protocol of this review is documented on the International prospective register of systematic reviews (PROSPERO) registry under ID CRD42022322633^6^.

### 2.1 Eligibility criteria

Eligible studies included adult patients (age > 18) with concomitant atrial fibrillation and valvular disease (stenosis, prolapse, or regurgitation of the mitral, tricuspid, pulmonary, or aortic valve) or adult patients with concomitant atrial fibrillation and chronic kidney disease who are treated with warfarin or one of the following DOACs: rivaroxaban, dabigatran, apixaban, or edoxaban. Eligible studies investigated the safety and/or efficacy of the anticoagulant by assessing at least one of the following primary outcomes: incidence of bleeding, incidence of stroke, or incidence of systemic/arterial thromboembolism. If available, secondary outcome of all-cause mortality was also included. In addition, study design was limited to randomized-controlled trials, cohort studies, or case-control studies published in peer-reviewed journals. Conference proceedings and case studies were not included. Studies were also limited to those published in English to mitigate risk of mistranslation. Patients with valvular disease who have mechanical valves were also excluded.

### 2.2 Information Sources

A research librarian was consulted in the development of the search strategies. Relevant documents were identified through literature searches conducted in databases including MEDLINE, Embase, and EBM Reviews - Cochrane Central Register of Controlled Trials (all via Ovid interface). Keywords and medical subject headings (MeSH) were used to identify all relevant studies and no date restrictions were placed on the searches. Manual citation screening of included trials was also conducted to ensure a comprehensive search. Complete search strategy is available in the **Supplementary materials**.

### 2.3 Study selection and screening

All literature search results were uploaded to Covidence (Covidence systematic review software, Veritas Health Innovation, Melbourne, Australia. Available at www.covidence.org) and duplicates were removed through an automated duplication check conducted by Covidence and a manual duplicate check conducted by reviewers (AL and CW).

Two reviewers (AL and CW) independently conducted a title abstract screening of the eligible retrieved articles using Covidence. Potentially relevant studies then underwent full text screening by both reviewers independently. After each level of screening, conflicts were resolved during a consensus meeting between the two reviewers. Cohen’s kappa (K) coefficient was calculated for each level of screening to assess for inter-rater reliability in screening.

### 2.4 Data items

Data was extracted from the included studies independently by two reviewers using a standard form. Any discrepancies were resolved during a consensus meeting between the two reviewers. Extracted data pertained to study characteristics (authors, publication date, study design, location, number of participants), participant demographics (age, sex, concurrent antiplatelet use, comorbid conditions including heart failure, coronary artery disease, diabetes, hypertension, previous stroke, and previous venous thromboembolism), stage of CKD, intervention (medication and dosage), and measured outcomes (major bleeds, GI bleeds, intracranial bleeds, fatal bleeds, overall strokes, TIA, systemic/arterial thromboembolism, and all-cause mortality). Major bleeding events were defined individually by each trial but, in most cases, the definitions and criteria from the International Society on Thrombosis and Haemostasis were used^7^. For patients with CKD, stages were defined by individual studies, but most used estimated glomerular filtration rate (eGFR) based on criteria from National Kidney Foundation’s KDOQI clinical practice guidelines: stage 1 eGFR >90, stage 2 eGFR 60-89, stage 3 eGFR 30-59, stage 4 eGFR 15-29, and stage 5 eGFR <15^8^.

### 2.5 Risk of bias assessment

For randomized studies, the second version of the Cochrane risk-of-bias tool for randomized trials (RoB 2) and the Jadad Scale were used to assess the risk of bias in all the studies analyzed in this paper^9,10^. RoB2 will identify bias in the following domains: 1) risk of bias arising from the randomization process, 2) bias due to deviations from intended interventions, 3) bias due to missing outcome data, 4) bias in measurement of the outcome, and 5) bias in selection of the reported result. The Jadad Scale assessed randomization, blinding, and accounts of all patients. For non-randomized studies, the risk of bias in non-randomized studies of interventions (ROBINS-I) tool and the Newcastle-Ottawa Scale were used to assess the risk of bias^11,12^. ROBINS-I identified the following domains: 1) bias due to confounding, 2) bias in the selection of participants into the study, 3) bias in classification of interventions, 4) bias due to deviations from intended interventions, 5) bias due to missing data, 6) bias in measurement of outcomes, and 7) bias in the selection of the reported result. The Newcastle-Ottawa Scale assessed selection, comparability, and outcome/exposure. Complete risk-of-bias information can be found in the **Supplementary materials (supplementary figures 1 and 2 and supplementary tables 1 and 2)**.

### 2.6 Data synthesis methods

Meta-analysis was conducted on eligible studies. Data extraction of the number of patients and incidence of events were used. Pooled odds ratios with 95% confidence intervals were obtained. A p-value less than 0.05 was considered statistically significant. A random effects model was used as statistical heterogeneity was high (I^2^>50%) based on Higgins *I*^2^-statistics. The results of the meta-analysis were reported using forest plots and publication bias was assessed using funnel plots. Meta-analysis was conducted using Review Manager (RevMan, Version 5; The Nordic Cochrane Center, Copenhagen).

### 2.7 Subgroup and sensitivity analyses

When data allowed, we compared primary and secondary outcomes based on subgroups of 1) RCT vs non-RCTs, 2) specific DOAC, 3) stages of CKD, 4) types of bleeding (GI, intracranial, fatal), and 5) overall stroke vs TIA.

## 3. Results

### 3.1 Study selection and characteristics

The PRISMA flow diagram of screening process is shown in **Figure 1**. The search strategy yielded 3,172 results that were published between 1946 and 2022. After title and abstract screening, 284 studies remained and underwent full-text screening. Through full-text screening, 110 studies met the inclusion criteria. The PRISMA flow diagram of screening process is shown below. The characteristics of the included studies are shown in the **Supplementary material (Supplementary tables 3-6)**. For RCTs examining CKD patients, the range for participants’ mean age was 64.2-81 years of age, and the percentage of male participants was between 38.2% - 86.3%. For non-RCTs of CKD patients, the mean age ranged from 59.8 - 84.6 years and the percentage of male participants ranged from 36.7%-95.9%. For both non-RCT and RCT analyses, there are patients included with all five stages of CKD. For RCTs assessing valve disease patients, the mean age ranged from 45.7-92 years and the % male ranged from 20.2-60.9%. Valve disease profiles included mitral stenosis, mitral regurgitation, aortic regurgitation, aortic stenosis, and tricuspid regurgitation with or without valve repair and replacements. For non-RCTs examining valve disease patients, the participants’ mean age ranged from 59.2-84 years and the % male participants ranged from 5% to 66.7%. Valve disease profiles included mitral stenosis, mitral regurgitation, aortic regurgitation, aortic stenosis, tricuspid regurgitation, and pulmonary valve disease with or without valve repair and replacements. The funnel plots for each one of the main analyses can be found in the **Supplementary material**.

**Figure 1.**
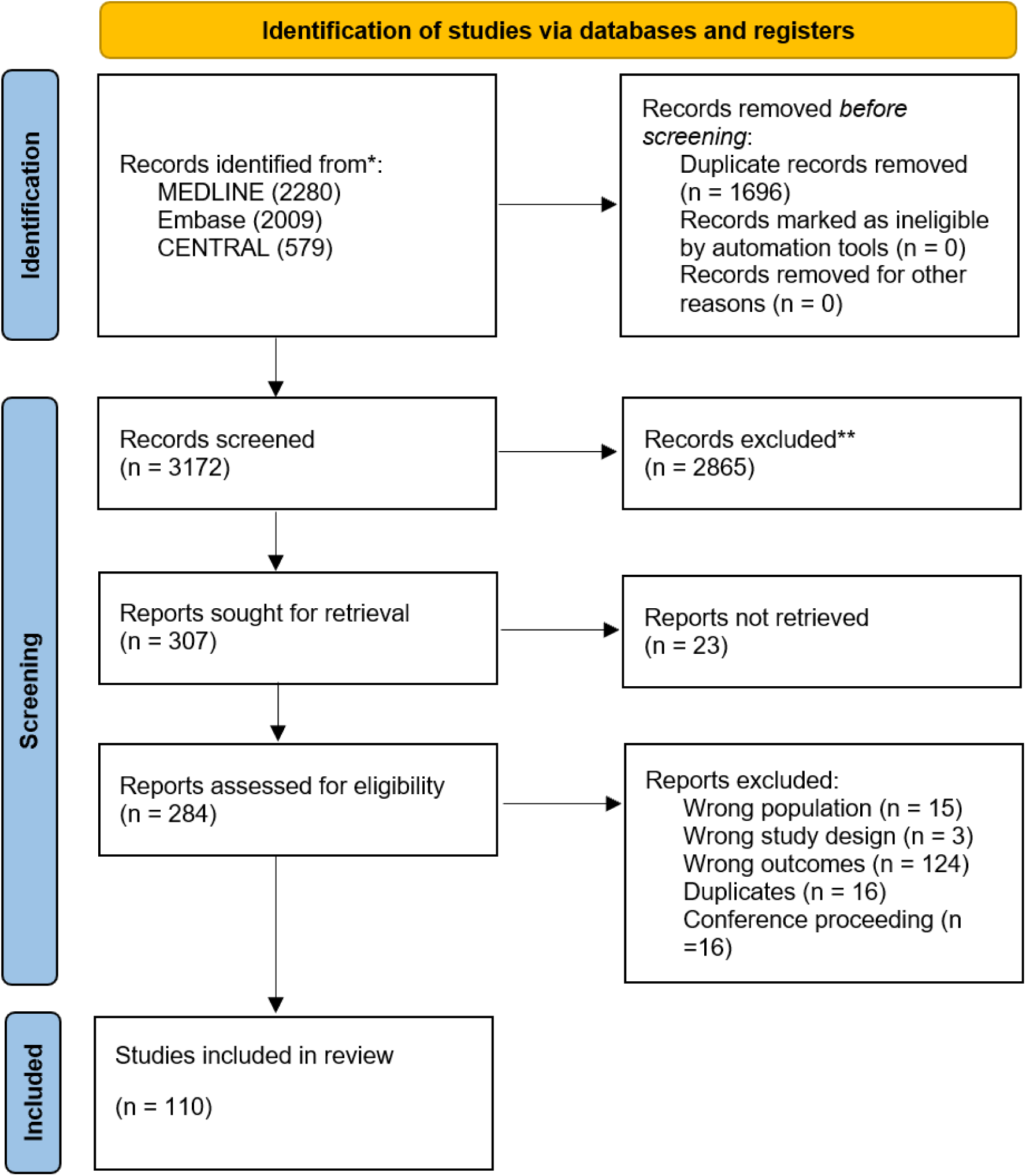
PRISMA flow diagram of screening process

### 3.2 Patients with concomitant AF and CKD

Our analysis of patients with concomitant AF and CKD assessed 15 RCTs and 65 non-RCTs including 310,478 patients with 55,259 total participants in the RCTs and 255,219 participants in the non-RCTs. For studies that include both patients with and without CKD, only participants with CKD were included in the analysis in this study. If eGFR was specified in study, the patients were included in both overall and sensitivity analyses with different eGFR stages.

#### 3.2.1 Bleeding events

All 15 RCTs and 65 non-RCTs included in our review evaluated overall bleeding as an outcome. Meta-analysis of RCTs showed no significant difference in the number of overall bleeding events between warfarin and DOACs (OR 0.84, 95% Cl [0.68, 1.02], p=0.08). Meta-analysis of the non-RCTs favored DOACs for fewer bleeding events than warfarin (OR 0.65, 95% Cl [0.49, 0.86], p=0.003). (**Figure 2**)

**Figure 2.**
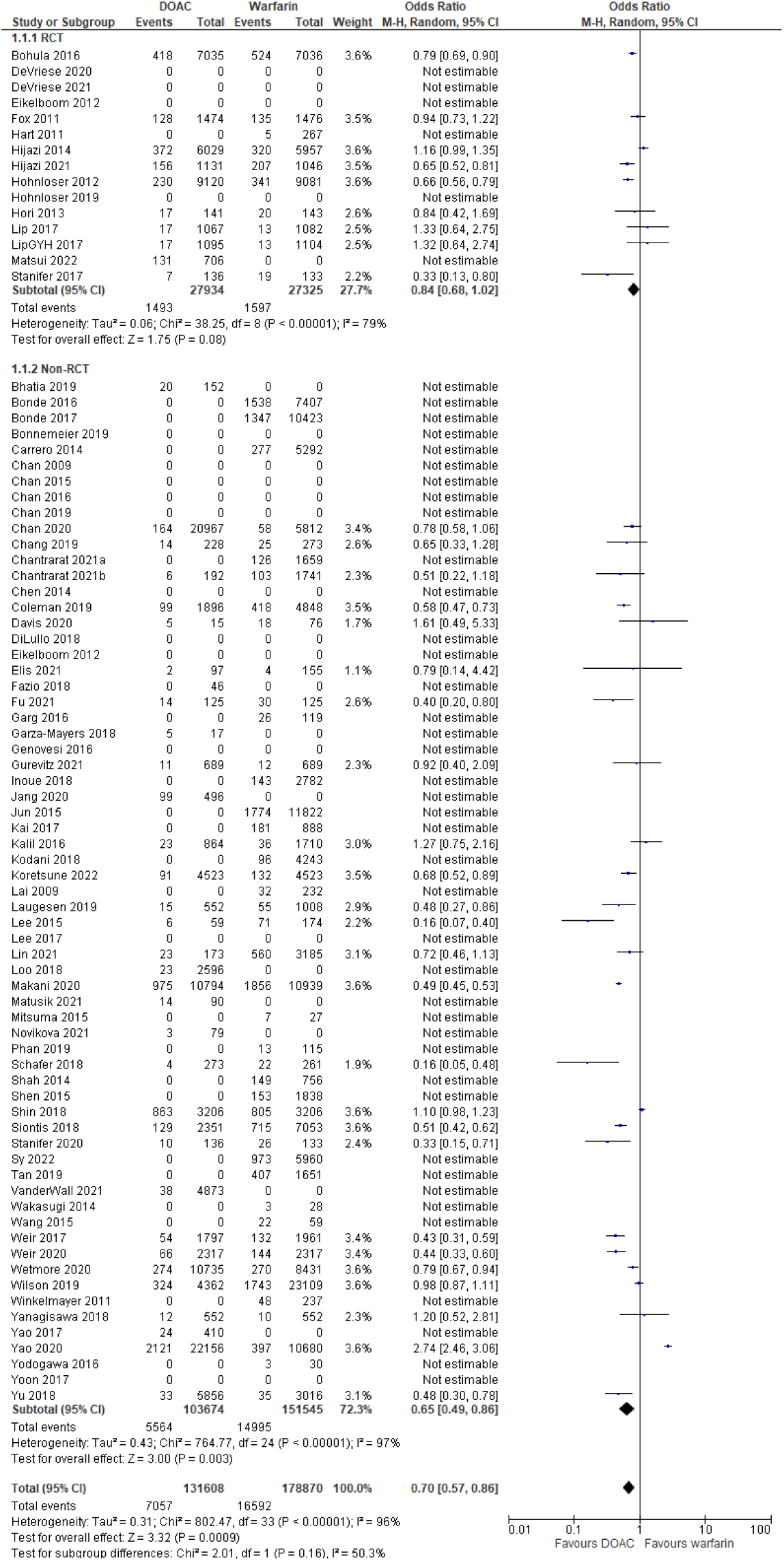
Association between number of overall bleeds and anticoagulation choice of DOAC vs warfarin in patients with concomitant atrial fibrillation and CKD.

Subgroup analyses included studies that reported on numbers of specific etiologies of bleed (GI bleeds or intracranial bleeds) or bleeds that were fatal. Out of the included studies, 2 RCTs (17,021 participants) and 19 non-RCTs (48,705 participants) reported outcomes for number of GI bleeds (**Supplementary Figure 3 in Supplementary Data**). The two RCTs found that DOACs were associated with an increased odds of GI bleeds compared to warfarin (OR 1.37, 95% Cl [1.04, 1.80], p=0.03). In contrast, the non-RCTs found that DOACs were associated with a decreased odd of GI bleeds compared to warfarin (OR 0.68, 95% Cl [0.50, 0.92], p=0.01).

There were 4 RCTs and 14 non-RCTs that examined incidences of intracranial bleeds (**Supplementary Figure 4 in Supplementary Data**). RCTs (29,274 participants) showed that DOAC use over warfarin use had no statistically significant associations with intracranial bleeds (OR 0.67, 95% Cl [0.38, 1.17, p=0.16). In contrast, non-RCTs (33,917 participants) found that DOAC use was associated with decreased incidence of intracranial bleeds (OR 0.52, 95% Cl [0.36, 0.76], p<0.001).

Regarding fatal bleeding events, there were 2 RCTs (14,355 participants) and 3 non-RCTs (total 5,434 participants) that examined this outcome (**Supplementary Figure 5 in Supplementary Data**). We found that DOAC usage was associated with a decreased odds of fatal bleeds in RCTs (OR 0.56, 95% Cl [0.37, 0.85], p=0.007) but not in non-RCTs (OR 0.14, 95% Cl [0.01, 2.63], p=0.19).

#### 3.2.2 Stroke Incidence

Stroke incidence was assessed in 15 RCTs and 40 non-RCTs including 40,147 and 163,081 participants, respectively.

The analyses of both RCTs and non-RCTs found that using DOACs were associated with decreased odds of stroke in both RCTs (OR 0.81; 95% Cl [0.73, 0.90], p<0.001) and non-RCTs (OR 0.60; 95% Cl [0.43, 0.85], p=0.005). (**Figure 3**)

**Figure 3.**
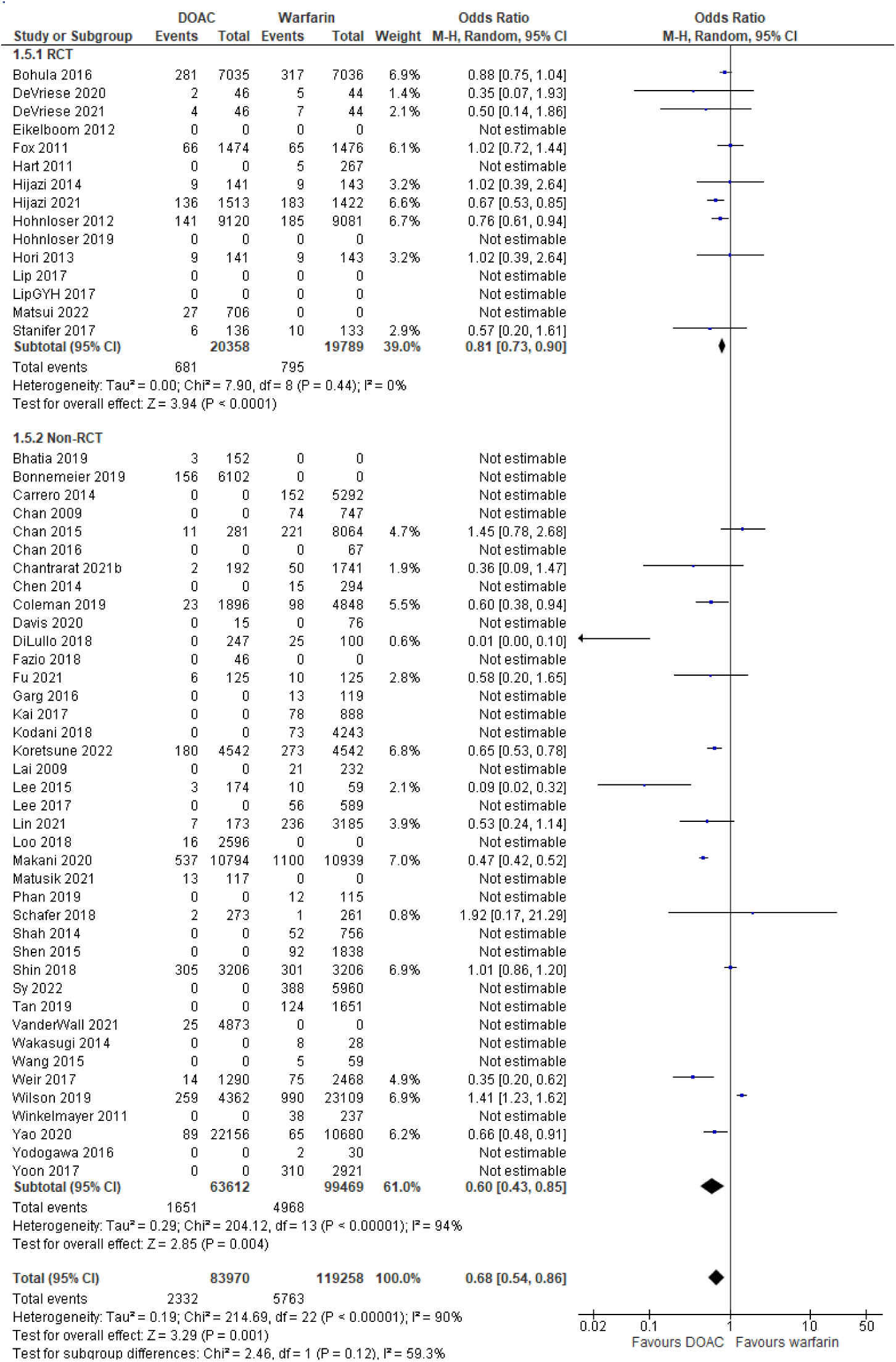
Association between incidence of strokes and anticoagulation choice of DOAC vs warfarin in patients with concomitant atrial fibrillation and CKD.

Five non-RCTs (6,597 participants) examined TIA in particular (**Supplementary Figure 6 in supplementary data**). The meta-analysis showed that, compared to warfarin, DOAC use compared was not associated with a difference in the incidence of TIA (OR 0.36, 95% Cl [0.09, 1.47], p=0.15).

#### 3.2.3 Systemic/arterial embolism and all-cause mortality

There were 13 non-RCTs including 47,687 participants that analyzed the incidence of systemic/arterial embolism (**Supplementary Figure 7 in supplementary data**). Only one RCT assessed this outcome, so a meta-analysis was not done. The meta-analysis of non-RCTs did not find a difference in the incidence of systemic/arterial embolism between DOACs and warfarin (OR 0.49, 95% Cl [0.13, 1.81], p=0.28).

There were 15 RCTs (45,957 total participants) and 32 non-RCTs (127,806 total participants) that assessed the incidence of all-cause mortality (**Supplementary Figure 8 in supplementary data**). The meta-analysis of both RCTs and non-RCTs found that DOACs were associated with decreased odds of all-cause mortality (RCTs OR 0.89, 95% Cl [0.83, 0.96], p=0.003; non-RCTs OR 0.71, 95% Cl [0.61, 0.81], p<0.001).

#### 3.2.4 Sensitivity analyses for individual DOACs

We conducted a sensitivity analysis for comparisons between warfarin and each individual DOAC. In comparisons between warfarin and apixaban, there were 3 RCTs with 20,647 participants and 11 non-RCTs with 48,091 participants that assessed the incidence of major bleeding (**Supplementary Figure 9 in supplementary data**). The results found apixaban usage to be associated with a decreased odds of bleeding in both meta-analyses (RCTs OR 0.64; 95% Cl [0.55, 0.75], p<0.001; non-RCTs OR 0.52; 95% Cl [0.44, 0.63], p<0.001). The incidence of stroke was assessed in 3 RCTs (21,405 participants) and 3 non-RCTs (22,144 participants) (**Supplementary Figure 10 in supplementary data**). Results favoured apixaban in both RCTs (OR 0.71; 95% Cl [0.61, 0.83], p<0.001) and non-RCTs (OR 0.60; 95% Cl [0.41, 0.87], p=0.007).

Regarding comparisons between warfarin with dabigatran, there were 9 non-RCTs that assessed the number of overall bleeds with 42,120 total participants (**Supplementary Figure 11 in supplementary data**). There was no significant difference found between drugs (OR 0.74, 95% Cl [0.53, 1.02], p=0.07). Three non-RCTs with 26,905 total studies compared stroke incidence in warfarin and dabigatran treatments (**Supplementary Figure 12 in supplementary data**). There was no significant difference found (OR 0.97, 95% CI [0.44, 2.13], p=0.94).

In comparison of warfarin with edoxaban, there was no significant difference in the odds of number of bleeding events (**Supplementary Figure 13 in supplementary data**) in RCTs (3 studies with 18,419 participants) (OR 0.97, 95% CI [0.65, 1.44], p=0.88) and non-RCTs (2 studies with 6078 participants) (OR 0.65, 95% CI [0.39, 1.11], p=0.12). Sample was insufficient for assessing stroke incidences.

Finally, for comparisons between warfarin and rivaroxaban (**Supplementary Figure 14 in supplementary data**). Bleeding events were assessed in 5 RCTs (3,940 participants) and 9 non-RCTs (50,725 participants). There was no significant difference in RCTs (OR 0.93, 95% CI [0.73, 1.18], p=0.56) but in non-RCT studies, rivaroxaban was favoured over warfarin (OR 0.79, 95% CI [0.71, 0.87], p<0.001). Stroke incidence was assessed in 5 RCTs with 4,120 participants and 7 non-RCTs with 43,892 participants (**Supplementary Figure 15 in supplementary data**). The RCTs analysis found no significant difference (OR 0.94, 95% CI [0.69, 1.29], p=0.71) while the analysis of non-RCTs favoured rivaroxaban (OR 0.56, 95% CI [0.38, 0.83], p=0.004).

#### 3.2.5 Subgroup analysis by CKD stages

There was a total of 31 non-RCT study including 118,187 patients that assessed number of bleeds for patients of one of more specific stages of CKD. Of those studies, there were 8 studies with 37,718 participants who had stage 1 or 2 CKD, 11 studies with 43,669 patients who had stage 3 CKD, 11 studies with 5,952 patients who had stage 4 or 5 CKD, and 15 studies with 30,848 patients who were on dialysis.

Except for patients with stage 1 or 2 CKD, in all groups DOACs resulted in decreased odds of bleeding over warfarin. Results are shown in **Figure 4**. Each study used some or all of the four DOACs included in the study, although specific data for specific agents were not available. There was no difference in bleeding events in patients with stage 1 or 2 CKD (OR 0.84, 95% CI [0.54, 1.30], p=0.43). DOAC use was associated with decreased odds of bleeding in patients with stage 3 CKD (OR 0.44, 95% CI [0.22, 0.88], p=0.02), stage 4 or 5 CKD (OR 0.66, 95% CI [0.51, 0.86], p=0.002), and patients on dialysis (OR 0.58, 95% CI [0.47, 0.72], p<0.001).

**Figure 4.**
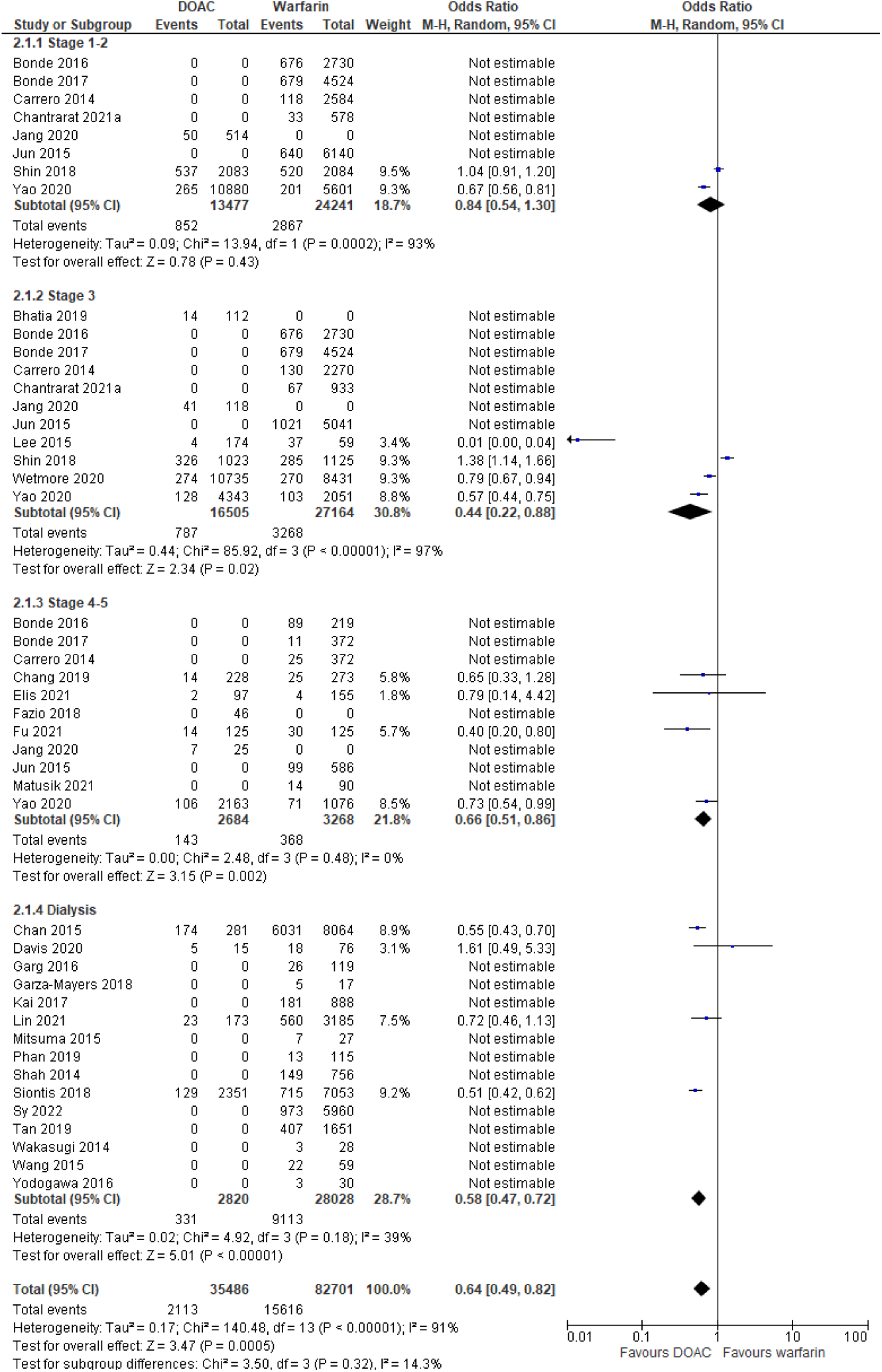
Association between number of bleeds and anticoagulation choice of DOAC versus warfarin in patients with concomitant atrial fibrillation and CKD of stages 1-2, stage 3, stage 4-5, or on dialysis.

There were 31 non-RCT studies with 61,379 total patients that assessed incidence of stroke in patients of one or more specific CKD stages **(Supplementary Figure 16 in supplementary data**). Of these, there were 3 studies with 24,060 patients who had stage 1 or 2 CKD, 5 studies with 11,157 patients who had stage 3 CKD, 13 studies with 7,217 patients who had stage 4 or 5 CKD and 17 studies with 18,945 patients who were on dialysis. The subgroup analyses found no significant difference between DOAC usage and the incidence of stroke for all subgroups: stage 1 or 2 CKD patients (OR 0.96, 95% CI [0.78,1.17], p=0.67), stage 3 CKD patients (OR 0.63, 95% CI [0.26, 1.52], p=0.31), stage 4 or 5 CKD patients (OR 0.75, 95% CI [0.49, 1.17], p=0.20), and dialysis patients (OR 0.90, 95% CI [0.33, 2.46], p=0.83).

### 3.3 Patients with concomitant AF and Valve disease

The analysis of patients with concomitant AF and valve disease assessed 7 RCTs and 23 non-RCTs including a total of 99,299 patients, with 4947 patients included in RCTs and 94,352 patients included in non-RCTs.

#### 3.3.1 Bleeding events

Overall, GI and intracranial bleeds were assessed. Overall bleeding was assessed in 4 RCTs and 11 non RCTs including 6,851 participants in the RCT group and 91,596 in the non-RCT group. There were no significant differences between warfarin and DOACs in the RCTs (OR 0.89, 95% CI [0.61, 1.31], p=0.56), but results favored DOACs in non-RCT studies (OR 0.75, 95% CI [0.57, 0.97], p=0.03). (**Figure 5**)

**Figure 5.**
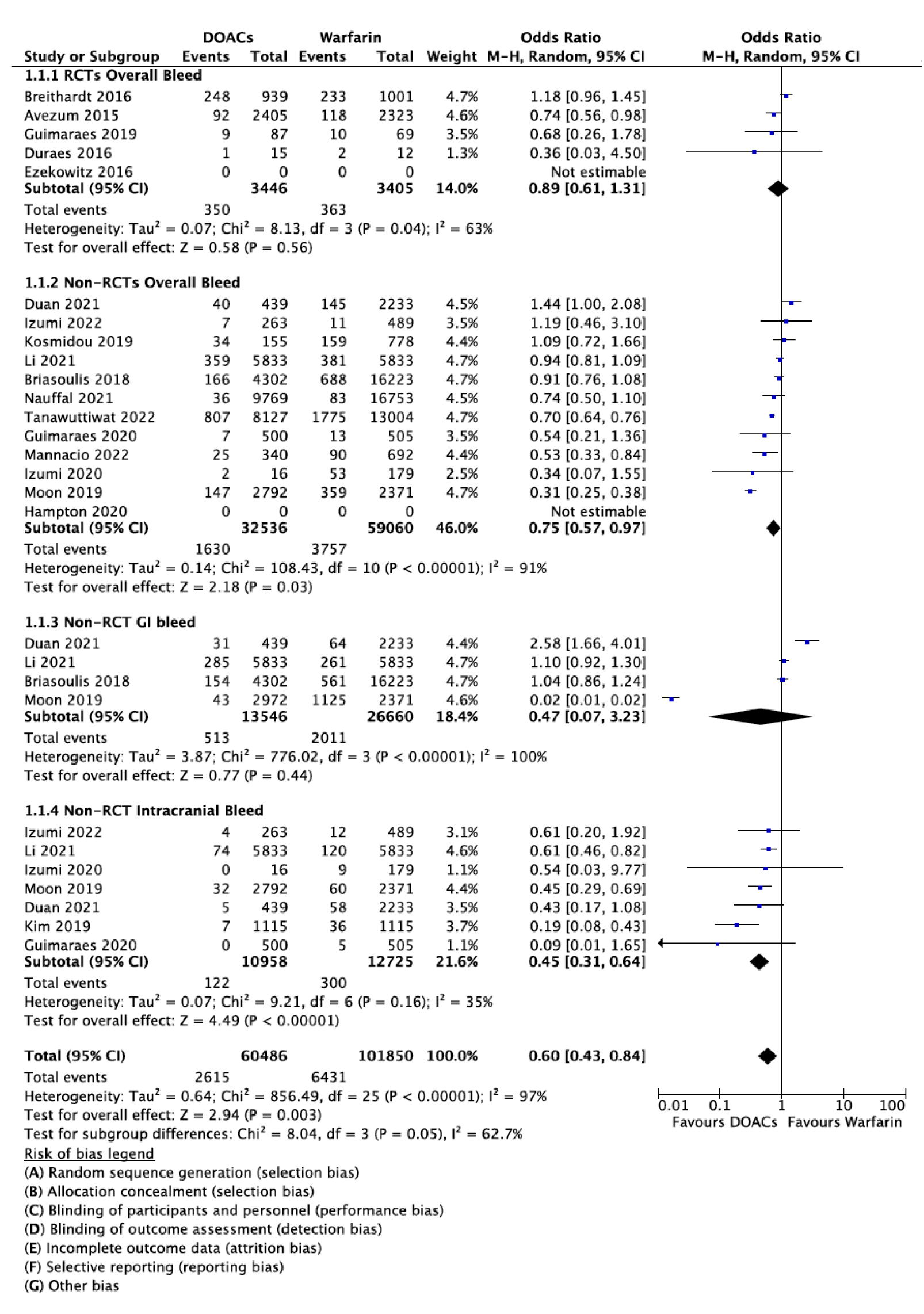
Bleeding outcomes with warfarin vs DOAC in patients with concomitant atrial fibrillation and valve disease.

Non-RCT demonstrated no significant differences between warfarin and DOACs for GI bleed (OR 0.47, 95% Cl [0.07, 3.23], p=0.44) and favored DOAC for intracranial bleeding (OR 0.45, 95% CI [0.31, 0.64], p<0.001). The number of RCTs reporting GI and intracranial bleeding was not sufficient for statistical analysis.

Furthermore, when comparing warfarin and the apixaban, there was no significant differences in overall bleeding in non-RCTs (OR 0.28, 95% Cl [0.01, 9.56], p=0.48) **(Supplementary Figure 17 in supplementary data**). The number of studies assessing other DOACs were not sufficient for individual analyses.

#### 3.3.2 Stroke incidence

The incidence of overall stroke and transient ischemic attack between warfarin and DOAC users was evaluated in 4 RCTs including 4,947 participants and 14 non-RCTs including 94,352 participants in this analysis. For stroke, the RCTs demonstrated no significant difference between warfarin and DOAC (OR 0.86, 95% CI [0.64, 1.14], p=0.29) while non-RCTs favoured DOACs (OR 0.66, 95% CI [0.47, 0.93], p=0.02). Non-RCTs demonstrated no significant difference between warfarin and DOACs for TIA (OR 1.05, 95% CI [0.68, 1.64], p=0.81). (**Figure 6**) The numbers of RCTs reporting TIAs was not sufficient for statistical analysis.

**Figure 6.**
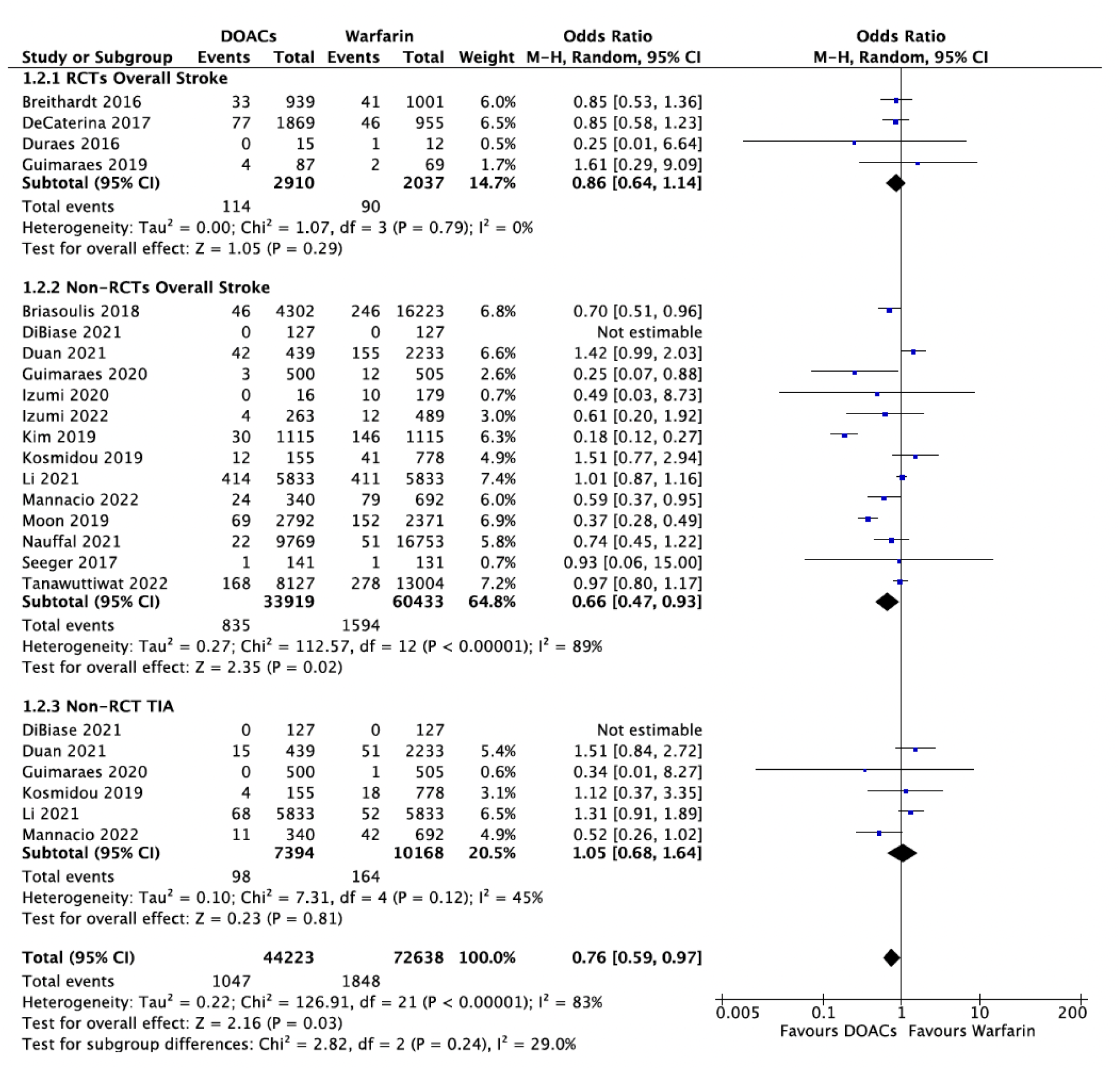
Association between stroke and TIA incidences with anticoagulation choice of DOAC versus warfarin in patients with concomitant atrial fibrillation and valve disease.

#### 3.3.3 Arterial/systemic embolism and all-cause mortality

Non-RCT’s demonstrated no significant differences in arterial/systemic embolisms between warfarin and DOAC users (OR 2.18, 95% CI [0.89, 5.32], p=0.09), but the number of RCTs reporting this outcome was not sufficient for statistical analysis **(Supplementary Figure 18 in supplementary data**). Regarding all-cause mortality, there were no significant differences between warfarin and DOACs in both RCTs (OR 1.01, 95% CI [0.85, 1.19], p=0.94) and non-RCTs (OR 0.77, 95% CI [0.54, 1.09], p=0.14) (**Supplementary Figure 19 in supplementary data**).

## 4. Discussion

Aging patient population often presents with increasing comorbidities, in particular chronic kidney disease (CKD), which notably increases the risk of embolism and hemorrhage.^13^ Additionally, AF is also associated with a higher risk of developing end-stage renal disease in patients with CKD.^14,15^ Thus, it is important to consider how patients with concomitant CKD will respond to anticoagulation therapy compared to those with normal kidney function.^16^ Research into the use of DOACs in valvular AF is also limited.^17^ Unlike mechanical heart valves, bioprosthetic heart valves eliminate the need for lifelong warfarin therapy, but not anticoagulation altogether.^18^ There is a high risk of thromboembolic events within 3-6 months following bioprosthetic valve surgery, and recent studies note that the risk is further increased with transcatheter aortic valve replacement compared to a traditional open-heart procedure.^19,20^ Recently, more studies have started to compare clinical outcomes of DOACs with warfarin in this population of patients with AF and bioprosthetic valve replacement and some results have suggested that DOACs could serve as a preferred alternative to warfarin.^21^

In the present study we conducted and exhaustive systematic review of studies evaluating the use of DOACs in AF patients with CKD and valvular disease since such patients are usually either underrepresented or excluded from large-scale studies. To the best of our knowledge this is the largest and most comprehensive study in this area.

There are several findings from our research. First, in patients with concomitant AF and CKD, we found that DOAC use was associated with a significant reduction in overall bleeding, particularly for patients with more severe CKD (eGFR < 60mL/min/1.73m^2^), as well as a significantly reduction in all-cause mortality. Second, when analyzing specific DOACs in patients with concomitant AF and CKD, apixaban in particular was associated with reduced overall bleeding and stroke incidence. Lastly, in patients with concomitant AF and valvular disease, DOAC use was associated with significant reduction in bleeding and stroke incidence as well. Taken together, these findings suggest that DOACs are probably a reasonable option in these populations, however, these are still considered off-label indications by regulatory authorities.

The majority of our results appear to be in line with previous meta-analyses looking at similar populations. A systematic review and pairwise network meta-analysis by Su *et al.* focused on patients with AF and CKD stage 3-5 who received one of the four DOACs analyzed in our study^22^. They also found DOACs to be superior to warfarin in reducing bleeding events and preventing thromboembolic events. Their comparison of different DOACs found apixaban and edoxaban to be superior for reducing bleeds. Another systematic review and meta-analysis by Chen *et al.* also found improve efficacy of DOACs over warfarin particularly in early CKD, but noted research to be still lacking in patients with stage 4-5 CKD and on dialysis^23^.

Similar to our analysis of patients with concomitant AF and valvular disease, another meta-analysis by Gerfer *et al.* looking specifically at patients with valvular disease who have undergone valve repair or replacement, they found that DOAC reduced major bleeds as well as stroke incidence^24^. However, a separate meta-analysis by Zhang et al. found that DOACs and warfarin usage were associated with similar risks of stroke incidence, while also acknowledging that more trials were needed^25^.

Our study does have some limitations. Due to the nature of a systematic review, we could not ensure that all the individual trials included in our analysis defined relevant outcomes in the same manner. In assessing our outcome of bleeding events, most studies did report major bleeding events specifically in accordance to the criteria of International Society of Thrombosis. However, some studies chose to include clinically relevant non-major bleeding which might make the interpretation of the results more difficult, although these events are widely accepted as appropriate outcomes for studies evaluating anticoagulants. As well, we do acknowledge that there may be some extent of reporting bias present as not all of the studies reported all of the outcomes.

Our study also does identify areas that would benefit from further research. Specifically, there is a relative paucity of data examining specific DOACs or comparisons between them. In patients with concomitant AF and CKD, our study only identified apixaban as superior to warfarin in association with bleeding events, but more trials are needed before a clear clinical recommendation can be made. Similarly, our study identified differences in responses to treatments in patients of specific CKD stages. More studies contrasting patients with early-stage CKD, late-stage CKD, or end-stage renal disease on dialysis will help in forming a stronger conclusion. In summary, our findings suggest that the use of DOACs in patients with CKD, in particular advanced CKD, as well as AF in valvular disease may be appropriate, although these agents are not approved for use in such populations and formal regulatory studies are required.

## Data Availability

All data produced in the present study are available upon reasonable request to the authors.

