## Supplementary Material for "Comparing DOACs with warfarin in AF patients with chronic kidney disease or valvular disease: A systematic review and meta-analysis"

**Supplementary Figure 20**. Funnel plot for the association between number of overall bleeds and anticoagulation choice of DOAC vs warfarin in patients with concomitant atrial fibrillation and CKD. ..49

**Supplementary Figure 21**. Funnel plot for the association between incidence of strokes and anticoagulation choice of DOAC vs warfarin in patients with concomitant atrial fibrillation and CKD. ..50

**Complete search strategies**

**Database: Ovid MEDLINE(R) ALL <1946 to March 28, 2022>**
Search Strategy:
--------------------------------------------------------------------------------
1 atrial fibrillation/ or atrial flutter/ (67692)
2 ((atrial$ or auricular$) adj3 (fibrillation$ or flutter$)).tw,kf. (88133)
3 ((AVAF or AF) and (atrial or flutter)).tw,kf. or (atrial or flutter).ab. /freq=2 (80540)
4 or/1-3 (126427)
5 exp Heart Valve Diseases/ or Heart Valve Prosthesis/ or exp Heart Valve Prosthesis Implantation/ (152939)
6 ((TAVI or TAVR) and (trans$ or (aort$ adj5 valv$) or implant$ or replac$)).tw,kf. (9268)
7 exp heart valves/ or ((heart$ or cardiac$ or cardio$ or mitral$ or aortic or tricuspid or pulmon$) and valv$).tw,kf. (174068)
8 7 and (exp "Prostheses and Implants"/ or (prosthetic$ or prosthes$ or implant$ or graft$ or bioprosthe$ or bio-prosthe$).tw,kf.) (61799)
9 7 and (valv$ adj3 (surg$ or dissect$ or prosthe$ or bioprosthe$ or bio-prosthe$ or implant$ or graft$ or stent$ or artificial$ or repair$ or replac$)).tw,kf. (72193)
10 7 and (disease$ or stenos$ or prolapse$ or regurgitation$).tw,kf. (91450)
11 or/5-6,8-10 (196893)
12 4 and 11 (14321)
13 (((chronic$ or progressive or diabetic) adj (kidney or renal or nephro$ or glomerul$)) or dialy$ or h?emodia$).mp. or ckd.tw. or esrd.tw. or ((diabet$.mp. or Disease Progression/ or Recurrence/) and nephropath$.mp.) or ur?emi$.mp. or m?croalbuminuri$.mp. or albuminuri$.mp. or proteinuri$.mp. or nephrosclerosis.mp. or glomerulosclerosis.mp. or glomerular sclerosis.mp. or *Glomerular Filtration Rate/ or (secondary adj2 hyperparathyroidism).mp. or ((tubulointerstitial or interstitial or renal or kidney) adj fibrosis).tw. or hyperphosphat?emia.tw. or vascular calcification$.tw. or alport$.mp. or denys-drash.mp. or glomerulopathy.tw. or hypoalbumin?emi$.mp. or multicystic kidney$.mp. or polycystic kidney$.mp. or cystic kidney$.mp. or calciphylaxis.mp. or tenckhoff.tw. or ((kidney or renal) adj (disease$ or failur$ or function$ or insufficienc$ or disorder$ or dysfunction or replacement or impairment$)).mp. or ((kidney or renal) and (ckf or crd or crf or eskd or eskf or esrf or hyperparathyroidism or end-stage or endstage or eGFR)).mp. or (((kidney or renal) adj transplant$) and (candidates or wait$ list$)).tw. or ((sclerosi$ or fibrosi$ or fibrotic).mp. and ((ureteral obstruction or nephritis or glomerulonephritis or nephrop$).mp. or (obstruct$ and (kidney$ or renal or nephropathy)).tw.)) or *Creatinine/ or creatinine$.tw,kw,kf. [High-Sensitivity Filter for Ovid Medline Chronic kidney disease] (734754)
14 4 and 13 (7285)
15 12 or 14 (20621)
16 exp warfarin/ or exp Vitamin K/ or (VKA or VKAs or (vitamin$ adj2 K) or warfarin$ or Waran or Coumadin$ or Marevan or bristol-myers squibb).tw,kf. (58335)
17 exp anticoagulants/ or (anticoagula$ or anti-coagulant$ or anti-coagulation$ or anti-thrombin$ or antithrombin$ or NOAC or DOAC or NOACs or DOACs).tw,kf. (288674)

18 (TSOAC or TOAC or TSOACs or TOACs or (thrombin$ adj2 inhibitor$) or (factor$ adj3 Xa adj3 inhibitor$) or (non-VKA or non-VKAs or ((non-vitamin$ or anti-vitamin$) adj2 K) or ((nonvitamin$ or antivitamin$) adj2 K))).tw,kf. (10982)
19 (acenocoumarol or coumarin or phenprocoumon or sintrom or sinthrome or jantoven or nicoumalone or dicoumarol or dicumarol or phenindione or dabigatran or ximelagatran or apixaban or rivaroxaban or edoxaban or betrixaban or idraparinux).tw,kf. (27401)
20 or/17-19 (302092)
21 15 and 16 and 20 (2243)
22 ((systemic$ adj3 embol$) or thrombo?embol$ or micro?embol$).tw,kf. (79122)
23 exp Stroke/ or (stroke$ or ((cerebr$ or brain$) adj3 infarct$) or apoplex$).tw,kf. or Ischemic Attack, Transient/ or (transient$ and ((cerebr$ or brain$ or attack$) adj3 ischemi$)).tw,kf. or TIA.tw,kf. (355768)
24 (hemorrhag$ or haemorrhag$ or bleed$ or rebleed$ or re-bleed$).mp. or (clotting or (blood$ adj clot$)).tw,kf. or exp HEMORRHAGE/ (664442)
25 Comparative Study/ or (vs$1 or vs or versus or compar$ or during-and-after).tw. or (receiv$ adj2 either).tw,kf. (7632574)
26 ((multicenter$ or multi-center$) and ((between or each or both or either or than or across) adj2 group$1)).tw,kf. or groups.tw. or group$1.ab. /freq=2 (2975954)
27 ((atrial$ adj3 fibrillation$) and (VKA or VKAs or (vitamin$ adj2 K) or warfarin$ or Waran or Coumadin$ or Marevan or bristol-myers squibb)).ti. (2234)
28 27 and 20 and 22 and 23 and 24 and (or/25-26) (561)
29 21 or 28 (2675)
30 limit 29 to english language (2468)
31 30 not (exp Animals/ not (Human/ and exp Animals/)) (2465)
32 limit 31 to "all adult (19 plus years)" (1314)
33 limit 31 to "all child (0 to 18 years)" (60)
34 31 not (33 not (32 and 33)) (2464)
35 (mice or rat or rats or cat$1 or cattle$1 or dog$1 or goat$1 or horse$1 or rabbit$1 or sheep$1 or swine$1 or pig$1 or canine$1 or feline$1 or porcine$ or calf).ti. (1840018)
36 (pediatr$ or paediatr$ or child$ or adolescent$ or infan$ or newborn$ or neonat$).ti. (1487544)
37 34 not (or/35-36) (2456)
38 exp case-control studies/ or (case$ and control$).tw. or (case$ and series).tw. [Medline case series ] (1842176)
39 case reports/ or case report$.mp. (2349336)
40 37 not (39 not (38 and 39)) (2285)
**41 40 not case report.ti. [ Removing case Reports and retaining Case Series] (2278)**

***************************

**Database: Embase Classic+Embase <1947 to 2022 March 28>**
Search Strategy:
--------------------------------------------------------------------------------
1 exp *atrial fibrillation/ or *heart atrium flutter/ or *heart atrium fibrillation/ (90273)

2 ((atrial$ or auricular$) adj3 (fibrillation$ or flutter$)).ti. or ((atrial$ or auricular$) adj3 (fibrillation$ or flutter$)).ab. /freq=2 (97038)
3 ((AVAF or AF) and (atrial or flutter)).ti. or ((AVAF or AF) and (atrial or flutter)).ab. /freq=2 (30009)
4 or/1-3 (111322)
5 exp valvular heart disease/ or exp heart valve prosthesis/ or *heart valve replacement/ (214667)
6 ((TAVI or TAVR) and (trans$ or (aort$ adj5 valv$) or implant$ or replac$)).tw,kw. (19888)
7 exp *heart valve/ or ((heart$ or cardiac$ or cardio$ or mitral$ or aortic or tricuspid or pulmon$) and valv$).tw,kw. (235179)
8 7 and (exp *"prostheses and orthoses"/ or (prosthetic$ or prosthes$ or implant$ or graft$ or bioprosthe$ or bio-prosthe$).tw,kw.) (75104)
9 7 and (valv$ adj3 (surg$ or dissect$ or prosthe$ or bioprosthe$ or bio-prosthe$ or implant$ or graft$ or stent$ or artificial$ or repair$ or replac$)).tw,kw. (106304)
10 7 and (disease$ or stenos$ or prolapse$ or regurgitation$).tw,kw. (136947)
11 or/5-6,8-10 (297616)
12 4 and 11 (11358)
13 (((chronic$ or progressive or diabetic) adj (kidney or renal or nephro$ or glomerul$)) or ((kidney$ or renal$) adj disease) or (h?emodial$ or peritoneal dialysis)).mp. or kidney failure/ or dialy$.tw. or ((kidney or renal) adj (failur$ or function$ or insufficienc$ or disorder$ or dysfunction or replacement or damage or impairment$)).tw. or exp *renal replacement therapy/ or (ckd or esrd or ur?emia).mp. or (diabet$ and nephropath$) (mp) or ur?emi$.tw. or (nephrosclerosis or glomerulosclerosis or glomerular sclerosis).mp. or (hyperphosphat?emia or hyperuric?emia or hypoalbumin?emi$).mp. or (secondary adj2 hyperparathyroidism).mp. or m?croalbuminuri$.tw. or ((renal or kidney or vascular) adj calcification$).mp. or exp *glomerulus filtration/ or ((tubulointerstitial or interstitial or renal or kidney) adj fibrosis).mp. or ((multicystic or polycystic or cystic) adj2 kidney).mp. or glomerulopathy.mp. or (alport$ or denys-drash).mp. or *kidney dysfunction/ or ((kidney or renal) and (ckf or crd or crf or eskd or eskf or esrf or hyperparathyroidism or end-stage or endstage or eGFR)).mp. or (((kidney or renal) adj transplant$) and (candidates or wait$ list$)).tw. or ((m?croalbuminuri$ or albuminuri$ or proteinuri$) and (diabet$ or hypertension)).mp. or creatinine$.tw,kw. [High-Sensitivity Filter for Ovid Embase Chronic kidney disease] (1169766)
14 4 and 13 (7978)
15 12 or 14 (17978)
16 *warfarin/ or (VKA or VKAs or (vitamin$ adj2 K) or warfarin$ or Waran or Coumadin$ or Marevan or bristol-myers squibb).tw,kw. or warfarin/cm [Drug Comparison] (96506)
17 exp *anticoagulant agent/ or (anticoagula$ or anti-coagulant$ or anti-coagulation$ or anti-thrombin$ or antithrombin$ or NOAC or DOAC or NOACs or DOACs).tw,kw. or exp anticoagulant agent/cm (417616)
18 antivitamin K/ or (TSOAC or TOAC or TSOACs or TOACs or (thrombin$ adj2 inhibitor$) or (factor$ adj3 Xa adj3 inhibitor$) or (non-VKA or non-VKAs or ((non-vitamin$ or anti-vitamin$) adj2 K) or ((nonvitamin$ or antivitamin$) adj2 K))).tw,kw. (30548)
19 *dabigatran/ or *rivaroxaban/ or *apixaban/ or (acenocoumarol or coumarin or phenprocoumon or sintrom or sinthrome or jantoven or nicoumalone or dicoumarol or dicumarol or phenindione or dabigatran or ximelagatran or apixaban or rivaroxaban or

edoxaban or betrixaban or idraparinux).tw,kw. or dabigatran/cm or rivaroxaban/cm or apixaban/cm (46629)
20 or/17-19 (439589)
21 15 and 16 and 20 (3117)
22 ((systemic$ adj3 embol$) or thrombo?embol$ or micro?embol$).tw,kw. (123526)
23 exp *cerebrovascular accident/ or (stroke$ or ((cerebr$ or brain$) adj3 infarct$) or apoplex$).tw,kw. or transient ischemic attack/ or (transient$ and ((cerebr$ or brain$ or attack$) adj3 ischemi$)).tw,kw. or TIA.tw,kw. (534389)
24 (hemorrhag$ or haemorrhag$ or bleed$ or rebleed$ or re-bleed$ or (clotting or (blood$ adj clot$))).tw,kw. or exp bleeding/ (1352257)
25 exp comparative study/ or (vs$1 or vs or versus or compar$ or during-and-after).tw. or cm.fs. or (receiv$ adj2 either).tw,kw. (10393847)
26 ((multicenter$ or multi-center$) and ((between or each or both or either or than or across) adj2 group$1)).tw,kw. (30130)
27 ((atrial$ adj3 fibrillation$) and (VKA or VKAs or (vitamin$ adj2 K) or warfarin$ or Waran or Coumadin$ or Marevan or bristol-myers squibb)).ti. (3670)
28 27 and 20 and 22 and 23 and 24 and (or/25-26) (991)
29 21 or 28 (3846)
30 limit 29 to english language (3693)
31 30 not ((exp animal/ or nonhuman/) not exp human/) (3687)
32 limit 31 to (adult <18 to 64 years> or aged <65+ years>) (2169)
33 limit 31 to (embryo or infant or child) (15)
34 31 not (33 not (32 and 33)) (3681)
35 (mice or rat or rats or cat$1 or cattle$1 or dog$1 or goat$1 or horse$1 or rabbit$1 or sheep$1 or swine$1 or pig$1 or canine$1 or feline$1 or porcine$ or calf).ti. (2249971)
36 (pediatr$ or paediatr$ or child$ or adolescent$ or infan$ or newborn$ or neonat$).ti. (1916755)
37 34 not (or/35-36) (3676)
38 exp case control study/ or (case$ and control$).tw. or exp case study/ or (case$ and series).tw. [EMBASE case series ] (1235118)
39 case report/ or case report$.tw,kw. (2915284)
40 37 not (39 not (38 and 39)) (3524)
41 40 not case report.ti. [ Removing case Reports and retaining Case Series] (3519)
**42 limit 41 to embase (2009)**

***************************

**Database: EBM Reviews - Cochrane Central Register of Controlled Trials <January 2022>**
Search Strategy:
--------------------------------------------------------------------------------
1 atrial fibrillation/ or atrial flutter/ (5218)
2 ((atrial$ or auricular$) adj3 (fibrillation$ or flutter$)).tw. (13833)
3 ((AVAF or AF) and (atrial or flutter)).tw,kw. or (atrial or flutter).ab. /freq=2 (11078)
4 or/1-3 (15634)

5 exp Heart Valve Diseases/ or Heart Valve Prosthesis/ or exp Heart Valve Prosthesis Implantation/ (2807)
6 ((TAVI or TAVR) and (trans$ or (aort$ adj5 valv$) or implant$ or replac$)).tw. (1090)
7 exp heart valves/ or ((heart$ or cardiac$ or cardio$ or mitral$ or aortic or tricuspid or pulmon$) and valv$).tw. (8130)
8 7 and (exp "Prostheses and Implants"/ or (prosthetic$ or prosthes$ or implant$ or graft$ or bioprosthe$ or bio-prosthe$).tw,kw.) (3166)
9 7 and (valv$ adj3 (surg$ or dissect$ or prosthe$ or bioprosthe$ or bio-prosthe$ or implant$ or graft$ or stent$ or artificial$ or repair$ or replac$)).tw. (5023)
10 7 and (disease$ or stenos$ or prolapse$ or regurgitation$).tw. (4383)
11 or/5-6,8-10 (7944)
12 4 and 11 (1158)
13 (((chronic$ or progressive or diabetic) adj (kidney or renal or nephro$ or glomerul$)) or dialy$ or h?emodia$).mp. or ckd.tw. or esrd.tw. or ((diabet$.mp. or Disease Progression/ or Recurrence/) and nephropath$.mp.) or ur?emi$.mp. or m?croalbuminuri$.mp. or albuminuri$.mp. or proteinuri$.mp. or nephrosclerosis.mp. or glomerulosclerosis.mp. or glomerular sclerosis.mp. or *Glomerular Filtration Rate/ or (secondary adj2 hyperparathyroidism).mp. or ((tubulointerstitial or interstitial or renal or kidney) adj fibrosis).tw. or hyperphosphat?emia.tw. or vascular calcification$.tw. or alport$.mp. or denys-drash.mp. or glomerulopathy.tw. or hypoalbumin?emi$.mp. or multicystic kidney$.mp. or polycystic kidney$.mp. or cystic kidney$.mp. or calciphylaxis.mp. or tenckhoff.tw. or ((kidney or renal) adj (disease$ or failur$ or function$ or insufficienc$ or disorder$ or dysfunction or replacement or damage or impairment$)).mp. or ((kidney or renal) and (ckf or crd or crf or eskd or eskf or esrf or hyperparathyroidism or end-stage or endstage or eGFR)).mp. or (((kidney or renal) adj transplant$) and (candidates or wait$ list$)).tw. or ((sclerosi$ or fibrosi$ or fibrotic).mp. and ((ureteral obstruction or nephritis or glomerulonephritis or nephrop$).mp. or (obstruct$ and (kidney$ or renal or nephropathy)).tw.)) or *Creatinine/ or creatinine$.tw,kw. or Cystatin C/ or cystatin c.tw. or glomerular$ filtration$ rate$.tw. (88135)
14 4 and 13 (1503)
15 12 or 14 (2463)
16 exp warfarin/ or exp Vitamin K/ or (VKA or VKAs or (vitamin$ adj2 K) or warfarin$ or Waran or Coumadin$ or Marevan or bristol-myers squibb).tw. (8063)
17 exp anticoagulants/ or (anticoagula$ or anti-coagulant$ or anti-coagulation$ or anti-thrombin$ or antithrombin$ or NOAC or DOAC or NOACs or DOACs).tw. (21178)
18 (TSOAC or TOAC or TSOACs or TOACs or (thrombin$ adj2 inhibitor$) or (factor$ adj3 Xa adj3 inhibitor$) or (non-VKA or non-VKAs or ((non-vitamin$ or anti-vitamin$) adj2 K) or ((nonvitamin$ or antivitamin$) adj2 K))).tw. (1611)
19 (acenocoumarol or coumarin or phenprocoumon or sintrom or sinthrome or jantoven or nicoumalone or dicoumarol or dicumarol or phenindione or dabigatran or ximelagatran or apixaban or rivaroxaban or edoxaban or betrixaban or idraparinux).tw. (4744)
20 or/17-19 (23208)
21 15 and 16 and 20 (521)
22 ((systemic$ adj3 embol$) or thrombo?embol$ or micro?embol$).tw. (10733)

23 exp Stroke/ or (stroke$ or ((cerebr$ or brain$) adj3 infarct$) or apoplex$).tw. or Ischemic Attack, Transient/ or (transient$ and ((cerebr$ or brain$ or attack$) adj3 ischemi$)).tw. or TIA.tw. (63957)
24 (hemorrhag$ or haemorrhag$ or bleed$ or rebleed$ or re-bleed$).mp. or (clotting or (blood$ adj clot$)).tw. or exp HEMORRHAGE/ (78910)
25 Comparative Study/ or (vs$1 or vs or versus or compar$).tw. (1016018)
26 (((multicenter$ or multi-center$) and (across adj2 group$1)) or groups).tw. or group$1.ab. /freq=2 (727661)
27 ((atrial$ adj3 fibrillation$) and (VKA or VKAs or (vitamin$ adj2 K) or warfarin$ or Waran or Coumadin$ or Marevan or bristol-myers squibb)).ti. (862)
28 27 and 20 and 22 and 23 and 24 and (or/25-26) (297)
29 21 or 28 (733)
30 limit 29 to english language (579)
31 (mice or rat or rats or cat$1 or cattle$1 or dog$1 or goat$1 or horse$1 or rabbit$1 or sheep$1 or swine$1 or pig$1 or canine$1 or feline$1 or porcine$ or calf or (pediatr$ or paediatr$ or child$ or adolescent$ or infan$ or newborn$ or neonat$)).ti. (146435)
**32 30 not 31 (579)**

#### Risk of Bias assessment

**Supplementary Figure 1**. ROBINS-1 Risk of Bias Assessment for non-RCTs

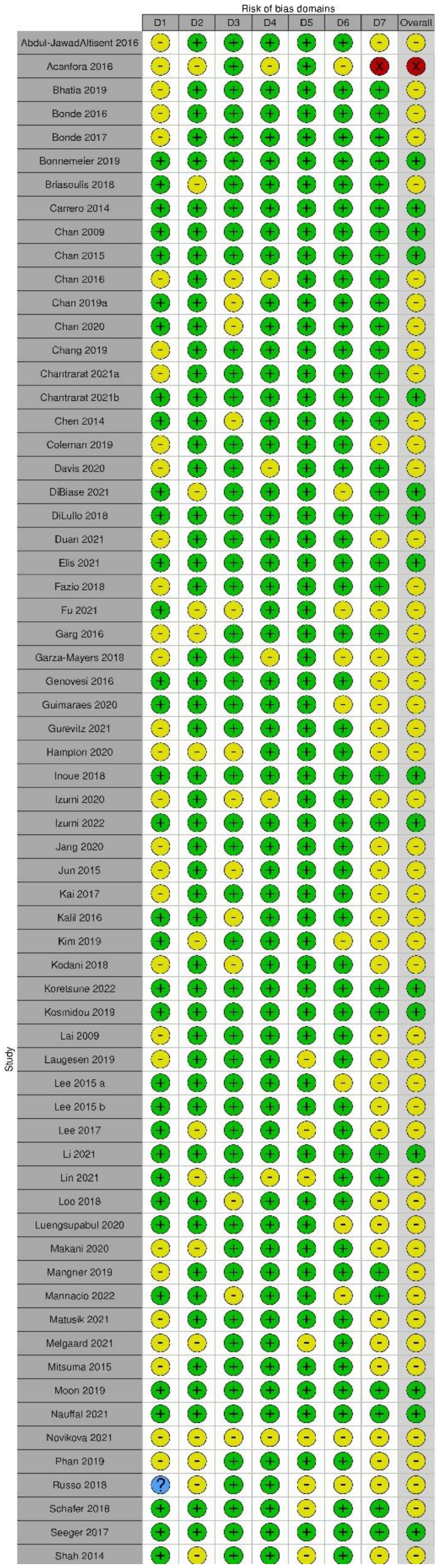

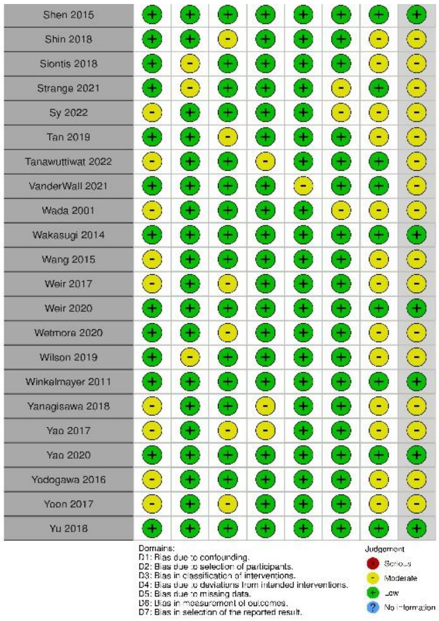
Shen 2015
Shin 2018
Siontis 2018
Strange 2021
Sy 2022
Tan 2019
Tanawuttiwat 2022
VanderWall 2021
Wada 2001
Wakasugi 2014
Wang 2015
Weir 2017
Weir 2020
Wetmore 2020
Wilson 2019
Winkelmayer 2011
Yanagisawa 2018
Yao 2017
Yao 2020
Yodogawa 2016
Yoon 2017
Yu 2018
Domains:
D1: Bias due to confounding.
D2: Bias due to selection of participants.
D3: Bias in classification of interventions.
D4: Bias due to deviations from intended interventions.
D5: Bias due to missing data.
D6: Bias in measurement of outcomes.
D7: Bias in selection of the reported result.
Judgement
Serious
Moderate
Low
No information

**Supplementary Figure 2**. ROB-2 Quality Assessment Scale for RCTs

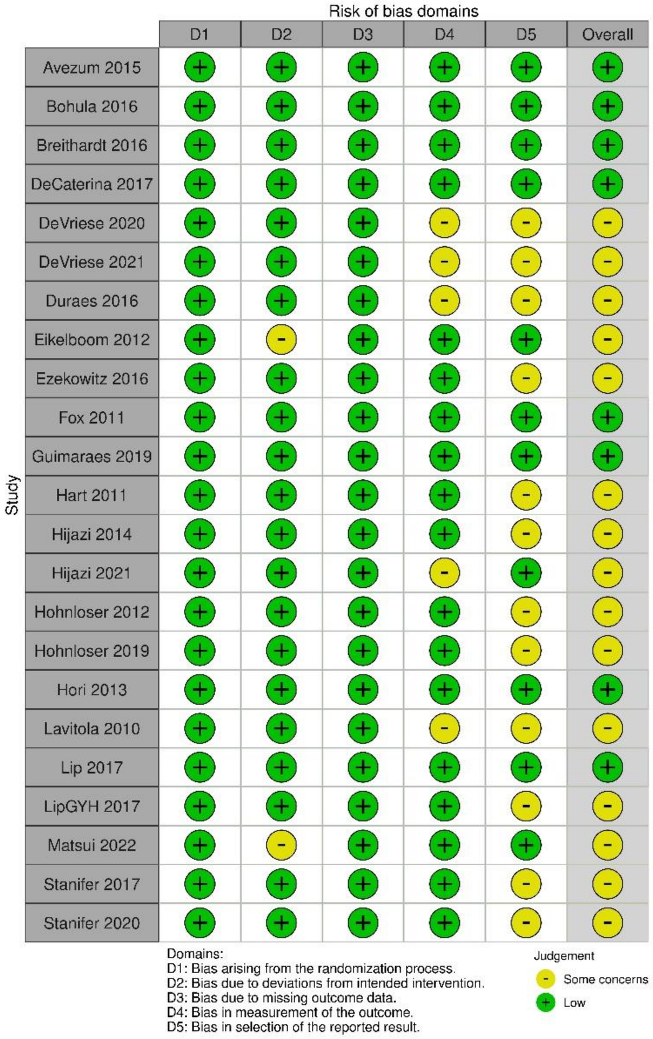

**Supplementary Table 1**. Newcastle-Ottawa Quality Assessment Scale for Cohort Studies

| Study ID | Selection | Comparability | Outcome/Exposure | Total Stars |
|---|---|---|---|---|
| Yao 2017 | 3 | 2 | 2 | 7 |
| Loo 2018 | 3 | 2 | 3 | 8 |
| Weir 2017 | 3 | 2 | 2 | 7 |
| Shin 2018 | 3 | 2 | 3 | 8 |
| Tan 2019 | 3 | 2 | 3 | 8 |
| Yoon 2017 | 3 | 2 | 3 | 8 |
| Guimaraes 2020 | 4 | 2 | 3 | 9 |
| Duan 2021 | 3 | 2 | 3 | 8 |
| VanderWall 2021 | 4 | 1 | 3 | 8 |
| Luengsupabul 2020 | 3 | 0 | 3 | 6 |
| Makani 2020 | 3 | 2 | 3 | 8 |
| Shen 2015 | 3 | 2 | 3 | 8 |
| Hampton 2020 | 3 | 1 | 3 | 7 |
| Weir 2020 | 3 | 2 | 3 | 8 |
| Kosmidou 2019 | 3 | 2 | 3 | 8 |
| Mannacio 2022 | 3 | 2 | 3 | 8 |
| Tanawuttiwat 2022 | 3 | 2 | 3 | 8 |
| Strange 2021 | 3 | 1 | 3 | 7 |
| Izumi 2022 | 4 | 2 | 3 | 9 |
| Melgaard 2021 | 3 | 0 | 3 | 6 |
| Kim 2019 | 3 | 2 | 3 | 8 |
| Lee 2017 | 3 | 2 | 3 | 8 |
| Shah 2014 | 3 | 2 | 2 | 7 |
| Inoue 2018 | 4 | 1 | 3 | 8 |
| Siontis 2018 | 3 | 2 | 2 | 7 |
| Lin 2021 | 3 | 2 | 3 | 8 |
| Fu 2021 | 3 | 2 | 2 | 7 |
| Garg 2016 | 3 | 1 | 3 | 7 |
| Wetmore 2020 | 3 | 2 | 3 | 8 |
| Sy 2022 | 3 | 0 | 3 | 6 |
| Koretsune 2022 | 3 | 2 | 3 | 8 |
| Chen 2014 | 3 | 2 | 3 | 8 |
| Chan 2019 533 | 3 | 2 | 3 | 8 |
| Lee 2015 | 3 | 2 | 3 | 8 |
| Chan 2020 | 3 | 2 | 3 | 8 |
| DiBiase 2021 | 4 | 2 | 3 | 9 |
| Briasoulis 2018 | 3 | 2 | 3 | 8 |

| | | | | |
|---|---|---|---|---|
| Acanfora 2016 | 0 | 1 | 1 | 2 |
| Abdul-JawadAltisent 2016 | 3 | 2 | 3 | 8 |
| Mangner 2019 | 4 | 2 | 2 | 8 |
| Seeger 2017 | 4 | 0 | 3 | 7 |
| Wada 2002 | 3 | 0 | 3 | 6 |
| Russo 2018 | 4 | 0 | 2 | 6 |
| Moon 2019 | 3 | 2 | 3 | 8 |
| Izumi 2020 | 4 | 1 | 3 | 8 |
| Bonde 2017 | 3 | 1 | 3 | 7 |
| Li 2021 | 3 | 2 | 3 | 8 |
| Bhatia 2019 | 3 | 1 | 3 | 7 |
| Chantrarat 2021 272 | 4 | 2 | 3 | 9 |
| Chan 2016 | 3 | 2 | 3 | 8 |
| Chan 2009 | 3 | 2 | 3 | 8 |
| Chan 2015 | 3 | 2 | 3 | 8 |
| Carrero 2014 | 3 | 2 | 3 | 8 |
| Yao 2020 | 3 | 2 | 2 | 7 |
| Wilson 2019 | 3 | 2 | 3 | 8 |
| Genovesi 2016 | 4 | 2 | 3 | 9 |
| Fazio 2018 | 3 | 0 | 2 | 5 |
| Schafer 2018 | 3 | 2 | 2 | 7 |
| DiLullo 2018 | 3 | 2 | 3 | 8 |
| Davis 2020 | 3 | 0 | 3 | 6 |
| Coleman 2019 | 3 | 2 | 3 | 8 |
| Chantrarat 2021 348 | 3 | 2 | 3 | 8 |
| Chang 2019 | 3 | 2 | 3 | 8 |
| Gurevitz 2021 | 4 | 2 | 3 | 9 |
| Bonnemeier 2019 | 3 | 2 | 3 | 8 |
| Garza-Mayers 2018 | 3 | 0 | 2 | 5 |
| Phan 2019 | 3 | 0 | 3 | 5 |
| Novikova 2021 | 4 | 0 | 1 | 5 |
| Lai 2009 | 3 | 0 | 3 | 7 |
| Winkelmayer 2011 | 3 | 2 | 3 | 8 |
| Jun (2015) | 3 | 2 | 3 | 8 |
| Kodani 2018 | 4 | 2 | 3 | 9 |
| Yanagisawa 2018 | 3 | 2 | 2 | 7 |
| Kai 2017 | 3 | 2 | 3 | 8 |

| | | | | |
|---|---|---|---|---|
| Laugesen 2019 | 3 | 2 | 3 | 8 |
| Yu 2018 | 3 | 2 | 2 | 7 |
| Matusik 2021 | 3 | 0 | 3 | 6 |
| Yodogawa 2016 | 3 | 2 | 3 | 8 |
| Wang 2015 | 3 | 2 | 3 | 8 |
| Wakasugi 2014 | 4 | 2 | 3 | 9 |
| Mitsuma 2015 | 3 | 2 | 3 | 8 |
| Lee 2015 | 3 | 2 | 3 | 8 |
| Kalil 2016 | 3 | 2 | 3 | 8 |
| Elis 2021 | 4 | 2 | 3 | 9 |
| Nauffal 2021 | 3 | 2 | 3 | 8 |
| Jang 2020 | 3 | 2 | 2 | 7 |

**Supplementary Table 2**. Jadad Scale Quality Assessment for RCTs

| Study ID | Randomization | Blinding | Account of all Patients | Total |
|---|---|---|---|---|
| Duraes 2016 | 2 | 0 | 1 | 3 |
| Hohnloser 2019 | 2 | 1 | 1 | 4 |
| Guimaraes 2019 | 2 | 1 | 1 | 4 |
| LipGYH 2017 | 2 | 2 | 1 | 5 |
| Eikelboom 2012 | 2 | 2 | 1 | 5 |
| Ezekowitz 2016 | 2 | 2 | 1 | 5 |
| Stanifer 2017 | 2 | 2 | 1 | 5 |
| DeCaterina 2017 | 2 | 2 | 1 | 5 |
| Breithardt 2016 | 2 | 2 | 1 | 5 |
| Avezum 2015 | 2 | 2 | 1 | 5 |
| Lavitola 2010 | 1 | 0 | 1 | 2 |
| DeVriese 2020 | 2 | 2 | 1 | 5 |
| Matsui 2022 | 2 | 1 | 1 | 4 |
| Hijazi 2021 | 2 | 0 | 1 | 3 |
| Hori 2013 | 2 | 2 | 1 | 5 |
| DeVriese 2021 | 2 | 2 | 1 | 5 |
| Fox 2011 | 2 | 2 | 1 | 5 |
| Bohula 2016 | 2 | 2 | 1 | 5 |
| Lip 2017 | 2 | 2 | 1 | 5 |
| Hohnloser 2012 | 2 | 2 | 1 | 5 |
| Hart 2011 | 2 | 0 | 1 | 3 |
| Hijazi 2014 | 2 | 2 | 1 | 5 |
| Stanifer 2020 | 2 | 2 | 1 | 5 |

### Characteristics of included studies

#### Studies in patients with chronic kidney disease

##### Supplementary Table 3. Randomized controlled trials

| Author/Year | Country | Sample Size | Intervention | mean age | % male | Comorbidities: N using concurrent antiplatelet, N heart failure, N CAD, N hypertension N previous stroke, N previous VTE | Included CKD profile | Mean eGFR or CrCl | Outcomes studied |
|---|---|---|---|---|---|---|---|---|---|
| Hohnloser 2019 | international | 347 | | 78.3 | 58.5 | stroke: 38 | CrCl: 30-50, 51-80, >80 | | bleed |
| Eikelboom 2012 | Canada | 2844 | apixaban | 70 | 59.0 | heart failure: 1109, hypertension: 2446; previou stroke: 398 | eGFR: 15-29 (stage 4 CKD), 30-59 (stage 3 CKD), ≥60, | eGFR: 69 | bleed, death |
| Bohula 2016 | USA | 14071 | warfarin (7036), edoxaban (7035) | | 65.4 | heart failure: 10699; CAD: 4686; hypertension: 13152; previous stroke: 3971 | eGFR: 30-50, 51-95, >95 | median: 43 (eGFR 30-50), 69 (eGFR 50-95), 115 (eGFR >95) | bleed, stroke, death |
| Hart 2011 | USA | 538 | warfarin | 73 | 69.0 | CAD: 161; hypertension: 334; stroke: 102 | eGFR: 30-59 (stage 3 CKD) | eGFR: 50 | bleed |
| Hijazi 2021 | USA | 4456 | warfarin, apixaban | 71 | 70.8 | heart failure: 1930; hypertension: 3949; previous stroke: 609 | eGFR: 30-50, 51-80 | | |
| Stanifer 2017 | USA | 269 | warfarin (133), apixaban (136) | 81 | 40.6 (warfarin), 38.2 (apixaban) | concurrent antiplatelet use: 55 (warfarin), 44 (apixaban); heart failure: 54 (warfarin), 77 (apixaban), CAD: 46 (warfarin), 45 (apixaban); hypertension: 118 (warfarin), 119 (apixaban)), previous stroke: 33 (warfarin), 40 (apixaban) | CrCl: 25-30 | CrCl: 27.3 (warfarin), 27.5 (apixaban) | bleed, stroke, death |
| Stanifer 2020 | multicountry (39) | 269 | warfarin (133), apixaban (136) | | 40.6 (warfarin), 38.2 (apixaban) | concurrent antiplatelet use: 58 (warfarin), 46 (apixaban); heart failure: 54 (warfarin), 77 (apixaban); CAD: 46 (warfarin), 45 (apixaban), hypertension: 118 (warfarin), 119 (apixaban), previous stroke: 33 (warfarin), 40 (apixaban) | CrCl: 25-30 | median CrCl: 27.3 (warfarin), 27.5 (apixaban) | bleed, death |
| Fox 2011 | international | 2950 | warfarin (1476), rivaroxaban (1474) | 79 | 44.1 (warfarin), 44 (rivaroxaban) | hypertension: 1360 (warfarin), 1352 (rivaroxaban); previous stroke: 725 (warfarin), 738 (rivaroxaban) | CrCl: 30-49, ≥50 | | bleed, stroke |
| DeVriese 2021 | Belgium | 90 | warfarin (44), rivaroxaban (46) | 80.3 (warfarin), 79.9 rivaroxaban) | 56.8 (warfarin), 76.1 (apixaban) | concurrent antiplatelet use: 14 (warfarin), 15 (rivaroxaban); heart failure: 9 (warfarin), 17 (rivaroxaban), previous stroke: 15 (warfarin), 16 (rivaroxaban) | on hemodialysis | | stroke, arterial/systemic embolism, death |

| | | | | | | | | | |
|---|---|---|---|---|---|---|---|---|---|
| Hohnloser 2012 | USA | 18201 | warfarin (9081), apixaban (9120) | 70 | 65 (warfarin), 86.3 (apixaban) | concurrent antiplatelet use: 2773 (warfarin), 2859 (apixaban); heart failure: 3216 (warfarin), 3235 (apixaban); hypertension: 7954 (warfarin), 7962 (apixaban); previous stroke: 1790 (warfarin), 1748 (apixaban) | eGFR: <50, 51-80 | | bleed, stroke, death |
| LipGYH 2017 | UK | 2199 | warfarin (1104), endoxaban (1095) | 64.2 (warfarin), 64.3 endoxaban) | 65.4 (warfarin), 65.8 (endoxaban) | heart failure: 484 (warfarin), 476 (edoxaban), CAD: 197 (warfarin), 181 (edoxaban); hypertension: 864 (warfarin), 850 (edoxaban), previous stroke: 66 (warfarin), 68 (edoxaban) | CrCl: 15-30, 31-50, 51-80, >80, >95 | CrCl: 94.3 (warfarin), 94.1 (edoxaban) | bleed |
| Hori 2013 | Japan | 284 | warfarin (143), rivaroxaban (141) | 78 | 66.4 (warfarin), 74.5 (rivaroxaban) | concurrent antiplatelet use: 57 (warfarin), 61 (rivaroxaban); heart failure: 67 (warfarin), 74 (rivaroxaban); hypertension: 118 (warfarin), 116 (rivaroxaban) | CrCl: 30-49, ≥50 | | bleed, stroke, death |
| Lip 2017 | multicentre: Denmark, UK, US, Poland, germany | 2149 | warfarin (+enoxaparin) (1082), edoxaban (1067) | 64.2 (warfarin), 64.3 endoxaban) | 66.7 (warfarin), 67.6 (edoxaban) | Heart failure: 484 (warfarin), 476 (edoxaban) CAD: 197 (warfarin), 181 (edoxaban); hypertension: 864 (warfarin), 850 (edoxaban); previous stroke: 66 (warfarin), 68 (edoxaban) | CrCl: 15-30, 31-50, 51-80, >80, >95 | | bleed |
| Hijazi 2014 | multicountry (44) | 17951 | warfarin (5965), dabigatran (5957) | | | heart failure, CAD, hypertension, previous stroke | eGFR: ≤50, 51-80, >80 | | bleed, death |

#### Supplementary Table 4. Non-randomized studies

| Author /Year | Study Design | Country | Sample Size | Intervention | mean age | % male | Comorbidities: N using concurrent antiplatelet, N heart failure, N CAD, N hypertension N previous stroke, N previous VTE | Included CKD profile | Mean eGFR or CrCl | Outcomes studied |
|---|---|---|---|---|---|---|---|---|---|---|
| Bhatia 2019 | retrospective cohort | USA | 152 | DOAC (rivaroxaban: 33; apixaban: 97; dabigatran: 20; edoxaban: 2) | 78.6 | 66.4 | concurrent antiplatelet use: 69; heart failure: 57; hypertension: 135; previous stroke: 37 | CKD stage 3, CKD stage 4/5 | CrCl: 38.8 | bleed, stroke, death |
| Bonde 2016 | retrospective cohort | denmark | 7407 | warfarin | 61 (eGFR>90), 70 (eGFR 60-89), 80 (eGFR 30-59), 83 (eGFR-29), 78 (<15) | | | eGFR: >90, 60-89, 30-59, 16-29, <15 | median: eGFR>90: 98 , eGFR>60-89: 73, eGFR 30-59: 51, eGFR 15-29: 27, eGFR<15: 14 | bleed |

| | | | | | | | | | | |
|---|---|---|---|---|---|---|---|---|---|---|
| Bonde 2017 | retrospective cohort | Denmark | 10423 | warfarin | 68 (eGFR>=60), 77 (eGFR 30-59), 80 (eGFR<30) | 107.3 | concurrent antiplatelet use: 3618; heart failure: 1915; hypertension: 5880; previous stroke: 2310 | eGFR: ≥60, 30-59, <30 | eGFR≥60: 73.2, eGFR 30-59: 47.9, eGFR<30: 25 | bleed |
| Bonnemeier 2019 | retrospective cohort | Germany | 6102 | rivaroxaban | 76.9 | 45.5 | concurrent antiplatelet use: 2135; heart failure: 3175; CAD: 2939; hypertension: 5589; previous stroke: 943 | renal impairment as defined by ICD-10 codes | | bleed, stroke |
| Carrero 2014 | retrospective cohort | Sweden | 5292 | warfarin | 78 | 63.9 | concurrent antiplatelet use: 3807; heart failure: 2677; hypertension: 2869; previous stroke: 1398 | eGFR: >60, 30-60, 15-30, <15 | | bleed, stroke, death |
| Chan 2009 | retrospective cohort | USA | 747 | warfarin | 72.6 | 57.8 | heart failure: 436; CAD: 311; hypertension: 595; previous stroke: 108 | patients on hemodialysis | | stroke |
| Chan 2015 | retrospective cohort | USA | 8345 | warfarin (8064), dabigatran (281) | 70.6 (warfarin), 68.4 (dabigatran) | 61.2 | concurrent antiplatelet use: 250 (warfarin), 16 (dabigatran); heart failure: 1677 (warfarin), 41 (dabigatran); hypertension: 7137 (warfarin), 244 (dabigatran); previous stroke: 1024 (warfarin), 35 (dabigatran); | patients on chronic hemodialysis | | bleed, stroke, arterial/systemic embolism |
| Chan 2016 | retrospective cohort | China | 67 | Warfarin | 69.5 | 58.2 | heart failure: 20; CAD: 24; hypertension: 42; previous stroke: 12 | patients on peritoneal dialysis | | stroke |
| Chan 2019 a | retrospective cohort | Taiwan | 2622 | rivaroxaban | 72.44 | 66.5 | concurrent antiplatelet use: 946; heart failure: 328; hypertension: 1736; previous stroke: 587 | eGFR: >50, ≤50 | | bleed |
| Chan 2020 | retrospective cohort | Taiwan | 5812 | warfarin (5812), apixaban/edoxaban/dabigatran/rivaroxaban (20967) | 72.5 (warfarin), 75.1 (DOACs) | 52.3 | concurrent antiplatelet use: 2603 (warfarin), 3929 (DOAC); heart failure: 919 (warfarin), 2621 (DOAC); hypertension: 4446 (warfarin), 16989 (DOAC); previous stroke: 1034 (warfarin), 4971 (DOAC) | CKD (not further specifications) | | bleed |
| Chang 2019 | retrospective cohort | taiwan | 520 | warfarin (520), apixaban/dabigatran/rivaroxaban/edoxaban (280) | | 44.6 | concurrent antiplatelet use: 496 (warfarin), 173 (DOAC); heart failure: 317 (warfarin), 142 (DOAC); hypertension: 428 (warfarin), 231 (DOAC); previous stroke: 59 (warfarin), 36 (DOAC) | eGFR: 15-29 (stage 4), <15 (stage 5) or on dialysis | eGFR: 17.3 (warfarin); 25.15 (DOAC) | bleed |
| Chantrarat 2021 a | proospective cohort | Thailand | 1659 | Warfarin | 68.5 | 57.7 | concurrent antiplatelet use: 227; heart failure: 521; CAD: 289; hypertension: 1242 | CKD 1-2, CKD 3, CKD 4-5 | | bleed, death |
| Chantrarat 2021 b | retrospective cohort | Thailand | 1933 | warfarin (1741), OAC (192) | 67.7 | | | eGFR<60 | | bleed, stroke |
| Chen 2014 | retrospective cohort | Taiwan | 294 | no warfarin (2983), warfarin (294) | | 41.5 | heart failure: 1554 (no warfarin), 170 (warfarin); CAD: 1613 (no warfarin), 203 (warfarin); hypertension: 2479 (no warfarin), 238 (warfarin) | ESRD requiring renal replacement therapy | | stroke or TIA |

| | | | | | | | | | |
|---|---|---|---|---|---|---|---|---|---|
| Coleman 2019 | retrospective cohort | Germany | 6744 | warfarin (4848), rivaroxaban (1896) | 72 | 61.6 (warfarin), 58.4 (rivaroxaban) | concurrent antiplatelet use: 887 (warfarin), 212 (rivaroxaban); heart failure: 2477 (warfarin), 944 (rivaroxaban); hypertension: 4286 (warfarin), 972 (rivaroxaban); | CKD stage 4 and 5 | | bleed, stroke |
| Davis 2020 | retrospective cohort | USA | 76 | warfarin (76), apixaban/dabigatran (15) | 68.3 (warfarin), 73 (DOAC) | 52.6 | | ESRD requiring renal replacement therapy | | stroke, arterial/systemic embolism |
| DiLullo 2018 | retrospective cohort | Italy | 347 | warfarin (100), rivaroxaban (247) | 66.5 (warfarin), 66 (rivaroxaban) | 58 (warfarin), 54.3 (rivaroxaban | hypertension: 100 (warfarin), 240 (rivaroxaban); previous stroke: 0 (warfarin), 0 (rivaroxaban) | stage 3b-4 (eGFR: 15-45) | eGFR: 38.4 (warfarin); 37.7 (rivaroxaban) | bleed, stroke, arterial/systemic embolism |
| Elis 2021 | prospective cohort | | 252 | warfarin (155), apixaban (97) | 76.6 (warfarin), 82.1 (apixaban) | 56.8 | heart failure: 84 (warfarin), 63 (apixaban); CAD: 64 (warfarin), 46 (apixaban); hypertension: 91 (warfarin), 66 (apixaban); previous stroke: 24 (warfarin), 15 (apixaban); | 15 ml/min/BSA < eGFR MDRD < 30 ml/min/BSA. | eGFR: 24.7 | bleed |
| Fazio 2018 | retrospective cohort | Italy | 46 | edoxaban | 84.6 | 37.0 | heart failure: 15; hypertensino: 46; previous stroke: 7 | eGFR: 15-29 | eGFR (mL/min): 22.9 | bleed, stroke, arterial/systemic embolism, death |
| Fu 2021 | retrospective cohort | Taiwan | 3250 | warfarin (1625), apixaban (1625) | | 57 (warfarin), 57.7 (apixaban) | concurrent antiplatelet use: 708 (warfarin), 680 (apixaban); heart failure: hypertension: 537 (warfarin), 519 (apixaban) | | advanced CKD: eGFR <30 | bleed, stroke, death, arterial/systemic embolism |
| Garg 2016 | retrospective cohort | USA | 119 | warfarin | 75 | 55.5 | concurent antiplatelet use: 90; heart failure: 107; CAD: 92; hypertension: 101; previous stroke: 24 | patients on chronic hemodialysis | | bleeding, stroke, death |
| Garza-Mayers 2018 | retrospective cohort | USA | 20 | apixaban | 62.45 | 45 | | patients on hemodialysis, peritoneal dialysis | | bleed |
| Genovesi 2016 | prospective cohort | Italy | 134 | warfarin | 76 | 64.2 | concurrent antiplatelet use: 32; heart failure: 58; hypertension: 102; stroke: 21 | patients with ESRD on hemodialysis | | bleed, death |
| Gurevitz 2021 | prospective cohort | USA | 1378 | warfarin (689), apixaban (689) | 79.5 (warfarin), 79.4 (apixaban) | 53.6 | concurrent antiplatelet use: 238 (warfarin), 237 (apixaban); CAD: 276 (warfarin), 302 (apixaban); hypertension: 392 (warfarin), 465 (apixaban); previous stroke: 134 (warfarin), 137 (apixaban); | eGFR < 60 | eGFR: 46.74 (warfarin), 47.27 (apixaban) | bleed, death |
| Inoue 2018 | prospective cohort | Japan | 2782 | warfarin | 75 | 64.8 | concurrent antiplatelet use: 707; heart failure: 922; CAD: 377; hypertension: 1855; previous stroke: 468 | CrCl: >80, 51-80, 30-50, <30 | CrCl (m:/min): 54 | bleeding, arterial/systemic embolism, death |

| | | | | | | | | | | |
|---|---|---|---|---|---|---|---|---|---|---|
| Jang 2020 | retrospective cohort | USA | 496 | apixaban (245), rivaroxaban (204), dabigatran (47) | | | | eGFR>60 (non-CKD); eGFR 30-59.9 (CKD stage 3); 15-29.9 (CKD stage 4); <15 (CDK stage 5) | | bleed |
| Jun (2015) | retrospective cohort | Canada | 11822 | warfarin | | 50.3 | concurrent antiplatelet use: 163; heart failure: 4821; hypertension: 1791 | eGFR: 60-90, 50-80, 30-50, <30 | | bleed |
| Kai 2017 | retrospective cohort | USA | 888 | warfarin | 68.9 | 61.6 | concurrent antiplatelet use: 122; heart failure: 656; CAD: 482; hypertension: 881; previous stroke: 213 | patients on hemodialysis | | bleed, stroke, death |
| Kalil 2016 | retrospective cohort | USA | 2574 | warfarin (1710), dabigatran (864) | | | | eGFR<50, 50-80, >80 | | bleed, death |
| Kodani 2018 | prospective cohort | Japan | 4243 | warfarin | | 66.4 | concurrent antiplatelet use: 1610; CAD: 676; hypertension: 2996 | CrCl: 50-80, 30-49, <30 | | bleed, stroke, TIA, arterial/systemic embolism, death |
| Koretsune 2022 | retrospective cohort | Japan | 9046 | warfarin (4523), apixaban (4523) | 72.5 (warfarin), 73 (apixaban) | 61 (warfarin), 60 (apixaban) | concurrent antiplatelet use: 125 (warfarin); 1163 (apixaban); heart failure: 1312 (warfarin), 1253 (apixaban); CAD: 756 (warfarin), 748 (apixaban); hypertension: 3482 (warfarin), 3482 (apixaban) | CrCl: >50, CrCl ≤50 | | bleed, stroke |
| Lai 2009 | retrospective cohort | USA | 232 | warfarin | 73 | 71.1 | concurrent antiplatelet use: 92; CAD: 145; hypertension: 148; previous stroke: 19 | | | bleed |
| Laugesen 2019 | retrospective cohort | Denmark | 1560 | warfarin (1008), apixaban/dabigatran/rivaroxaban (552) | 78 (warfarin), 80 (DOAC) | 64.0 | concurrent antiplatelet use: 537 (warfarin), 277 (DOAC); heart failure: 395 (warfarin), 195 (DOAC); hypertension: 914 (warfarin), 494 (DOAC); stroke: 175 (warfarin), 102 (DOAC) | CKD patients not no dialysis | | arterial/systemic embolism, death |
| Lee 2015 | retrospective cohort | South Korea | 1319 | warfarin (993), DOAC (326) | 69.3 (warfarin), 71.9 (DOAC) | 34.1 (warfarin), 37.7 (DOAC) | heart failure: 91 (warfarin), 46 (DOAC); hypertension: 517 (warfarin), 229 (DOAC); previous stroke: 320 (warfarin), 215 (DOAC) | eGFR: <60 | | bleed, stroke |
| Lee 2015 | retrospective cohort | South Korea | 174 | warfarin (174), dabigatran/rivaroxaban (59) | | | | eGFR <60 | | bleed, stroke, death |
| Lee 2017 | retrospective cohort | Taiwan | 589 | warfarin | 69.3 | 41.3 | concurrent antiplatelet use: 336; heart failure: 186; previous stroke: 39 | patients on hemodialysis | | stroke, death |
| Lin 2021 | retrospective cohort | Taiwan | 3358 | warfarin (3185), rivaroxaban (173) | 69 (warfarin), 75 (rivaroxaban) | 51 (warfarin), 54.9 (rivaroxaban) | concurrent use of antiplatelet: 1752 (warfarin), 99 (rivaroxaban); heart failure: 1178 (warfarin), 57 (rivaroxaban); hypertension : 2484 (warfarin), 142 (apixaban); previous stroke: 414 (warfarin), 33 (rivaroxaban) | | ESDR patients: stage 5 CKD on dialysis | bleed, arterial/systemic embolism, death |

| | | | | | | | | | | |
|---|---|---|---|---|---|---|---|---|---|---|
| Loo 2018 | retrospective cohort | UK | 2596 | dabigatran, rivaroxaban, apixaban, edoxaban | 77.62 | 53 | concurrent antiplatelet use: 1681; heart failure: 265; CAD: 376; hypertension: 2104; previous stroke: 227; previous VTE: 51 | | | bleed, stroke |
| Makani 2020 | retrospective cohort | | 21733 | warfarin (10939), DOAC (10794) | 76.4 (warfarin), 61.2 (DOAC) | | heart failure: 3527 (warfarin), 2473 (DOAC); CAD: 3590 (warfarin), 3202 (DOAC); hypertension: 8033 (warfarin), 8362 (DOAC); previous stroke: 1640 (warfarin), 1720 (DOAC); previous VTE: 414 (warfarin), 281 (DOAC) | eGFR : ≤30 or on dialysis, 31-60, >60 | | bleed, stroke |
| Matsui 2022 | retrospective cohort | Japan | 706 | rivaroxaban | 80.3 | 66.0 | concurrent antiplatelet use: 253; heart failure: 336; previous stroke: 539; previous VTE: 112 | eGFR: <50 | eGFR (mL/min): 38.7 | bleed, stroke, arterial/systemic embolism, death |
| Matusik 2021 | prospective cohort | Poland | 90 | NOAC (apixaban, rivaroxaban) | median: 71 | | | stage 4 CKD | median eGFR: 24 | bleed, stroke or TIA |
| Mitsuma 2015 | retrospective cohort | Japan | 27 | warfarin | 69.4 | 74.1 | concurrent antiplatelet use: 13; heart failure: 16; CAD: 10; hyerptension: 10; previous stroke: 4 | patients on maintenance hemodialysis | | bleed, stroke, TIA arterial/systemic embolism, death |
| Novikova 2021 | prospective cohort | Russia | 79 | dabigatran | 64 | 36.7 | concurrent antiplatelet use: 20; heart failure: 14; hypertension: 51; previous stroke: 49 | eGFR: <90 | eGFR: 68 | bleed, death |
| Phan 2019 | retrospective cohort | USA | 115 | warfarin | 67.3 | 58.3 | concurrent antiplatelet use: 16; heart failure: 63; CAD: 53; hypertension: 115; previous stroke: 24 | patients on peritoneal dialysis | | bleed, stroke, death |
| Schafer 2018 | retrospective cohort | USA | 534 | warfarin (261), apixaban (273) | 70.6 (warfarin), 73.5 (apixaban) | 62.5 (warfarin), 50.9 (apixaban) | concurrent antiplatelet use: 152 (warfarin), 147 (apixaban); hypertension: 196 (warfarin), 171 (apixaban); previous stroke: 61 (warfarin), 58 (apixaban) | CKD stage 4, stage 5 (includes hemodialysis) | eGFR: 20.4, CrCl: 25.2 (warfarin); eGFR: 22, CrCl: 25.2 (apixaban) | bleed, stroke, arterial/systemic embolism |
| Shah 2014 | retrospective cohort | Canada | 756 | warfarin | 75.3 | 60.7 | concurrent antiplatelet use: 196; heart failure: 312; CAD: 470; hypertension: 582; previous stroke: 42 | patients on dialysis (hemodialysos or peritoneal dialysis) | | bleeding, stroke |
| Shen 2015 | retrospective cohort | USA | 1838 | wafarin | 61.2 | | concurrent antiplatelet use: 395; CAD: 1237; hypertension:1787 | | | bleed, stroke |
| Shin 2018 | retrospective cohort | USA | 6412 | warfarin (3206), dabigatran/rivaroxaban/apixaban (3206) | 72 (warfarin), 73 (DOAC) | 54 (warfarin), 53 (DOAC) | concurrent antiplatelet use: 737 (warfarin), 705 (DOAC); heart failure: 994 (warfarin), 994 (DOAC); CAD: 1314 (warfarin), 1314 (DOAC); hypertension: 2661 (warfarin), 2661 (DOAC); previous stroke: 288 (warfarin), 288 (DOAC); previous VTE: 481 (warfarin), 481 (DOAC) | eGFR: > 60 and ≤60 | | bleed, stroke |

| | | | | | | | | | | |
|---|---|---|---|---|---|---|---|---|---|---|
| Siontis 2018 | retrospective cohort | USA | 9404 | warfarin (7053), apixaban (2351) | 68.2 (warfarin), 68.9 (apixaban) | 54.3 (warfarin), 54.4 (apixaban) | concurrent antiplatelet use: 522 (warfarin), 154 (apixaban); heart failure: 5644 (warfarin), 1868 (apixaban); hypertention: 7025 (warfarin), 2342 (apixaban); previous VTE: 1333 (warfarin), 279 (apixaban) | ESRD patients on dialysis (hemodialysis or peritoneal dialysis) | | bleed, death |
| Sy 2022 | retrospective cohort | USA | 5960 | warfarin | 74 | 95.9 | heart failure: 5171; hypertension: 5911 | patients on hemodialysis | | bleed, stroke |
| Tan 2019 | retrospective cohort | USA | 1651 | warfarin | 73.9 | 43.6 | concurrent antiplatelet use: 428; heart failure: 1071; hypertension: 1620 | ESRD dialysis patients | | bleed |
| Vander Wall 2021 | prospective cohort | international | 4873 | Dabigatran | 64 (CrCl >80), 74 (CrCl 50-79), 80 (CrCl 30-49) | | concurrent antiplatelet use: 608 | CrCl: >80, 50-79, 30-49 | | bleed, stroke, death |
| Wakasugi 2014 | prospective cohort | Japan | 28 | warfarin | 67.8 | 57.1 | concurrent antiplatelet use: 17; heart failure: 28; CAD: 4; previous stroke: 5 | patients with end-stage kidney disease requiring hemodialysis | | bleed, stroke, death |
| Wang 2015 | retrospective cohort | new zealand | 59 | warfarin | 59.8 | 61.0 | concurrent antiplatelet use: 27; heart failure: 16; CAD: 42; hypertension: 58 | patients with end-stage renal disease who started hemodialysis or peritoneal dialysis | | bleed, stroke, arterial/systemic embolism, death |
| Weir 2017 | retrospective cohort | USA | 3758 | warfarin (1961), rivaroxaban (1797) | | 55.8 (warfarin), 56.3 (rivaroxaban) | heart failure: 587 (warfarin), 517 (rivaroxaban); hypertension: 451 (warfarin), 420 (rivaroxaban) | CrCl: >80, 50-80, <50 | | bleed, stroke |
| Weir 2020 | retrospective cohort | USA | 2317 | warfarin (1536), rivaroxaban (781) | 79.9 | 40.8 (warfarin), 38.4 (rivaroxaban) | concurrent antiplatelet use: 783 (warfarin), 391 (rivaroxaban); hypertension: 1441 (warfarin), 734 (rivaroxaban) | CrCl: ≥15 and <30 (stage 4 CKD), <15 (stage 5 CKD) without dialysis, <15 (stage 5 CKD) with dialysis | | bleed |
| Wetmore 2020 | retrospective cohort | USA | 22739 | warfarin (10529), apixaban (6738), rivaroxaban (3904), dabigatran (1568) | 78.4 (warfarin), 79.9 (apixaban), 78.7 (rivaroxaban), 77.9 (dabigatran) | 49 (warfarin), 48 (apixaban), 50.3 (rivaroxaban), 49 (dabigatran) | concurrent antiplatelet use: 1748 (warfarin), 1125(apixaban), 625(rivaroxaban), 259 (dabigatran); heart failure: 6517 (warfarin), 3537 (apixaban), 1925 (rivaroxaban), 768 (dabigatran); hypertension: 10318 (warfarin), 6596 (apixaban), 3807 (rivaroxaban), 1537 (dabigatran) | stage 3 CKD, stage 4-5 CKD | | bleed, death |

| | | | | | | | | | | |
|---|---|---|---|---|---|---|---|---|---|---|
| Wilson 2019 | retrospective cohort | USA | 27,471 | warfarin (23109), apixaban/dabigatran, rivaroxaban (4362) | | | | CKD stage 1-2, stage 3, stage 4, stage 5 | | bleed, stroke, death, arterial/systemic embolism |
| Winkelmayer 2011 | retrospective cohort | USA | 237 | warfarin | 68.9 | 41.4 | heart failure: 148; CAD: 111; hypertension: 196 | patients on hemodialysis | | bleed, stroke, death |
| Yanagisawa 2018 | retrospective cohort | Japan | 1104 | warfarin (552), dabigatran/rivaroxaban/apixaban/edoxaban (552) | | | | CrCl 50-80 and 15-49 | | bleed |
| Yao 2017 | retrospective cohort | USA | 410 | apixaban (77), dabigatran (19), rivaroxaban (314) | 77.5 | | | apixaban: SCr level >1.5; dabigatran: eGFR<30; rivaroxaban: eGFR<50 | eGFR: 37.8 | bleed |
| Yao 2020 | retrospective cohort | USA | 32836 | warfarin (10680), apixaban (10880), dabigatran (3007), rivaroxaban (8269) | 72.8 (warfarin), 72.3 (apixaban), 67.2 (dabigatran), 69.4 (rivaroxaban) | 54.4 (warfarin), 54.9 (apixaban), 72.7 (dabigatran), 62.5 (rivaroxaban) | concurrent antiplatelet use: 1283 (warfarin), 1315 (apixaban), 367 (dabigatran), 1006 (rivaroxaban); heart failure: 4099 (warfarin), 4041 (apixaban), 1078 (dabigatran), 3067 (rivaroxaban); CAD: 6094 (warfarin), 6106 (apixaban), 1710 (dabigatran), 4620 (rivaroxaban); hypertenion: 9942 (warfarin), 10096 (apixaban), 2801 (dabigatran), 7701 (rivaroxaban); previous stroke: 1674 (warfarin), 1705 (apixaban), 460 (dabigatran), 1238 (rivaroxaban); previous VTE: 601 (warfarin), 575 (apixaban), 149 (dabigatran), 426 (rivaroxaban) | eGFR: 60-90, 45-59, 30-44, 15-39 | | bleed, stroke, death |
| Yodogawa 2016 | retrospective cohort | Japan | 30 | warfarin | 69.5 | 80 | concurrrent antiplatelet use: 12; heart failure: 6; hyerpertension: 17; previous stroke: 3 | end-stage renal disease requiring maintenance hemodialysis | | bleed, stroke, death |
| Yoon 2017 | retrospective cohort | | 2921 | warfarin | 67.8 | 59.9 | hypertension: 2612 | ESRD | | bleed, stroke |
| Yu 2018 | retrospective cohort | Korea | 8872 | warfarin (3016), edoxaban (5856) | 72.6 (warfarin), 68.2 (edoxaban) | 53.3 | heart failure: 2037 (warfarin), 3787 (edoxaban); hypertension: 2853 (warfarin), 5521 (edoxaban) | | | |

#### Studies in patients with valvular disease

##### Supplementary Table 5. Randomized controlled trials

| Author/Year | Study Design | Country | Sample Size | Intervention | Mean Age | % male | Comorbidities: N using concurrent antiplatelet, N heart failure, N CAD, N hypertension N previous stroke, N previous VTE | Included valve disease profile | Outcomes studied |
|---|---|---|---|---|---|---|---|---|---|
| Avezum 2015 | RCT | brazil | 2323 | warfarin (2323), apixaban (2405) | 71 | 60.7 | concurrent antiplatelet use: 1522, hypertension: 4102 | mitral stenosis, mitral regurgitation, aortic regurgitation, aortic stenosis | stroke or embolism |
| Breithardt 2016 | RCT | germany | 1001 | warfarin (1001), riveroxaban (939) | 78 | 60.8 | heart failure: 1376, hypertension: 1739 | Mitral regurgitation, mitral stenosis | bleed, stroke, death |
| DeCaterina 2017 | RCT | international | 955 | warfarin (955), edoxaban (1869) | 71.85 (warfarin), 72.27 (edoxaban) | 57.8 | CAD: 372 (warfarin), 750 (edoxaban); N hypertension: 896 (warfarin), 1733 (edoxaban), previous stroke: 234 (warfarin), 434 (edoxaban) | Mitral regurgitation, aortic regurgitation, aortic stenosis, valve surgery, bioprosthetic valves, valve repair | stroke, death |
| Duraes 2016 | RCT | Brazil | 12 | warfarin (12), dabigatran (15) | 45.7 (warfarin), 48.8 (dabigatran) | 37.0 | hypertension: 6 (warfarin), 7 (dabigatran); previous strokes: 4 (warfarin), 4 (dabigatran) | bioprosthetic valve (mitral/aortic valve replacement) | bleeding, stroke, TIA, arterial/systemic embolism, death |
| Ezekowitz 2016 | RCT | USA | 3950 | warfarin, dabigatran | 74 | 59.3 | heart failure: 1570, CAD: 1285, hypertension: 3051 | mitral stenosis, mitral regurgitation, aortic stenosis, aortic regurgitation, tricuspid regurgitation, | death |
| Guimaraes 2019 | RCT | USA | 69 | warfarin (69), apixaban (87) | 74 (warfarin), 92 (apixaban) | 60.9 | CAD: 32 (warfarin), 36 (apixaban); N hypertension: 64 (warfarin), 68 (apixaban) | bioprosthetic replacement (104), native valve repair (52) | bleeding, stroke, death |
| Lavitola 2010 | RCT | brazil | 119 | warfarin | | 20.2 | | biological mitral prosthesis | thromboembolism |

##### Supplementary Table 6. Non-randomized studies

| Author/Year | Study Design | Country | Sample Size | Intervention (n) | Mean Age | % male | Comorbidities: N using concurrent antiplatelet, N heart failure, N CAD, N hypertension N previous stroke, N previous VTE |
|---|---|---|---|---|---|---|---|
| Tanawuttiwat 2022 | retrospective cohort | USA | 21131 | VKA (13004), dabigatran (714), factor Xa inhibitor (7413) | 84 | 5.0 | hypertension: 11876 (VKA), 7492 (non-VKA); previous stroke: 1377 (VKA), 793 (non-VKA) |
| Briasoulis 2018 | retrospective cohort | USA | 20525 | Warfarin (16223), dabigatran (2132), rivaroxaban (2170) | 80 (warfarin), 77 (dabigatran, rivaroxaban) | 34.3 | heart failure: 9734 (warfarin), 1002 (dabigatran), 955 (rivaroxaban); hypertension: 14763 (warfarin), 1919 (dabigatran), 1931 (rivaroxaban) |

| | | | | | | | |
|---|---|---|---|---|---|---|---|
| Guimaraes 2020 | prospective cohort | Brazil | 1005 | warfarin (505), rivaroxaban (500) | 59.2 (warfarin), 59.4 (rivaroxaban) | 39.0 | heart failure: 188 (warfarin), 202 (rivaroxaban); hypertension: 302 (warfarin), 308 (rivaroxaban) |
| Izumi 2022 | prospective cohort | | 752 | Warfarin (489), DOAC (263) | 79.5 (warfarin), 82.3 (DOAC) | 44.7 | heart failure: 273 (warfarin), 145 (DOAC); CAD: 15 (warfarin), 12 (DOAC); hypertension: 353 (warfarin), 215 (DOAC) |
| Abdul-JawadAltisent 2016 | retrospective cohort | multicentre: Canada, france, spain, belgium, columbia | 621 | warfarin, warfarin + single antiplatelet, warfarin + dual antiplatelet | 81 | 46.5 | concurrent antiplatelet use: CAD: 343 |
| Strange 2021 | retrospective cohort | | 1562 | Rivaroxaban (620), apixaban (942) | | 47.5 | heart failure: 167 (rivaroxaban), 290 (apixaban); CAD: 28 (rivaroxaban), 52 (apixaban); hypertension: 448 (rivaroxaban), 692 (apixaban); previous stroke: 109 (rivaroxaban), 165 (apixaban); |
| Li 2021 | retrospective cohort | Taiwan | 11666 | warfarin (5833), DOACs (5833) | 75.68 (warfarin), 76.48 (DOACs) | 47.7 | concurrent antiplatelet use: 3897 (warfarin), 3901 (DOAC); heart failure: 2809 (warfarin), 2747 (DOAC); hypertension: 4118 (warfarin), 4090 (DOAC); previous stroke: 937 (warfarin), 935 (DOAC) |
| Mangner 2019 | prospective cohort | Germany | 182 | Rivaroxaban (111), apixaban (41), dabigatran (29). edoxaban (1) | 80 | 48.9 | CAD: 71; hypertension: 175; previous stroke: 26 |
| Seeger 2017 | prospective cohort | Germany | 272 | warfarin (131), apixaban (141) | 80.5 (warfarin), 82.1 (apixaban) | 50.7 | CAD: 77 (warfarin), 93 (apixaban); previous stroke: 19 (warfarin), 16 (apixaban) |
| Hampton 2020 | retrospective cohort | USA | 200 | Apixaban (133), rivaroxaban (50), dabigatran (17) | 77.6 apixaban, 74.73 rivaroxaban, 76.43 dabigatran | 51.0 | heart failure: 34; hypertension: 178; previous stroke: 36 |
| Izumi 2020 | retrospective cohort | Japan | 179 | warfarin (179), DOAC (16) | 76.7 (warfarin), 77.3 (DOAC) | 53.1 | concurrrent antiplatelet use: 79 (warfarin), 6 (DOAC); hypertension: 124 (warfarin), 14 (DOAC); previous stroke: 34 (warfarin), 2 (DOAC); previous VTE: 1 (warfarin), 0 (DOAC) |
| Russo 2018 | prospective cohort | Italy | 122 | DOAC (dabigatran, rivaroxaban, apixaban) | 74 | 55.7 | concurrent antiplatelet use: 17; heart failure: 54; CAD: 7; hypertension: 88; previous stroke: 7 |
| Wada 2001 | retrospective cohort | Japan | 53 | warfarin | 67.4 | 56.6 | concurrent antiplatelet use: 18; hypertension: 26 |
| Moon 2019 | retrospective cohort | korea | 5163 | warfarin (2371), DOAC (2792) | 71.2 | 57.1 | heart failure: 1223 (warfarin), 1423 (DOAC); hypertension: 1819 (warfarin), 2136 (DOAC) |
| Duan 2021 | retrospective cohort | USA | 2672 | warfarin (2233), apixaban/dabigatran/rivaroxaban (439) | | 60.6 | concurrent antiplatelet use: 654 (warfarin), 173 (DOAC); heart failure: 1697(warfarin), 308 (DOAC); hypertension: |

| | | | | | | | |
|---|---|---|---|---|---|---|---|
| | | | | | | | 1212 (warfarin), 250 (DOAC); previous stroke: 272 (warfarin), 57 (DOAC) |
| Luengsupabul 2020 | retrospective cohort | Thailand | 200 | warfarin | 64.8 | 62.5 | concurrent antiplatelet use: 36; heart failure: 81; CAD: 16; hypertension: 81 |
| Kosmidou 2019 | retrospective cohort | USA | 933 | warfarin (778), NOAC (155) | 82.8 | 65.6 | heart failure: 836; CAD: 188; hypertension: 856; previous stroke: 205 |
| DiBiase 2021 | prospective cohort | USA | 254 | warfarin (127), rivaroxaban (89), apixaban (38) | 63 | 66.1 | CAD: 20 (warfarin), 14 (DOAC); hypertension: 96 (warfarin), 99 (DOAC) |
| Acanfora 2016 | retrospective cohort | Italy | 27 | rivaroxaban | 70 | 66.7 | heart failure: 27, CAD: 8, hypertension: 20 |
| Kim 2019 | retrospective cohort | | 2230 | warfarin (1115), DOAC (1115) | 70.2 (warfarin), 69.2 (DOAC) | | heart failure: 838 (warfarin), 832 (DOAC); CAD: 267 (warfarin), 265 (DOAC); hypertension: 1080 (warfarin), 1076 (DOAC); previous stroke: 521 (warfarin), 508 (DOAC) |
| Mannacio 2022 | retrospective cohort | Italy | 692 | warfarin (692), dabigatran/rivaroxaban/apixaban/edoxaban (340) | | | heart failure: 167 (rivaroxaban), 290 (apixaban); CAD: 28 (rivaroxaban), 52 (apixaban); hypertension: 448 (rivaroxaban), 692 (apixaban); previous stroke: 167 (rivaroxaban), 290 (apixaban); |
| Melgaard 2021 | retrospective cohort | | 10043 | NOAC | 81 | | heart failure: 3112; CAD: 1676, hypertension: 4928 |
| Nauffal 2021 | retrospective cohort | USA | 16753 | warfarin (16753), NOAC (9769) | | | |

#### Subgroup Analyses

**Supplementary Figure 3**. Forest plot (A) and funnel plot (B) of association between number of GI bleeds and anticoagulation choice of DOAC vs warfarin in patients with concomitant atrial fibrillation and CKD.

(A)

| | DOAC | | Warfarin | | | Odds Ratio |
|---|---|---|---|---|---|---|
| Study or Subgroup | Events | Total | Events | Total | Weight | M-H, Random, 95% CI |
| **1.2.1 RCT** | | | | | | |
| Bohula 2016 | 232 | 7035 | 190 | 7036 | 14.1% | 1.23 [1.01, 1.49] |
| Fox 2011 | 82 | 1474 | 51 | 1476 | 12.6% | 1.65 [1.15, 2.35] |
| **Subtotal (95% CI)** | | **8509** | | **8512** | **26.8%** | **1.37 [1.04, 1.80]** |
| Total events | 314 | | 241 | | | |
| Heterogeneity: Tau² = 0.02; Chi² = 1.98, df = 1 (P = 0.16); I² = 50% | | | | | | |
| Test for overall effect: Z = 2.22 (P = 0.03) | | | | | | |
| **1.2.2 Non-RCT** | | | | | | |
| Bhatia 2019 | 6 | 152 | 0 | 0 | | Not estimable |
| Chan 2015 | 29 | 281 | 874 | 8064 | 12.3% | 0.95 [0.64, 1.40] |
| Chan 2019 | 13 | 5282 | 0 | 0 | | Not estimable |
| Coleman 2019 | 90 | 1896 | 307 | 4848 | 13.8% | 0.74 [0.58, 0.94] |
| DiLullo 2018 | 2 | 247 | 8 | 100 | 3.3% | 0.09 [0.02, 0.45] |
| Fu 2021 | 11 | 125 | 25 | 125 | 8.2% | 0.39 [0.18, 0.82] |
| Garg 2016 | 0 | 0 | 15 | 119 | | Not estimable |
| Kalil 2016 | 17 | 864 | 18 | 1710 | 9.2% | 1.89 [0.97, 3.68] |
| Kodani 2018 | 0 | 0 | 15 | 2686 | | Not estimable |
| Lin 2021 | 32 | 173 | 1010 | 3185 | 12.3% | 0.49 [0.33, 0.72] |
| Loo 2018 | 43 | 2596 | 0 | 0 | | Not estimable |
| Mitsuma 2015 | 0 | 0 | 6 | 27 | | Not estimable |
| Phan 2019 | 0 | 0 | 7 | 115 | | Not estimable |
| Shen 2015 | 0 | 0 | 153 | 1838 | | Not estimable |
| Siontis 2018 | 155 | 2351 | 710 | 7053 | 14.2% | 0.63 [0.53, 0.76] |
| Tan 2019 | 0 | 0 | 80 | 1651 | | Not estimable |
| Wang 2015 | 0 | 0 | 11 | 59 | | Not estimable |
| Winkelmayer 2011 | 0 | 0 | 48 | 237 | | Not estimable |
| Yoon 2017 | 0 | 0 | 215 | 2921 | | Not estimable |
| **Subtotal (95% CI)** | | **13967** | | **34738** | **73.2%** | **0.68 [0.50, 0.92]** |
| Total events | 398 | | 3502 | | | |
| Heterogeneity: Tau² = 0.10; Chi² = 23.86, df = 6 (P = 0.0006); I² = 75% | | | | | | |
| Test for overall effect: Z = 2.51 (P = 0.01) | | | | | | |
| **Total (95% CI)** | | **22476** | | **43250** | **100.0%** | **0.81 [0.59, 1.12]** |
| Total events | 712 | | 3743 | | | |
| Heterogeneity: Tau² = 0.19; Chi² = 64.13, df = 8 (P < 0.00001); I² = 88% | | | | | | |
| Test for overall effect: Z = 1.26 (P = 0.21) | | | | | | |
| Test for subgroup differences: Chi² = 11.25, df = 1 (P = 0.0008), I² = 91.1% | | | | | | |

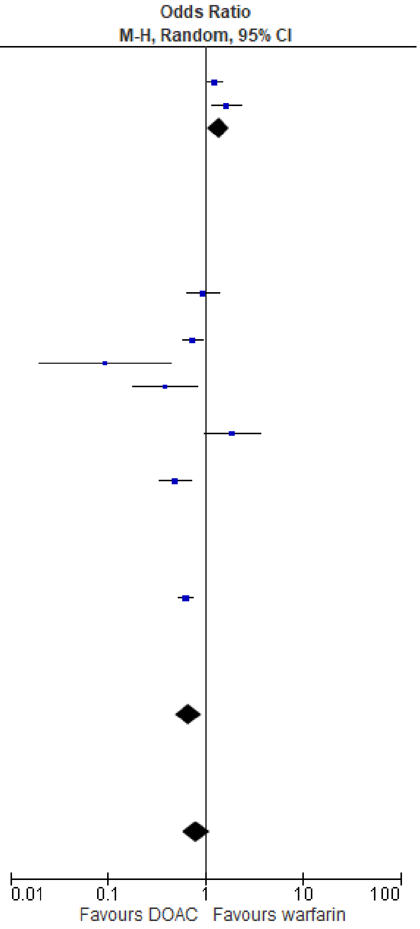

(B)

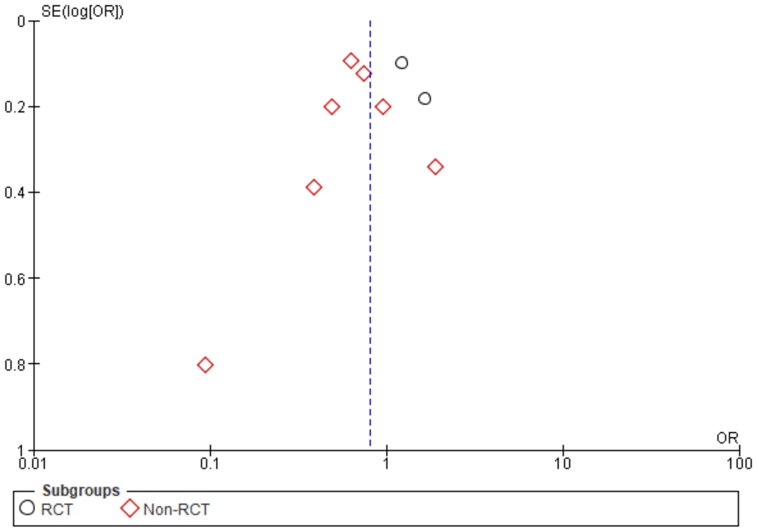

**Supplementary Figure 4**. Forest plot (A) and funnel plot (B) of association between number of intracranial bleeds and anticoagulation choice of DOAC vs warfarin in patients with concomitant atrial fibrillation and CKD.

(A)

| Study or Subgroup | DOAC Events | DOAC Total | Warfarin Events | Warfarin Total | Weight | Odds Ratio M-H, Random, 95% CI |
|---|---|---|---|---|---|---|
| **1.3.1 RCT** | | | | | | |
| Bohula 2016 | 61 | 7035 | 132 | 7036 | 21.1% | 0.46 [0.34, 0.62] |
| Fox 2011 | 20 | 1474 | 25 | 1476 | 15.0% | 0.80 [0.44, 1.44] |
| Hijazi 2014 | 150 | 6029 | 148 | 5957 | 22.6% | 1.00 [0.80, 1.26] |
| Stanifer 2017 | 0 | 135 | 4 | 132 | 1.5% | 0.11 [0.01, 1.98] |
| **Subtotal (95% CI)** | | **14673** | | **14601** | **60.2%** | **0.67 [0.38, 1.17]** |
| Total events | 231 | | 309 | | | |
| Heterogeneity: Tau² = 0.22; Chi² = 17.95, df = 3 (P = 0.0004); I² = 83% | | | | | | |
| Test for overall effect: Z = 1.40 (P = 0.16) | | | | | | |
| **1.3.2 Non-RCT** | | | | | | |
| Bhatia 2019 | 3 | 152 | 0 | 0 | | Not estimable |
| Bonnemeier 2019 | 38 | 6102 | 0 | 0 | | Not estimable |
| Chan 2019 | 12 | 5282 | 0 | 0 | | Not estimable |
| Chantrarat 2021a | 0 | 0 | 31 | 1659 | | Not estimable |
| Coleman 2019 | 2 | 90 | 19 | 307 | 4.9% | 0.34 [0.08, 1.51] |
| Fu 2021 | 2 | 125 | 4 | 125 | 3.9% | 0.49 [0.09, 2.74] |
| Garg 2016 | 0 | 0 | 19 | 119 | | Not estimable |
| Kodani 2018 | 0 | 0 | 39 | 4243 | | Not estimable |
| Lin 2021 | 7 | 173 | 236 | 3185 | 11.9% | 0.53 [0.24, 1.14] |
| Loo 2018 | 4 | 2596 | 0 | 0 | | Not estimable |
| Mitsuma 2015 | 0 | 0 | 0 | 27 | | Not estimable |
| Siontis 2018 | 21 | 2351 | 111 | 7053 | 17.6% | 0.56 [0.35, 0.90] |
| Stanifer 2020 | 0 | 136 | 4 | 133 | 1.5% | 0.11 [0.01, 1.98] |
| Wang 2015 | 0 | 0 | 4 | 59 | | Not estimable |
| **Subtotal (95% CI)** | | **17007** | | **16910** | **39.8%** | **0.52 [0.36, 0.76]** |
| Total events | 89 | | 467 | | | |
| Heterogeneity: Tau² = 0.00; Chi² = 1.56, df = 4 (P = 0.82); I² = 0% | | | | | | |
| Test for overall effect: Z = 3.43 (P = 0.0006) | | | | | | |
| **Total (95% CI)** | | **31680** | | **31511** | **100.0%** | **0.59 [0.41, 0.86]** |
| Total events | 320 | | 776 | | | |
| Heterogeneity: Tau² = 0.14; Chi² = 22.68, df = 8 (P = 0.004); I² = 65% | | | | | | |
| Test for overall effect: Z = 2.79 (P = 0.005) | | | | | | |
| Test for subgroup differences: Chi² = 0.53, df = 1 (P = 0.47), I² = 0% | | | | | | |

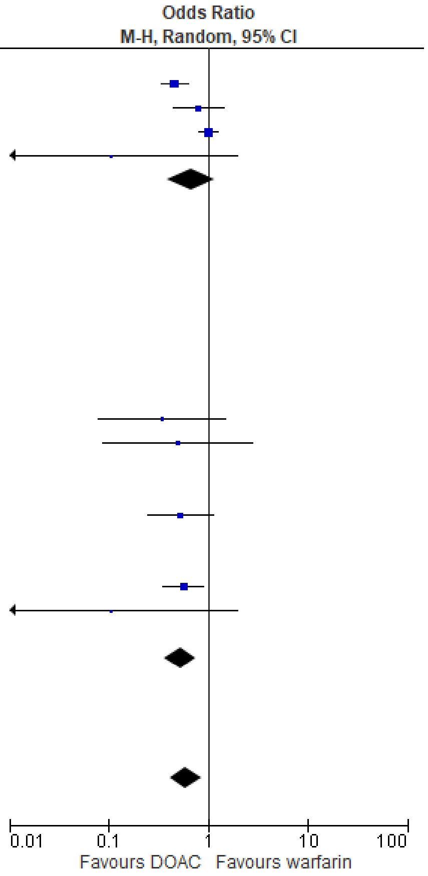

(B)

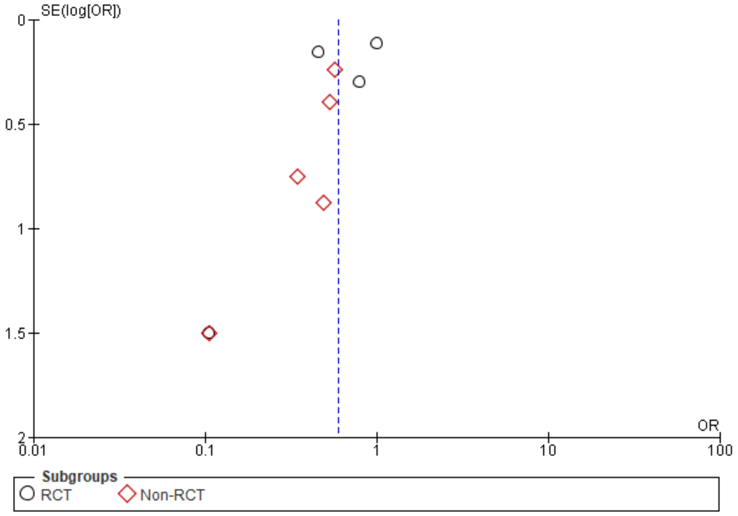

**Supplementary Figure 5**. Forest plot (A) and funnel plot (B) of association between number of fatal bleeds and anticoagulation choice of DOAC vs warfarin in patients with concomitant atrial fibrillation and CKD.

(A)

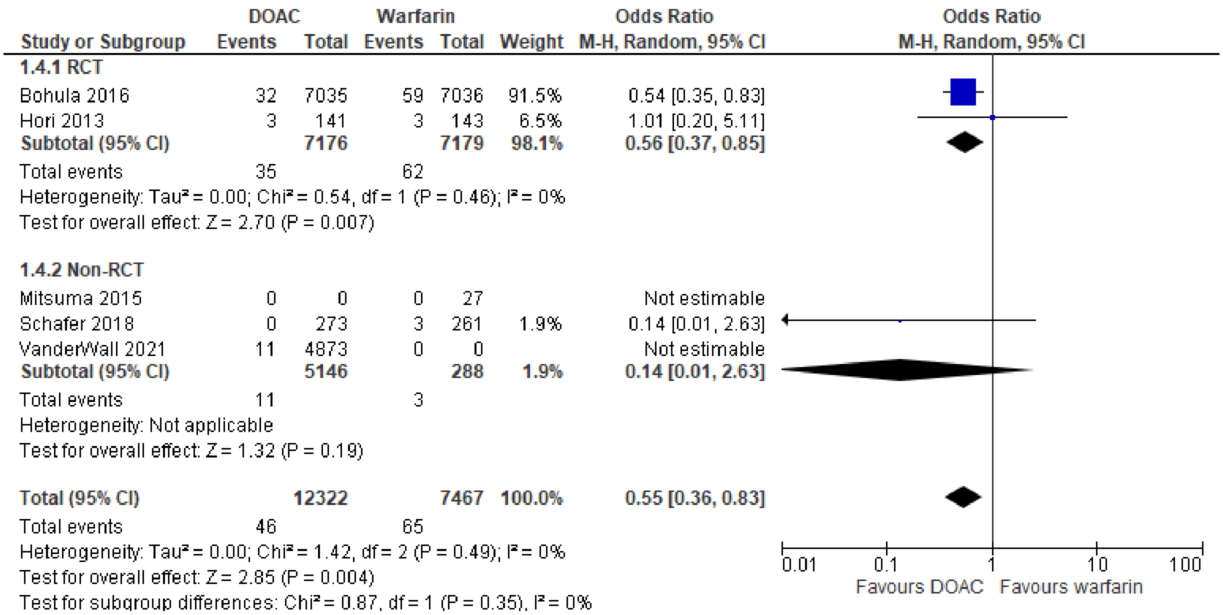

(B)

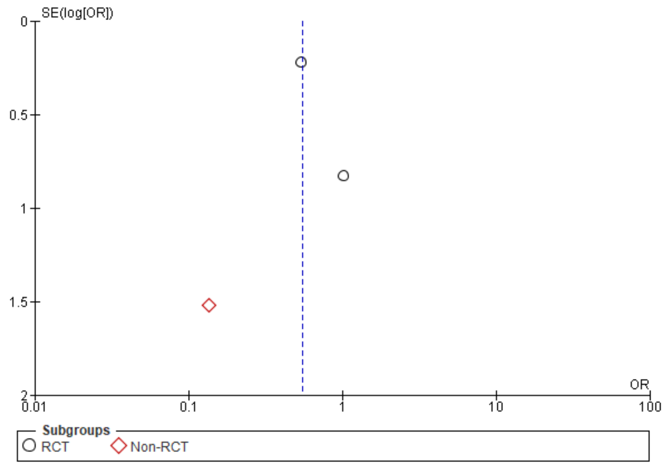

**Supplementary Figure 6**. Forest plot (A) and funnel plot (B) of association between incidence of TIAs and anticoagulation choice of DOAC vs warfarin in patients with concomitant atrial fibrillation and CKD.

(A)

| Study or Subgroup | DOAC Events | DOAC Total | Warfarin Events | Warfarin Total | Weight | Odds Ratio M-H, Random, 95% CI |
|---|---|---|---|---|---|---|
| **1.6.1 Non-RCT** | | | | | | |
| Chantrarat 2021b | 2 | 192 | 50 | 1741 | 100.0% | 0.36 [0.09, 1.47] |
| Chen 2014 | 0 | 0 | 15 | 294 | | Not estimable |
| Kodani 2018 | 0 | 0 | 8 | 4243 | | Not estimable |
| Matusik 2021 | 11 | 90 | 0 | 0 | | Not estimable |
| Mitsuma 2015 | 0 | 0 | 0 | 27 | | Not estimable |
| **Subtotal (95% CI)** | | **282** | | **6305** | **100.0%** | **0.36 [0.09, 1.47]** |
| Total events | 13 | | 73 | | | |
| Heterogeneity: Not applicable | | | | | | |
| Test for overall effect: Z = 1.42 (P = 0.15) | | | | | | |
| **Total (95% CI)** | | **282** | | **6305** | **100.0%** | **0.36 [0.09, 1.47]** |
| Total events | 13 | | 73 | | | |
| Heterogeneity: Not applicable | | | | | | |
| Test for overall effect: Z = 1.42 (P = 0.15) | | | | | | |
| Test for subgroup differences: Not applicable | | | | | | |

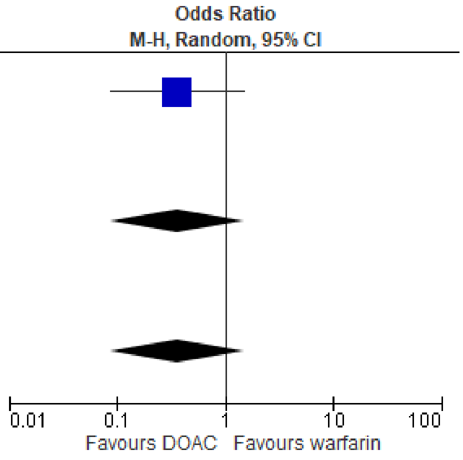

(B)

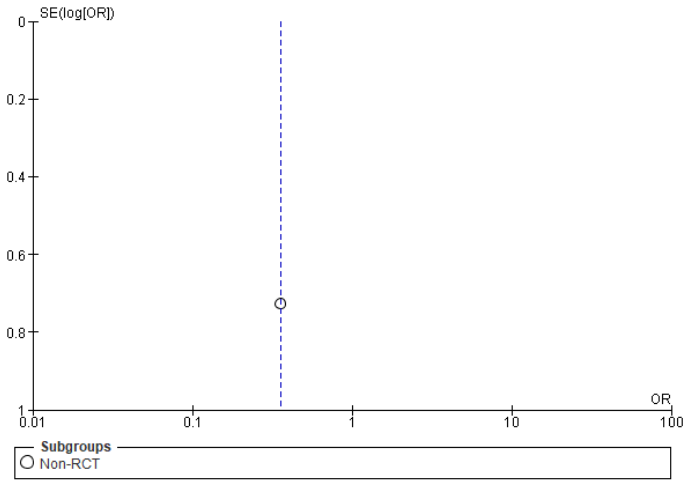

**Supplementary Figure 7**. Forest plot (A) and funnel plot (B) of association between incidence of systemic/arterial embolism and anticoagulation choice of DOAC vs warfarin in patients with concomitant atrial fibrillation and CKD.

(A)

| Study or Subgroup | DOAC Events | DOAC Total | Warfarin Events | Warfarin Total | Weight | Odds Ratio M-H, Random, 95% CI |
|---|---|---|---|---|---|---|
| **1.7.2 Non-RCT** | | | | | | |
| Chan 2015 | 2 | 281 | 25 | 8064 | 19.4% | 2.31 [0.54, 9.78] |
| Davis 2020 | 0 | 15 | 0 | 76 | | Not estimable |
| DiLullo 2018 | 0 | 247 | 5 | 100 | 11.3% | 0.04 [0.00, 0.64] |
| Fazio 2018 | 0 | 46 | 0 | 0 | | Not estimable |
| Fu 2021 | 0 | 125 | 6 | 125 | 11.3% | 0.07 [0.00, 1.31] |
| Genovesi 2016 | 0 | 0 | 11 | 134 | | Not estimable |
| Inoue 2018 | 0 | 0 | 63 | 2782 | | Not estimable |
| Kodani 2018 | 0 | 0 | 13 | 4243 | | Not estimable |
| Lin 2021 | 6 | 173 | 311 | 3185 | 23.2% | 0.33 [0.15, 0.76] |
| Mitsuma 2015 | 0 | 0 | 1 | 27 | | Not estimable |
| Schafer 2018 | 0 | 273 | 1 | 261 | 10.0% | 0.32 [0.01, 7.83] |
| Wang 2015 | 0 | 0 | 3 | 59 | | Not estimable |
| Wilson 2019 | 25 | 4362 | 67 | 23109 | 24.8% | 1.98 [1.25, 3.14] |
| **Subtotal (95% CI)** | | **5522** | | **42165** | **100.0%** | **0.49 [0.13, 1.81]** |
| Total events | 33 | | 506 | | | |
| Heterogeneity: Tau² = 1.73; Chi² = 28.18, df = 5 (P < 0.0001); I² = 82% | | | | | | |
| Test for overall effect: Z = 1.07 (P = 0.28) | | | | | | |
| **Total (95% CI)** | | **5522** | | **42165** | **100.0%** | **0.49 [0.13, 1.81]** |
| Total events | 33 | | 506 | | | |
| Heterogeneity: Tau² = 1.73; Chi² = 28.18, df = 5 (P < 0.0001); I² = 82% | | | | | | |
| Test for overall effect: Z = 1.07 (P = 0.28) | | | | | | |
| Test for subgroup differences: Not applicable | | | | | | |

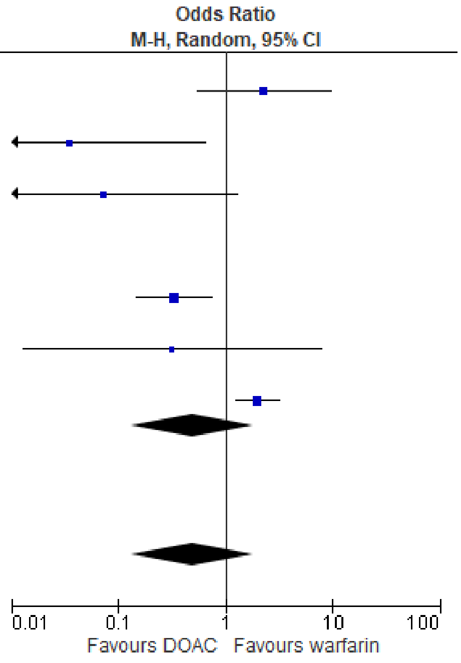

(B)

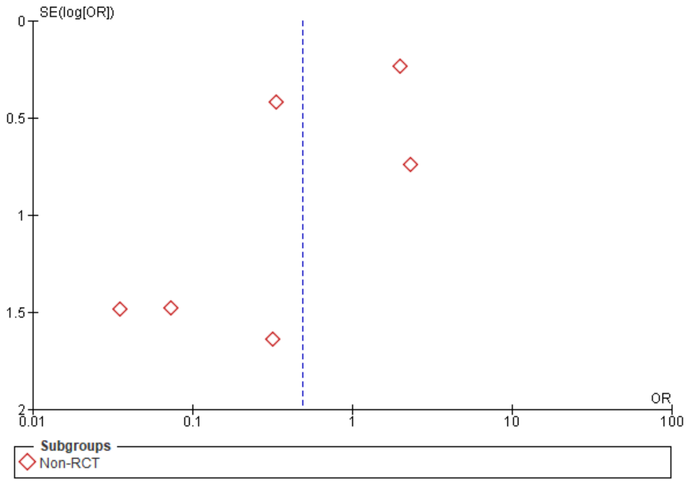

**Supplementary Figure 8**. Forest plot (A) and funnel plot (B) of association between all-cause mortality and anticoagulation choice of DOAC vs warfarin in patients with concomitant atrial fibrillation and CKD. (A)

| Study or Subgroup | DOAC Events | DOAC Total | Warfarin Events | Warfarin Total | Weight | Odds Ratio M-H, Random, 95% CI |
|---|---|---|---|---|---|---|
| **1.8.1 RCT** | | | | | | |
| Bohula 2016 | 773 | 7035 | 839 | 7036 | 11.2% | 0.91 [0.82, 1.01] |
| DeVriese 2020 | 15 | 46 | 19 | 44 | 1.3% | 0.64 [0.27, 1.50] |
| DeVriese 2021 | 30 | 46 | 32 | 44 | 1.2% | 0.70 [0.29, 1.73] |
| Eikelboom 2012 | 0 | 0 | 0 | 0 | | Not estimable |
| Fox 2011 | 0 | 0 | 0 | 0 | | Not estimable |
| Hart 2011 | 0 | 0 | 21 | 538 | | Not estimable |
| Hijazi 2014 | 401 | 6029 | 475 | 5965 | 10.3% | 0.82 [0.72, 0.95] |
| Hijazi 2021 | 0 | 0 | 0 | 0 | | Not estimable |
| Hohnloser 2012 | 244 | 9120 | 251 | 9081 | 9.2% | 0.97 [0.81, 1.16] |
| Hohnloser 2019 | 0 | 0 | 0 | 0 | | Not estimable |
| Hori 2013 | 0 | 0 | 0 | 0 | | Not estimable |
| Lip 2017 | 0 | 0 | 0 | 0 | | Not estimable |
| LipGYH 2017 | 0 | 0 | 0 | 0 | | Not estimable |
| Matsui 2022 | 69 | 706 | 0 | 0 | | Not estimable |
| Stanifer 2017 | 33 | 135 | 32 | 132 | 2.7% | 1.01 [0.58, 1.77] |
| **Subtotal (95% CI)** | | **23117** | | **22840** | **35.9%** | **0.89 [0.83, 0.96]** |
| Total events | 1565 | | 1669 | | | |
| Heterogeneity: Tau² = 0.00; Chi² = 3.30, df = 5 (P = 0.65); I² = 0% | | | | | | |
| Test for overall effect: Z = 3.02 (P = 0.003) | | | | | | |
| **1.8.2 Non-RCT** | | | | | | |
| Bhatia 2019 | 10 | 152 | 0 | 0 | | Not estimable |
| Carrero 2014 | 0 | 0 | 924 | 5292 | | Not estimable |
| Chan 2019 | 123 | 3162 | 0 | 0 | | Not estimable |
| Chantrarat 2021a | 0 | 0 | 126 | 1659 | | Not estimable |
| Elis 2021 | 19 | 97 | 45 | 155 | 2.3% | 0.60 [0.32, 1.10] |
| Fazio 2018 | 1 | 46 | 0 | 0 | | Not estimable |
| Fu 2021 | 11 | 125 | 13 | 125 | 1.3% | 0.83 [0.36, 1.93] |
| Garg 2016 | 0 | 0 | 97 | 119 | | Not estimable |
| Genovesi 2016 | 0 | 0 | 75 | 134 | | Not estimable |
| Gurevitz 2021 | 72 | 689 | 115 | 689 | 5.8% | 0.58 [0.42, 0.80] |
| Inoue 2018 | 0 | 0 | 100 | 2782 | | Not estimable |
| Kai 2017 | 0 | 0 | 433 | 888 | | Not estimable |
| Kodani 2018 | 0 | 0 | 19 | 864 | | Not estimable |
| Laugesen 2019 | 106 | 552 | 183 | 1008 | 6.9% | 1.07 [0.82, 1.40] |
| Lee 2015 | 2 | 59 | 15 | 174 | 0.5% | 0.37 [0.08, 1.68] |
| Lee 2017 | 0 | 0 | 340 | 589 | | Not estimable |
| Loo 2018 | 79 | 2596 | 0 | 0 | | Not estimable |
| Mitsuma 2015 | 0 | 0 | 8 | 27 | | Not estimable |
| Novikova 2021 | 13 | 79 | 0 | 0 | | Not estimable |
| Phan 2019 | 0 | 0 | 32 | 115 | | Not estimable |
| Siontis 2018 | 159 | 2351 | 753 | 7053 | 9.2% | 0.61 [0.51, 0.72] |
| Stanifer 2020 | 33 | 136 | 32 | 133 | 2.7% | 1.01 [0.58, 1.77] |
| Tan 2019 | 0 | 0 | 476 | 1651 | | Not estimable |
| VanderWall 2021 | 73 | 4873 | 0 | 0 | | Not estimable |
| Wakasugi 2014 | 0 | 0 | 9 | 28 | | Not estimable |
| Wang 2015 | 0 | 0 | 44 | 59 | | Not estimable |
| Wetmore 2020 | 1915 | 12210 | 1912 | 10529 | 12.0% | 0.84 [0.78, 0.90] |
| Wilson 2019 | 474 | 4362 | 3417 | 23109 | 11.3% | 0.70 [0.63, 0.78] |
| Winkelmayer 2011 | 0 | 0 | 181 | 237 | | Not estimable |
| Yao 2020 | 309 | 22156 | 210 | 10680 | 9.2% | 0.71 [0.59, 0.84] |
| Yodogawa 2016 | 0 | 0 | 6 | 30 | | Not estimable |
| Yu 2018 | 17 | 3016 | 58 | 3016 | 2.8% | 0.29 [0.17, 0.50] |
| **Subtotal (95% CI)** | | **56661** | | **71145** | **64.1%** | **0.71 [0.61, 0.81]** |
| Total events | 3416 | | 9623 | | | |
| Heterogeneity: Tau² = 0.03; Chi² = 40.63, df = 10 (P < 0.0001); I² = 75% | | | | | | |
| Test for overall effect: Z = 4.77 (P < 0.00001) | | | | | | |
| **Total (95% CI)** | | **79778** | | **93985** | **100.0%** | **0.77 [0.69, 0.85]** |
| Total events | 4981 | | 11292 | | | |
| Heterogeneity: Tau² = 0.02; Chi² = 55.47, df = 16 (P < 0.00001); I² = 71% | | | | | | |
| Test for overall effect: Z = 4.99 (P < 0.00001) | | | | | | |
| Test for subgroup differences: Chi² = 8.13, df = 1 (P = 0.004), I² = 87.7% | | | | | | |

Odds Ratio
M-H, Random, 95% CI

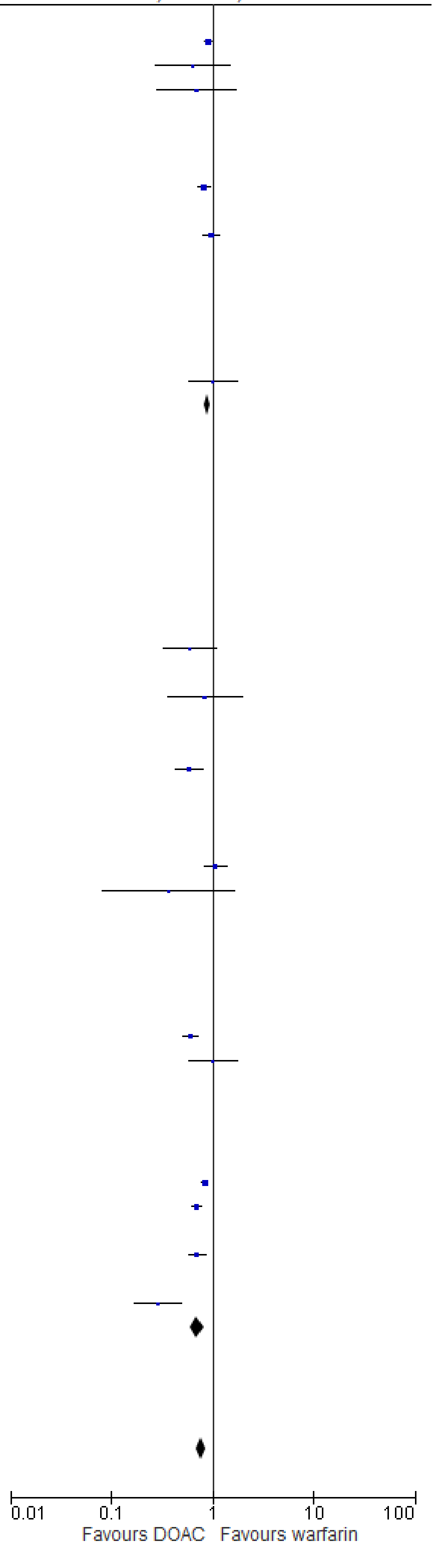

(B)

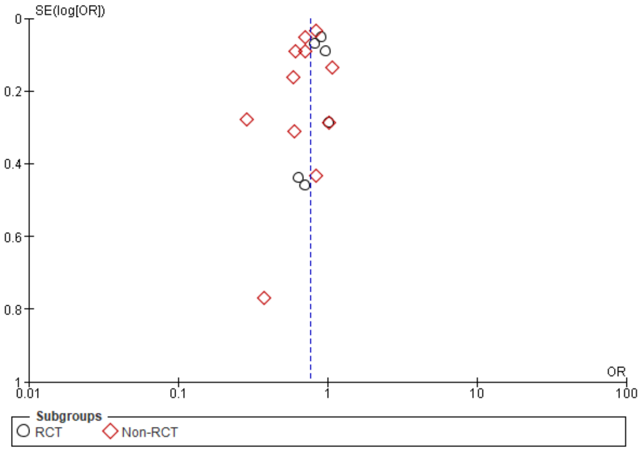

**Supplementary Figure 9**. Forest plot (A) and funnel plot (B) of association between number of bleeds and anticoagulation choice of apixaban versus warfarin in patients with concomitant atrial fibrillation and CKD.

(A)

| Study or Subgroup | Apixaban Events | Apixaban Total | Warfarin Events | Warfarin Total | Weight | Odds Ratio M-H, Random, 95% CI |
|---|---|---|---|---|---|---|
| **3.1.1 RCT** | | | | | | |
| Hijazi 2021 | 156 | 1131 | 207 | 1046 | 15.4% | 0.65 [0.52, 0.81] |
| Hohnloser 2012 | 230 | 9120 | 341 | 9081 | 18.6% | 0.66 [0.56, 0.79] |
| Stanifer 2017 | 7 | 136 | 19 | 133 | 2.3% | 0.33 [0.13, 0.80] |
| **Subtotal (95% CI)** | | **10387** | | **10260** | **36.3%** | **0.64 [0.55, 0.75]** |
| Total events | 393 | | 567 | | | |
| Heterogeneity: Tau² = 0.00; Chi² = 2.31, df = 2 (P = 0.32); I² = 13% | | | | | | |
| Test for overall effect: Z = 5.58 (P < 0.00001) | | | | | | |
| **3.1.2 Non-RCT** | | | | | | |
| Elis 2021 | 2 | 97 | 4 | 155 | 0.7% | 0.79 [0.14, 4.42] |
| Fu 2021 | 14 | 125 | 30 | 125 | 3.7% | 0.40 [0.20, 0.80] |
| Garza-Mayers 2018 | 5 | 17 | 0 | 0 | | Not estimable |
| Gurevitz 2021 | 11 | 689 | 12 | 689 | 2.7% | 0.92 [0.40, 2.09] |
| Jang 2020 | 55 | 245 | 0 | 0 | | Not estimable |
| Schafer 2018 | 4 | 273 | 22 | 261 | 1.7% | 0.16 [0.05, 0.48] |
| Siontis 2018 | 129 | 2351 | 715 | 7053 | 17.3% | 0.51 [0.42, 0.62] |
| Stanifer 2020 | 10 | 136 | 26 | 133 | 3.0% | 0.33 [0.15, 0.71] |
| Wetmore 2020 | 123 | 5874 | 270 | 8431 | 16.0% | 0.65 [0.52, 0.80] |
| Yao 2017 | 3 | 77 | 0 | 0 | | Not estimable |
| Yao 2020 | 205 | 10680 | 397 | 10680 | 18.6% | 0.51 [0.43, 0.60] |
| **Subtotal (95% CI)** | | **20564** | | **27527** | **63.7%** | **0.52 [0.44, 0.63]** |
| Total events | 561 | | 1476 | | | |
| Heterogeneity: Tau² = 0.02; Chi² = 12.30, df = 7 (P = 0.09); I² = 43% | | | | | | |
| Test for overall effect: Z = 6.99 (P < 0.00001) | | | | | | |
| **Total (95% CI)** | | **30951** | | **37787** | **100.0%** | **0.56 [0.48, 0.65]** |
| Total events | 954 | | 2043 | | | |
| Heterogeneity: Tau² = 0.02; Chi² = 19.62, df = 10 (P = 0.03); I² = 49% | | | | | | |
| Test for overall effect: Z = 7.93 (P < 0.00001) | | | | | | |
| Test for subgroup differences: Chi² = 2.96, df = 1 (P = 0.09), I² = 66.2% | | | | | | |

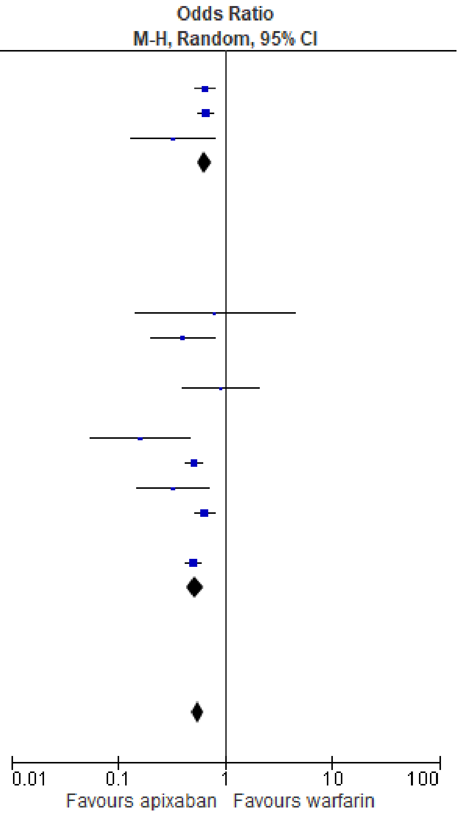

(B)

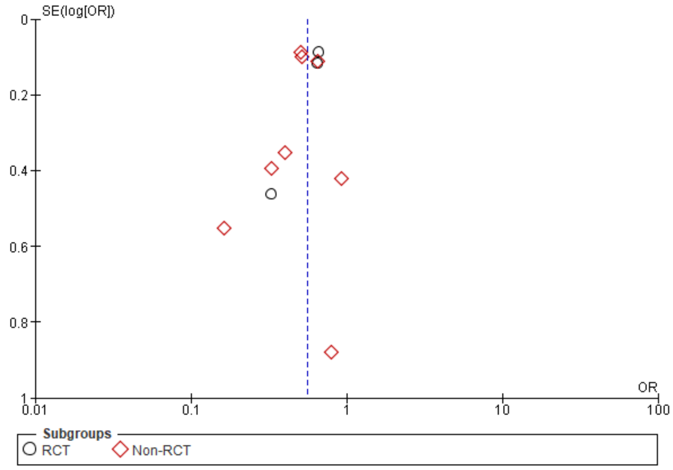

**Supplementary Figure 10**. Forest plot (A) and funnel plot (B) of association between incidence of stroke and anticoagulation choice of apixaban versus warfarin in patients with concomitant atrial fibrillation and CKD.

(A)

| Study or Subgroup | Apixaban Events | Apixaban Total | Warfarin Events | Warfarin Total | Weight | Odds Ratio M-H, Random, 95% CI |
|---|---|---|---|---|---|---|
| **3.2.1 RCT** | | | | | | |
| Hijazi 2021 | 136 | 1513 | 183 | 1422 | 38.7% | 0.67 [0.53, 0.85] |
| Hohnloser 2012 | 141 | 9120 | 185 | 9081 | 43.7% | 0.76 [0.61, 0.94] |
| Stanifer 2017 | 6 | 136 | 10 | 133 | 2.0% | 0.57 [0.20, 1.61] |
| **Subtotal (95% CI)** | | **10769** | | **10636** | **84.4%** | **0.71 [0.61, 0.83]** |
| Total events | 283 | | 378 | | | |
| Heterogeneity: Tau² = 0.00; Chi² = 0.73, df = 2 (P = 0.70); I² = 0% | | | | | | |
| Test for overall effect: Z = 4.23 (P < 0.0001) | | | | | | |
| **3.2.2 Non-RCT** | | | | | | |
| Fu 2021 | 6 | 125 | 10 | 125 | 2.0% | 0.58 [0.20, 1.65] |
| Schafer 2018 | 2 | 273 | 1 | 261 | 0.4% | 1.92 [0.17, 21.29] |
| Yao 2020 | 38 | 10680 | 65 | 10680 | 13.3% | 0.58 [0.39, 0.87] |
| **Subtotal (95% CI)** | | **11078** | | **11066** | **15.6%** | **0.60 [0.41, 0.87]** |
| Total events | 46 | | 76 | | | |
| Heterogeneity: Tau² = 0.00; Chi² = 0.92, df = 2 (P = 0.63); I² = 0% | | | | | | |
| Test for overall effect: Z = 2.71 (P = 0.007) | | | | | | |
| **Total (95% CI)** | | **21847** | | **21702** | **100.0%** | **0.69 [0.60, 0.80]** |
| Total events | 329 | | 454 | | | |
| Heterogeneity: Tau² = 0.00; Chi² = 2.32, df = 5 (P = 0.80); I² = 0% | | | | | | |
| Test for overall effect: Z = 4.96 (P < 0.00001) | | | | | | |
| Test for subgroup differences: Chi² = 0.67, df = 1 (P = 0.41), I² = 0% | | | | | | |

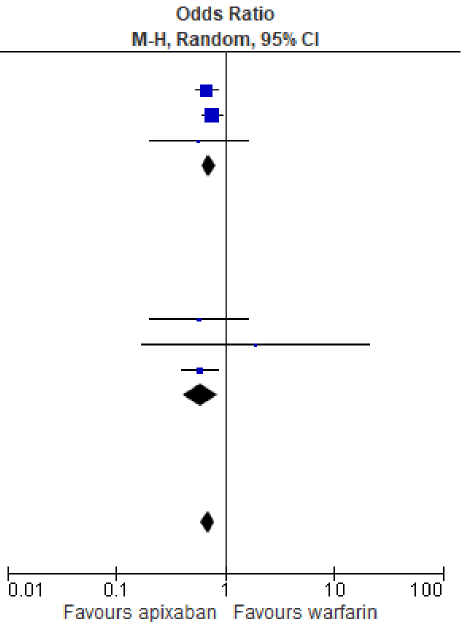

(B)

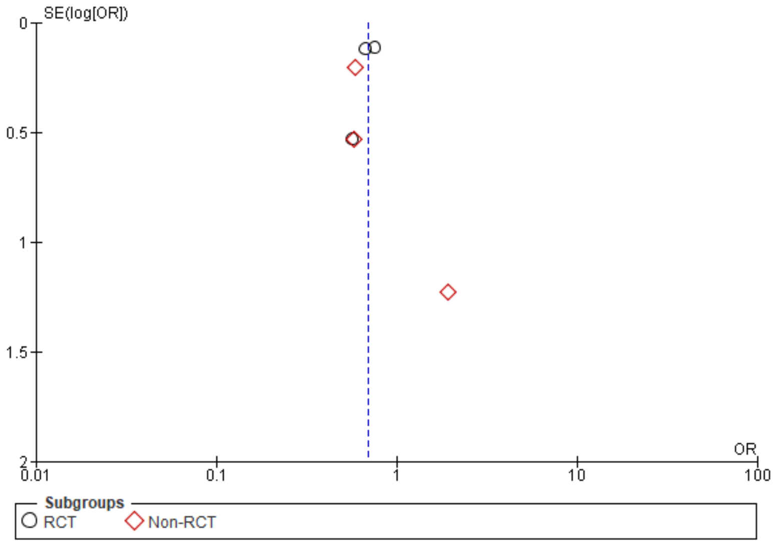

**Supplementary Figure 11**. Forest plot (A) and funnel plot (B) of association between number of bleeds and anticoagulation choice of dabigatran versus warfarin in patients with concomitant atrial fibrillation and CKD.

(A)

| Study or Subgroup | Dabigatran Events | Dabigatran Total | Warfarin Events | Warfarin Total | Weight | Odds Ratio M-H, Random, 95% CI |
|---|---|---|---|---|---|---|
| **4.1.1 Non-RCT** | | | | | | |
| Chan 2015 | 174 | 281 | 6031 | 8064 | 28.7% | 0.55 [0.43, 0.70] |
| Jang 2020 | 9 | 143 | 0 | 0 | | Not estimable |
| Kalil 2016 | 23 | 864 | 36 | 1710 | 18.1% | 1.27 [0.75, 2.16] |
| Loo 2018 | 23 | 2596 | 0 | 0 | | Not estimable |
| Novikova 2021 | 3 | 79 | 0 | 0 | | Not estimable |
| VanderWall 2021 | 38 | 4873 | 0 | 0 | | Not estimable |
| Wetmore 2020 | 40 | 1373 | 270 | 8431 | 25.1% | 0.91 [0.65, 1.27] |
| Yao 2017 | 2 | 19 | 0 | 0 | | Not estimable |
| Yao 2020 | 66 | 3007 | 397 | 10680 | 28.1% | 0.58 [0.45, 0.76] |
| **Subtotal (95% CI)** | | **13235** | | **28885** | **100.0%** | **0.74 [0.53, 1.02]** |
| Total events | 378 | | 6734 | | | |
| Heterogeneity: Tau² = 0.08; Chi² = 12.38, df = 3 (P = 0.006); I² = 76% | | | | | | |
| Test for overall effect: Z = 1.83 (P = 0.07) | | | | | | |
| **Total (95% CI)** | | **13235** | | **28885** | **100.0%** | **0.74 [0.53, 1.02]** |
| Total events | 378 | | 6734 | | | |
| Heterogeneity: Tau² = 0.08; Chi² = 12.38, df = 3 (P = 0.006); I² = 76% | | | | | | |
| Test for overall effect: Z = 1.83 (P = 0.07) | | | | | | |
| Test for subgroup differences: Not applicable | | | | | | |

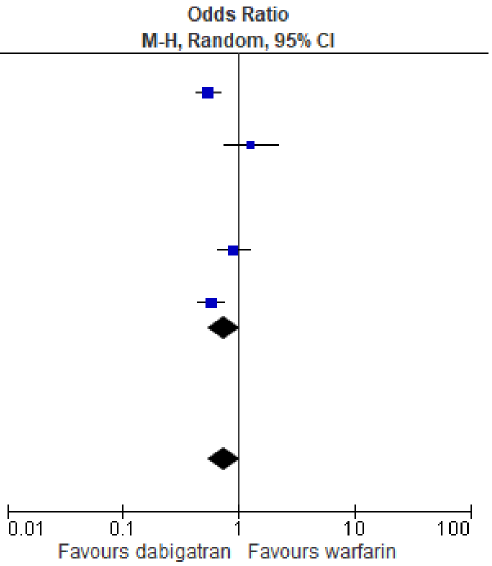

(B)

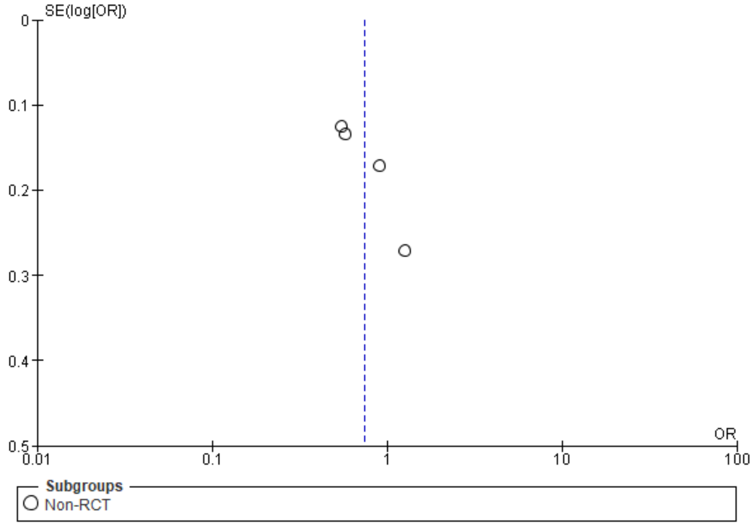

**Supplementary Figure 12**. Forest plot (A) and funnel plot (B) of association between stroke incidence and anticoagulation choice of dabigatran versus warfarin in patients with concomitant atrial fibrillation and CKD.

(A)

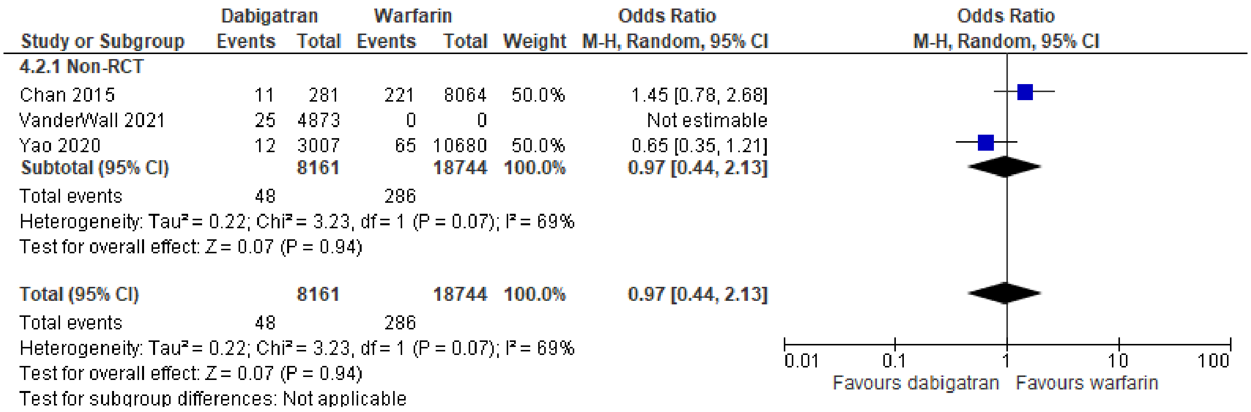

(B)

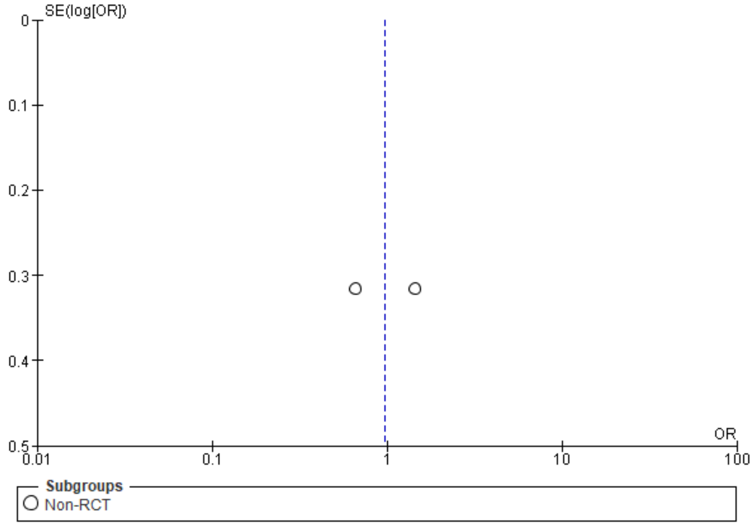

**Supplementary Figure 13**. Forest plot (A) and funnel plot (B) of association between number of bleeds and anticoagulation choice of edoxaban versus warfarin in patients with concomitant atrial fibrillation and CKD.

(A)

| | Edoxaban | | Warfarin | | | Odds Ratio |
|---|---|---|---|---|---|---|
| **Study or Subgroup** | **Events** | **Total** | **Events** | **Total** | **Weight** | **M-H, Random, 95% CI** |
| **5.1.1 RCT** | | | | | | |
| Bohula 2016 | 418 | 7035 | 524 | 7036 | 59.4% | 0.79 [0.69, 0.90] |
| Lip 2017 | 17 | 1067 | 13 | 1082 | 11.1% | 1.33 [0.64, 2.75] |
| LipGYH 2017 | 17 | 1095 | 13 | 1104 | 11.1% | 1.32 [0.64, 2.74] |
| **Subtotal (95% CI)** | | **9197** | | **9222** | **81.6%** | **0.97 [0.65, 1.44]** |
| Total events | 452 | | 550 | | | |
| Heterogeneity: Tau² = 0.06; Chi² = 3.76, df = 2 (P = 0.15); I² = 47% | | | | | | |
| Test for overall effect: Z = 0.15 (P = 0.88) | | | | | | |
| **5.1.2 Non-RCT** | | | | | | |
| Fazio 2018 | 0 | 46 | 0 | 0 | | Not estimable |
| Yu 2018 | 23 | 3016 | 35 | 3016 | 18.4% | 0.65 [0.39, 1.11] |
| **Subtotal (95% CI)** | | **3062** | | **3016** | **18.4%** | **0.65 [0.39, 1.11]** |
| Total events | 23 | | 35 | | | |
| Heterogeneity: Not applicable | | | | | | |
| Test for overall effect: Z = 1.57 (P = 0.12) | | | | | | |
| **Total (95% CI)** | | **12259** | | **12238** | **100.0%** | **0.85 [0.66, 1.11]** |
| Total events | 475 | | 585 | | | |
| Heterogeneity: Tau² = 0.03; Chi² = 4.36, df = 3 (P = 0.23); I² = 31% | | | | | | |
| Test for overall effect: Z = 1.18 (P = 0.24) | | | | | | |
| Test for subgroup differences: Chi² = 1.37, df = 1 (P = 0.24), I² = 27.0% | | | | | | |

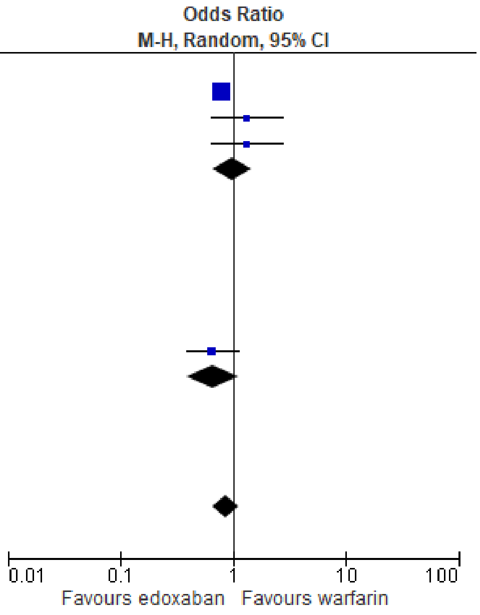

(B)

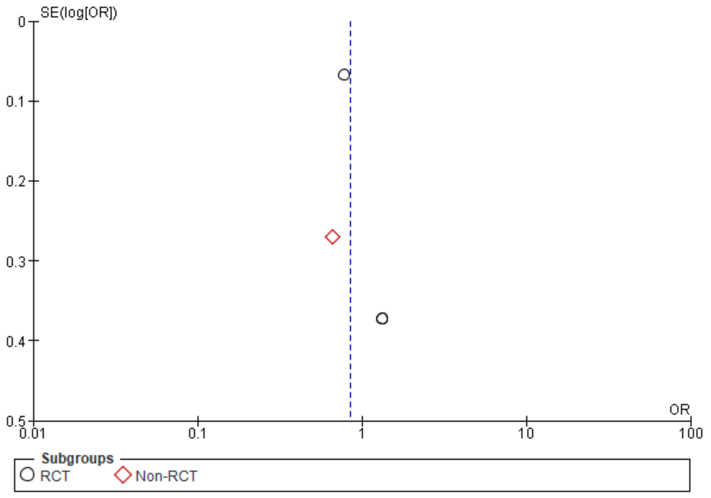

**Supplementary Figure 14**. Forest plot (A) and funnel plot (B) of association between number of bleeds and anticoagulation choice of rivaroxaban versus warfarin in patients with concomitant atrial fibrillation and CKD.

(A)

| | Rivaroxaban | | Warfarin | | | Odds Ratio |
|---|---|---|---|---|---|---|
| **Study or Subgroup** | **Events** | **Total** | **Events** | **Total** | **Weight** | **M-H, Fixed, 95% CI** |
| **6.1.1 RCT** | | | | | | |
| DeVriese 2020 | 0 | 0 | 0 | 0 | | Not estimable |
| DeVriese 2021 | 0 | 0 | 0 | 0 | | Not estimable |
| Fox 2011 | 128 | 1474 | 135 | 1476 | 12.5% | 0.94 [0.73, 1.22] |
| Hori 2013 | 17 | 141 | 20 | 143 | 1.8% | 0.84 [0.42, 1.69] |
| Matsui 2022 | 131 | 706 | 0 | 0 | | Not estimable |
| **Subtotal (95% CI)** | | **2321** | | **1619** | **14.2%** | **0.93 [0.73, 1.18]** |
| Total events | 276 | | 155 | | | |
| Heterogeneity: Chi² = 0.09, df = 1 (P = 0.76); I² = 0% | | | | | | |
| Test for overall effect: Z = 0.58 (P = 0.56) | | | | | | |
| **6.1.2 Non-RCT** | | | | | | |
| Chan 2019 | 26 | 3162 | 0 | 0 | | Not estimable |
| Coleman 2019 | 99 | 1896 | 418 | 4848 | 22.5% | 0.58 [0.47, 0.73] |
| Jang 2020 | 35 | 204 | 0 | 0 | | Not estimable |
| Lin 2021 | 23 | 173 | 560 | 3185 | 5.1% | 0.72 [0.46, 1.13] |
| Weir 2017 | 54 | 1290 | 132 | 2468 | 8.8% | 0.77 [0.56, 1.07] |
| Weir 2020 | 66 | 2317 | 144 | 0 | | Not estimable |
| Wetmore 2020 | 111 | 3488 | 270 | 8431 | 15.5% | 0.99 [0.79, 1.24] |
| Yao 2017 | 19 | 314 | 0 | 0 | | Not estimable |
| Yao 2020 | 259 | 8269 | 397 | 10680 | 33.9% | 0.84 [0.71, 0.98] |
| **Subtotal (95% CI)** | | **21113** | | **29612** | **85.8%** | **0.79 [0.71, 0.87]** |
| Total events | 692 | | 1921 | | | |
| Heterogeneity: Chi² = 11.61, df = 4 (P = 0.02); I² = 66% | | | | | | |
| Test for overall effect: Z = 4.59 (P < 0.00001) | | | | | | |
| **Total (95% CI)** | | **23434** | | **31231** | **100.0%** | **0.81 [0.73, 0.89]** |
| Total events | 968 | | 2076 | | | |
| Heterogeneity: Chi² = 13.22, df = 6 (P = 0.04); I² = 55% | | | | | | |
| Test for overall effect: Z = 4.47 (P < 0.00001) | | | | | | |
| Test for subgroup differences: Chi² = 1.67, df = 1 (P = 0.20), I² = 40.2% | | | | | | |

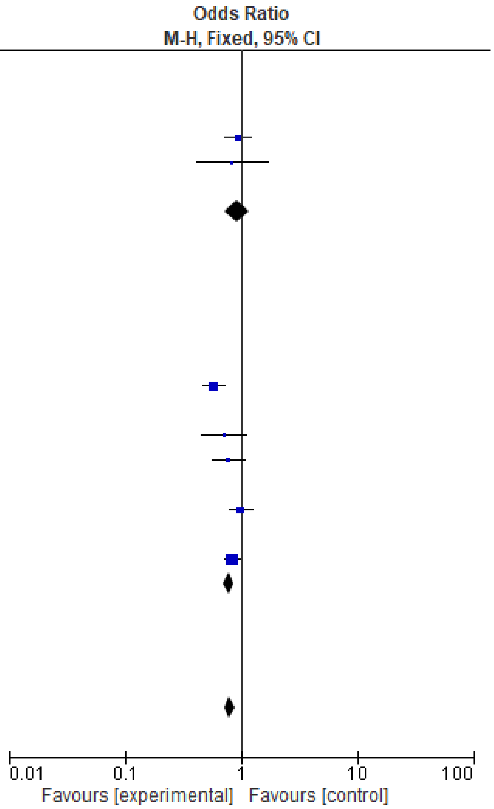

(B)

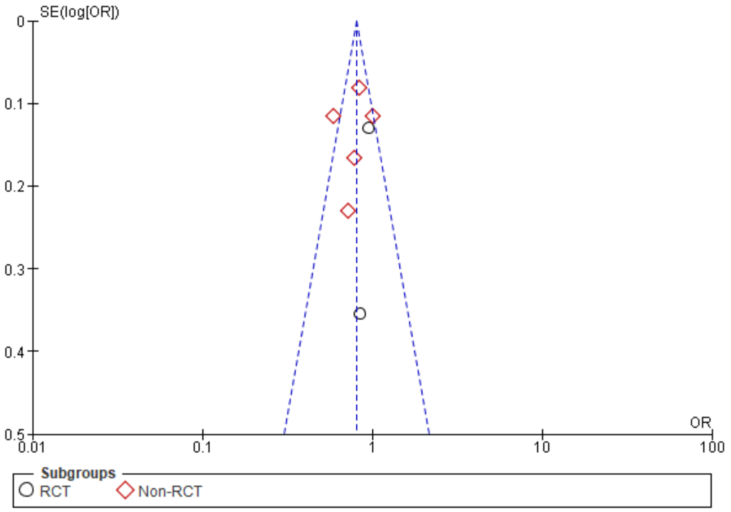

**Supplementary Figure 15**. Forest plot (A) and funnel plot (B) of association between stroke incidence and anticoagulation choice of rivaroxaban versus warfarin in patients with concomitant atrial fibrillation and CKD.

(A)

| Study or Subgroup | Rivaroxaban Events | Rivaroxaban Total | Warfarin Events | Warfarin Total | Weight | Odds Ratio M-H, Random, 95% CI |
|---|---|---|---|---|---|---|
| **6.2.1 RCT** | | | | | | |
| DeVriese 2020 | 2 | 46 | 5 | 44 | 2.9% | 0.35 [0.07, 1.93] |
| DeVriese 2021 | 4 | 46 | 7 | 44 | 4.5% | 0.50 [0.14, 1.86] |
| Fox 2011 | 66 | 1474 | 65 | 1476 | 15.4% | 1.02 [0.72, 1.44] |
| Hori 2013 | 9 | 141 | 9 | 143 | 7.0% | 1.02 [0.39, 2.64] |
| Matsui 2022 | 27 | 706 | 0 | 0 | | Not estimable |
| **Subtotal (95% CI)** | | **2413** | | **1707** | **29.7%** | **0.94 [0.69, 1.29]** |
| Total events | 108 | | 86 | | | |
| Heterogeneity: Tau² = 0.00; Chi² = 2.37, df = 3 (P = 0.50); I² = 0% | | | | | | |
| Test for overall effect: Z = 0.37 (P = 0.71) | | | | | | |
| **6.2.2 Non-RCT** | | | | | | |
| Bonnemeier 2019 | 156 | 6102 | 0 | 0 | | Not estimable |
| Coleman 2019 | 23 | 1896 | 98 | 4848 | 13.6% | 0.60 [0.38, 0.94] |
| DiLullo 2018 | 0 | 247 | 25 | 100 | 1.2% | 0.01 [0.00, 0.10] |
| Lin 2021 | 7 | 173 | 236 | 3185 | 9.0% | 0.53 [0.24, 1.14] |
| Weir 2017 | 54 | 1290 | 132 | 2468 | 15.8% | 0.77 [0.56, 1.07] |
| Weir 2020 | 66 | 2317 | 144 | 2317 | 16.2% | 0.44 [0.33, 0.60] |
| Yao 2020 | 39 | 8269 | 65 | 10680 | 14.6% | 0.77 [0.52, 1.15] |
| **Subtotal (95% CI)** | | **20294** | | **23598** | **70.3%** | **0.56 [0.38, 0.83]** |
| Total events | 345 | | 700 | | | |
| Heterogeneity: Tau² = 0.15; Chi² = 19.27, df = 5 (P = 0.002); I² = 74% | | | | | | |
| Test for overall effect: Z = 2.90 (P = 0.004) | | | | | | |
| **Total (95% CI)** | | **22707** | | **25305** | **100.0%** | **0.63 [0.46, 0.87]** |
| Total events | 453 | | 786 | | | |
| Heterogeneity: Tau² = 0.14; Chi² = 28.04, df = 9 (P = 0.0009); I² = 68% | | | | | | |
| Test for overall effect: Z = 2.82 (P = 0.005) | | | | | | |
| Test for subgroup differences: Chi² = 4.06, df = 1 (P = 0.04), I² = 75.4% | | | | | | |

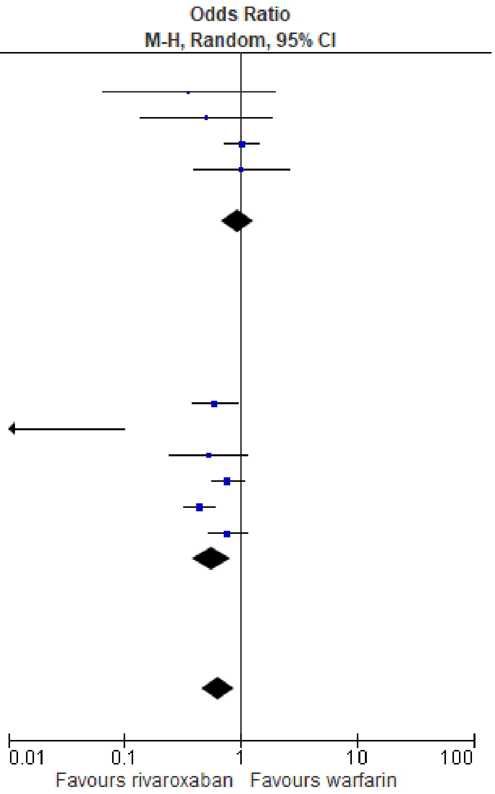

(B)

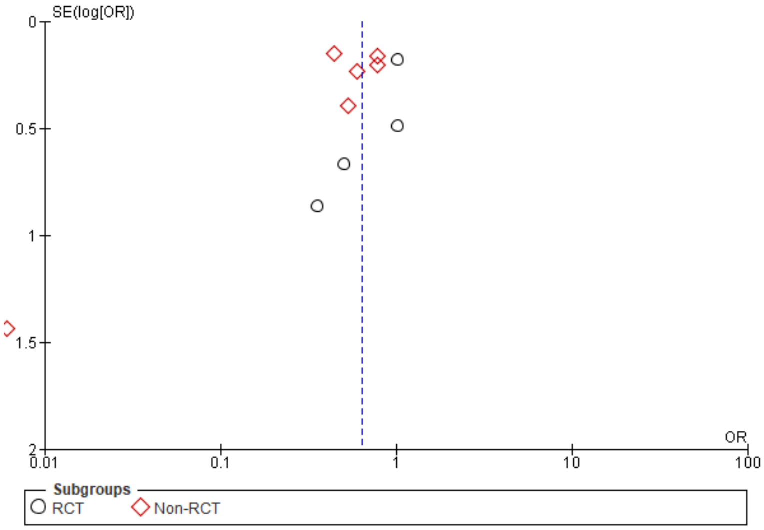

**Supplementary Figure 16**. Forest plot (A) and funnel plot (B) of association between stroke incidence and anticoagulation choice of DOAC versus warfarin in patients with concomitant atrial fibrillation and CKD of stages 1-2, stage 3, stage 4-5, or on dialysis.

(A)

| Study or Subgroup | DOAC Events | DOAC Total | Warfarin Events | Warfarin Total | Weight | Odds Ratio M-H, Random, 95% CI |
|---|---|---|---|---|---|---|
| **2.2.1 Stage 1-2** | | | | | | |
| Carrero 2014 | 0 | 0 | 65 | 2584 | | Not estimable |
| Shin 2018 | 191 | 2083 | 193 | 2084 | 15.3% | 0.99 [0.80, 1.22] |
| Yao 2020 | 24 | 11708 | 16 | 5601 | 8.3% | 0.72 [0.38, 1.35] |
| **Subtotal (95% CI)** | | **13791** | | **10269** | **23.6%** | **0.96 [0.78, 1.17]** |
| Total events | 215 | | 274 | | | |
| Heterogeneity: Tau² = 0.00; Chi² = 0.89, df = 1 (P = 0.34); I² = 0% | | | | | | |
| Test for overall effect: Z = 0.42 (P = 0.67) | | | | | | |
| **2.2.2 Stage 3** | | | | | | |
| Bhatia 2019 | 3 | 112 | 0 | 0 | | Not estimable |
| Carrero 2014 | 0 | 0 | 76 | 2270 | | Not estimable |
| Lee 2015 | 3 | 174 | 10 | 59 | 3.0% | 0.09 [0.02, 0.32] |
| Shin 2018 | 114 | 1023 | 108 | 1125 | 14.2% | 1.18 [0.89, 1.56] |
| Yao 2020 | 47 | 4343 | 22 | 2051 | 10.1% | 1.01 [0.61, 1.68] |
| **Subtotal (95% CI)** | | **5652** | | **5505** | **27.3%** | **0.63 [0.26, 1.52]** |
| Total events | 167 | | 216 | | | |
| Heterogeneity: Tau² = 0.47; Chi² = 14.37, df = 2 (P = 0.0008); I² = 86% | | | | | | |
| Test for overall effect: Z = 1.02 (P = 0.31) | | | | | | |
| **2.2.3 Stage 4-5** | | | | | | |
| Bhatia 2019 | 6 | 40 | 0 | 0 | | Not estimable |
| Bonde 2016 | 0 | 0 | 89 | 219 | | Not estimable |
| Bonde 2017 | 0 | 0 | 118 | 372 | | Not estimable |
| Carrero 2014 | 0 | 0 | 25 | 372 | | Not estimable |
| Chang 2019 | 52 | 475 | 25 | 273 | 10.2% | 1.22 [0.74, 2.01] |
| Elis 2021 | 2 | 97 | 4 | 155 | 1.9% | 0.79 [0.14, 4.42] |
| Fazio 2018 | 0 | 46 | 0 | 0 | | Not estimable |
| Fu 2021 | 14 | 125 | 30 | 125 | 7.5% | 0.40 [0.20, 0.80] |
| Jang 2020 | 8 | 31 | 0 | 0 | | Not estimable |
| Jun 2015 | 0 | 0 | 113 | 641 | | Not estimable |
| Matusik 2021 | 0 | 0 | 14 | 90 | | Not estimable |
| Mitsuma 2015 | 0 | 0 | 0 | 0 | | Not estimable |
| Yao 2020 | 137 | 2751 | 93 | 1405 | 14.3% | 0.74 [0.56, 0.97] |
| **Subtotal (95% CI)** | | **3565** | | **3652** | **34.0%** | **0.75 [0.49, 1.17]** |
| Total events | 219 | | 511 | | | |
| Heterogeneity: Tau² = 0.10; Chi² = 6.77, df = 3 (P = 0.08); I² = 56% | | | | | | |
| Test for overall effect: Z = 1.27 (P = 0.20) | | | | | | |
| **2.2.4 Dialysis** | | | | | | |
| Chan 2009 | 0 | 0 | 74 | 747 | | Not estimable |
| Chan 2015 | 11 | 281 | 221 | 8064 | 8.5% | 1.45 [0.78, 2.68] |
| Chan 2016 | 0 | 0 | 0 | 67 | | Not estimable |
| Davis 2020 | 0 | 15 | 0 | 76 | | Not estimable |
| Garg 2016 | 0 | 0 | 13 | 119 | | Not estimable |
| Kai 2017 | 0 | 0 | 78 | 888 | | Not estimable |
| Lee 2017 | 0 | 0 | 56 | 589 | | Not estimable |
| Lin 2021 | 7 | 173 | 236 | 3185 | 6.7% | 0.53 [0.24, 1.14] |
| Mitsuma 2015 | 0 | 0 | 2 | 27 | | Not estimable |
| Phan 2019 | 0 | 0 | 12 | 115 | | Not estimable |
| Shah 2014 | 0 | 0 | 52 | 756 | | Not estimable |
| Shen 2015 | 0 | 0 | 92 | 1838 | | Not estimable |
| Tan 2019 | 0 | 0 | 124 | 1651 | | Not estimable |
| Wakasugi 2014 | 0 | 0 | 8 | 28 | | Not estimable |
| Wang 2015 | 0 | 0 | 5 | 59 | | Not estimable |
| Winkelmayer 2011 | 0 | 0 | 38 | 237 | | Not estimable |
| Yodogawa 2016 | 0 | 0 | 2 | 30 | | Not estimable |
| **Subtotal (95% CI)** | | **469** | | **18476** | **15.1%** | **0.90 [0.33, 2.46]** |
| Total events | 18 | | 1013 | | | |
| Heterogeneity: Tau² = 0.41; Chi² = 4.21, df = 1 (P = 0.04); I² = 76% | | | | | | |
| Test for overall effect: Z = 0.21 (P = 0.83) | | | | | | |
| **Total (95% CI)** | | **23477** | | **37902** | **100.0%** | **0.83 [0.64, 1.07]** |
| Total events | 619 | | 2014 | | | |
| Heterogeneity: Tau² = 0.10; Chi² = 29.95, df = 10 (P = 0.0009); I² = 67% | | | | | | |
| Test for overall effect: Z = 1.44 (P = 0.15) | | | | | | |
| Test for subgroup differences: Chi² = 1.63, df = 3 (P = 0.65), I² = 0% | | | | | | |

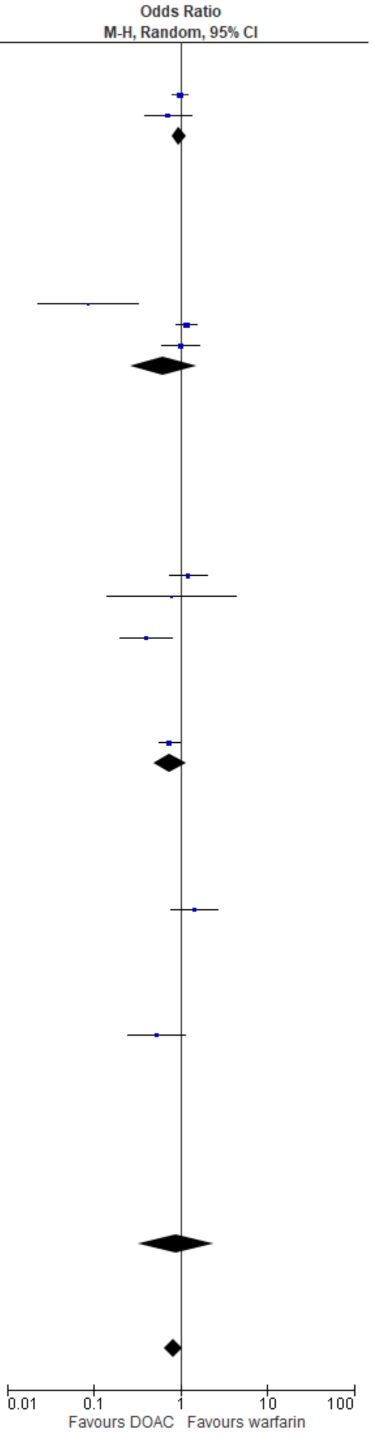

(B)

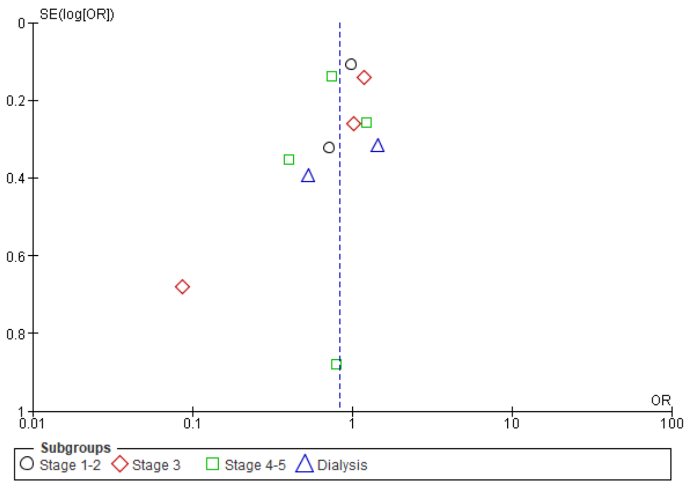
SE(log[OR])
0
0.2
0.4
0.6
0.8
1
0.01
0.1
1
10
100
OR
Subgroups
Stage 1-2
Stage 3
Stage 4-5
Dialysis

**Supplementary Figure 17**. Forest plot (A) and funnel plot (B) of association between number of bleeds with anticoagulation choice of apixaban versus warfarin in patients with concomitant atrial fibrillation and valve disease.

(A)

| | Rivaroxaban | | Warfarin | | | Odds Ratio |
|---|---|---|---|---|---|---|
| **Study or Subgroup** | **Events** | **Total** | **Events** | **Total** | **Weight** | **M-H, Random, 95% CI** |
| Briasoulis 2018 | 24 | 1270 | 264 | 16223 | 57.6% | 1.16 [0.76, 1.78] |
| DiBiase 2021 | 0 | 38 | 0 | 127 | | Not estimable |
| Guimaraes 2020 | 0 | 500 | 12 | 505 | 42.4% | 0.04 [0.00, 0.67] |
| **Total (95% CI)** | | **1808** | | **16855** | **100.0%** | **0.28 [0.01, 9.56]** |
| Total events | 24 | | 276 | | | |

Heterogeneity: $Tau^2 = 5.61$; $Chi^2 = 6.27$, df = 1 (P = 0.01); $I^2 = 84\%$

Test for overall effect: Z = 0.71 (P = 0.48)

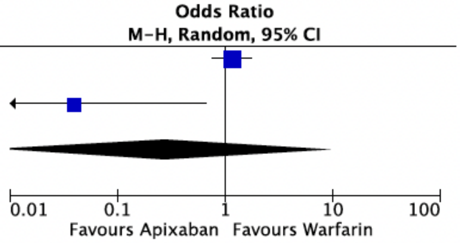

(B)

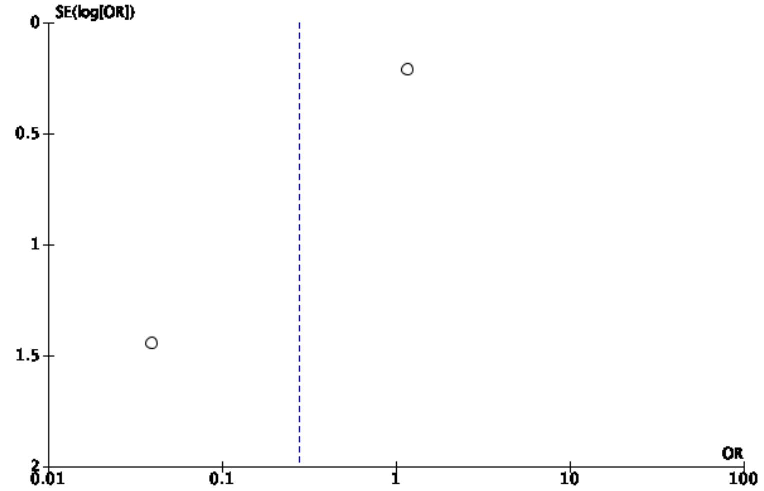

**Supplementary Figure 18**. Forest plot (A) and funnel plot (B) of association between incidences of arterial/systemic embolism with anticoagulation choice of apixaban versus warfarin in patients with concomitant atrial fibrillation and valve disease.

(A)

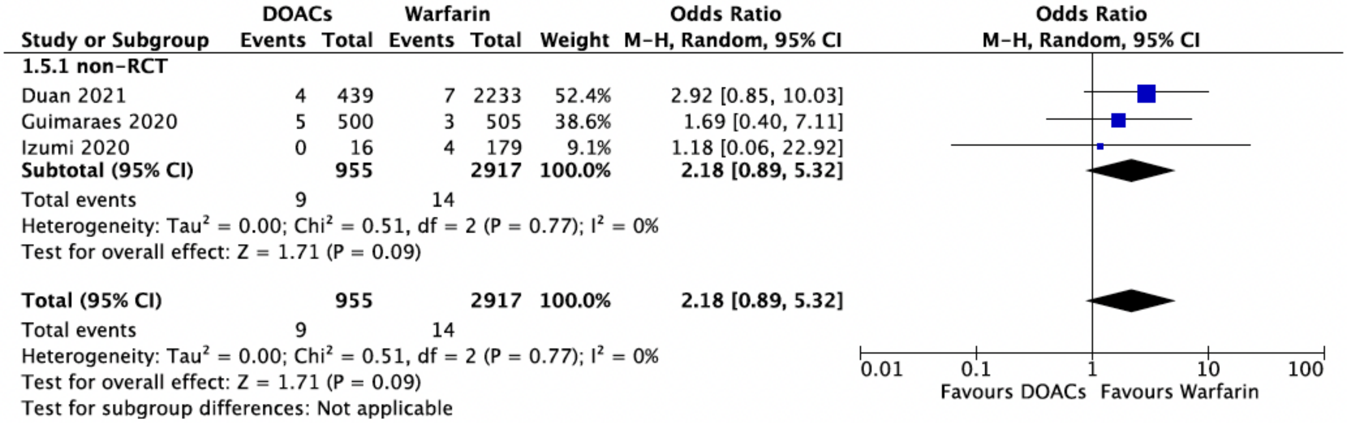

(B)

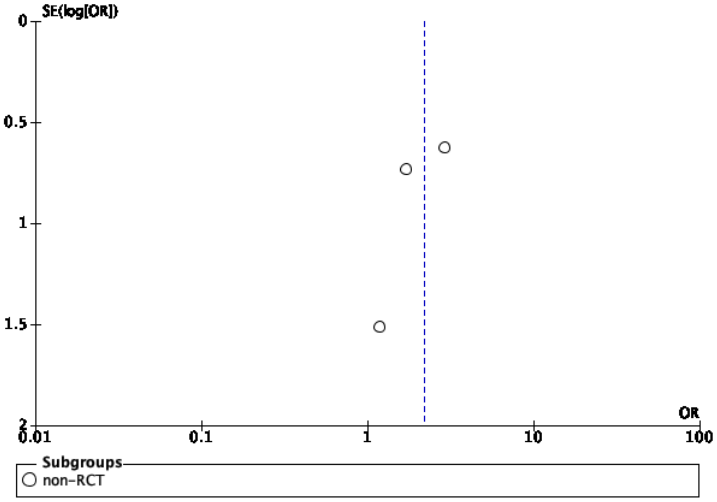

**Supplementary Figure 19**. Forest plot (A) and funnel plot (B) of association between incidences of all-cause mortality with anticoagulation choice of apixaban versus warfarin in patients with concomitant atrial fibrillation and valve disease.

(A)

| Study or Subgroup | DOACs Events | DOACs Total | Warfarin Events | Warfarin Total | Weight | Odds Ratio M-H, Random, 95% CI |
|---|---|---|---|---|---|---|
| **1.4.1 RCT** | | | | | | |
| Breithardt 2016 | 95 | 939 | 112 | 1001 | 10.1% | 0.89 [0.67, 1.19] |
| DeCaterina 2017 | 308 | 1869 | 147 | 955 | 10.7% | 1.08 [0.88, 1.34] |
| Duraes 2016 | 0 | 15 | 1 | 12 | 0.7% | 0.25 [0.01, 6.64] |
| Guimaraes 2019 | 7 | 87 | 6 | 69 | 3.8% | 0.92 [0.29, 2.87] |
| **Subtotal (95% CI)** | | **2910** | | **2037** | **25.3%** | **1.01 [0.85, 1.19]** |
| Total events | 410 | | 266 | | | |
| Heterogeneity: $Tau^2 = 0.00$; $Chi^2 = 1.84$, df = 3 (P = 0.61); $I^2 = 0\%$ | | | | | | |
| Test for overall effect: Z = 0.08 (P = 0.94) | | | | | | |
| **1.4.2 Non-RCT** | | | | | | |
| Briasoulis 2018 | 130 | 4302 | 1023 | 16223 | 10.9% | 0.46 [0.38, 0.56] |
| Duan 2021 | 39 | 439 | 259 | 2233 | 9.6% | 0.74 [0.52, 1.06] |
| Guimaraes 2020 | 20 | 500 | 20 | 505 | 7.1% | 1.01 [0.54, 1.90] |
| Izumi 2020 | 0 | 16 | 25 | 179 | 0.9% | 0.18 [0.01, 3.16] |
| Kosmidou 2019 | 33 | 155 | 207 | 778 | 9.0% | 0.75 [0.49, 1.13] |
| Moon 2019 | 161 | 2792 | 352 | 2371 | 10.8% | 0.35 [0.29, 0.43] |
| Nauffal 2021 | 96 | 9769 | 128 | 16753 | 10.3% | 1.29 [0.99, 1.68] |
| Seeger 2017 | 19 | 141 | 6 | 131 | 4.8% | 3.24 [1.25, 8.40] |
| Tanawuttiwat 2022 | 1284 | 8127 | 2367 | 13004 | 11.3% | 0.84 [0.78, 0.91] |
| **Subtotal (95% CI)** | | **26241** | | **52177** | **74.7%** | **0.77 [0.54, 1.09]** |
| Total events | 1782 | | 4387 | | | |
| Heterogeneity: $Tau^2 = 0.21$; $Chi^2 = 120.56$, df = 8 (P < 0.00001); $I^2 = 93\%$ | | | | | | |
| Test for overall effect: Z = 1.48 (P = 0.14) | | | | | | |
| **Total (95% CI)** | | **29151** | | **54214** | **100.0%** | **0.80 [0.61, 1.06]** |
| Total events | 2192 | | 4653 | | | |
| Heterogeneity: $Tau^2 = 0.17$; $Chi^2 = 133.42$, df = 12 (P < 0.00001); $I^2 = 91\%$ | | | | | | |
| Test for overall effect: Z = 1.56 (P = 0.12) | | | | | | |
| Test for subgroup differences: $Chi^2 = 1.85$, df = 1 (P = 0.17), $I^2 = 45.9\%$ | | | | | | |

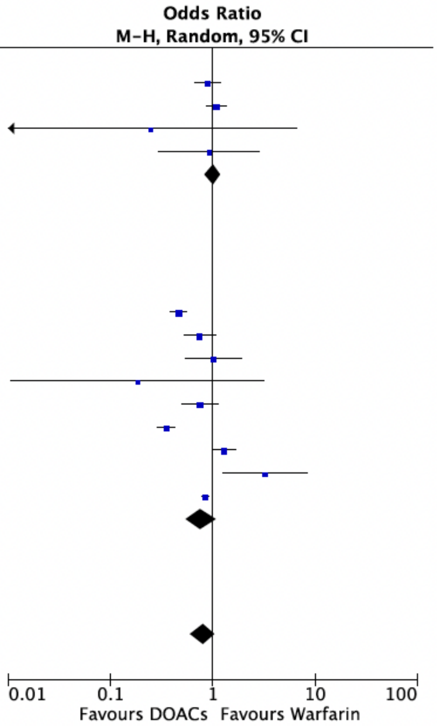

(B)

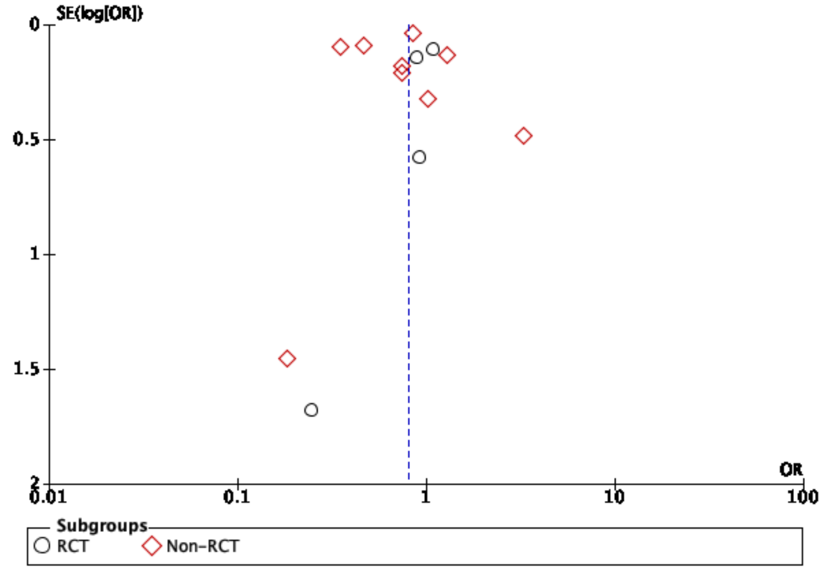

**Funnel plots for the main analyses**

**Supplementary Figure 20**. Funnel plot for the association between number of overall bleeds and anticoagulation choice of DOAC vs warfarin in patients with concomitant atrial fibrillation and CKD.

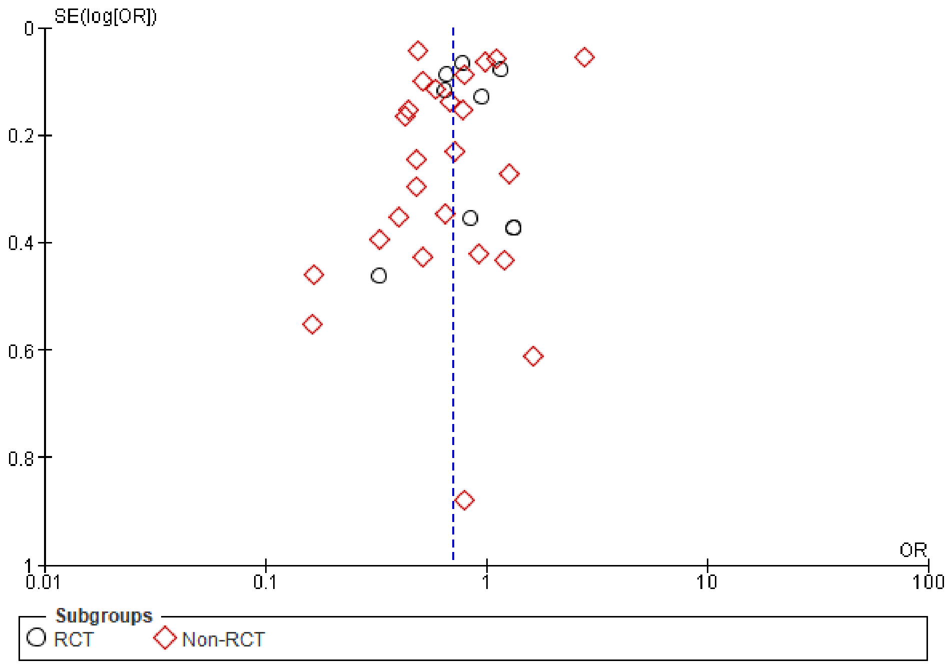

**Supplementary Figure 21**. Funnel plot for the association between incidence of strokes and anticoagulation choice of DOAC vs warfarin in patients with concomitant atrial fibrillation and CKD.

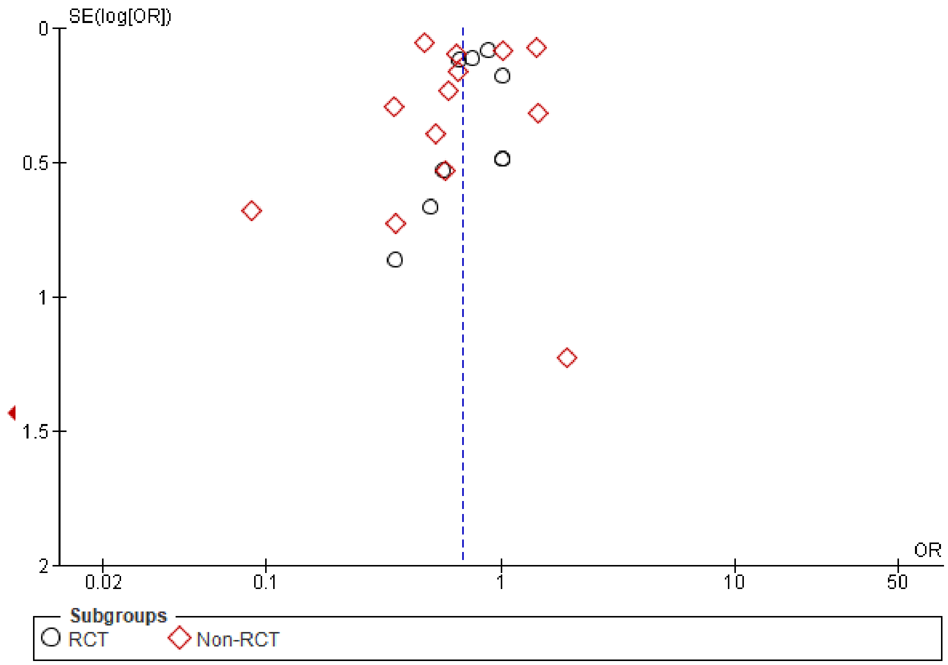

**Supplementary Figure 22**. Funnel plot for the association between number of bleeds and anticoagulation choice of DOAC versus warfarin in patients with concomitant atrial fibrillation and CKD of stages 1-2, stage 3, stage 4-5, or on dialysis.

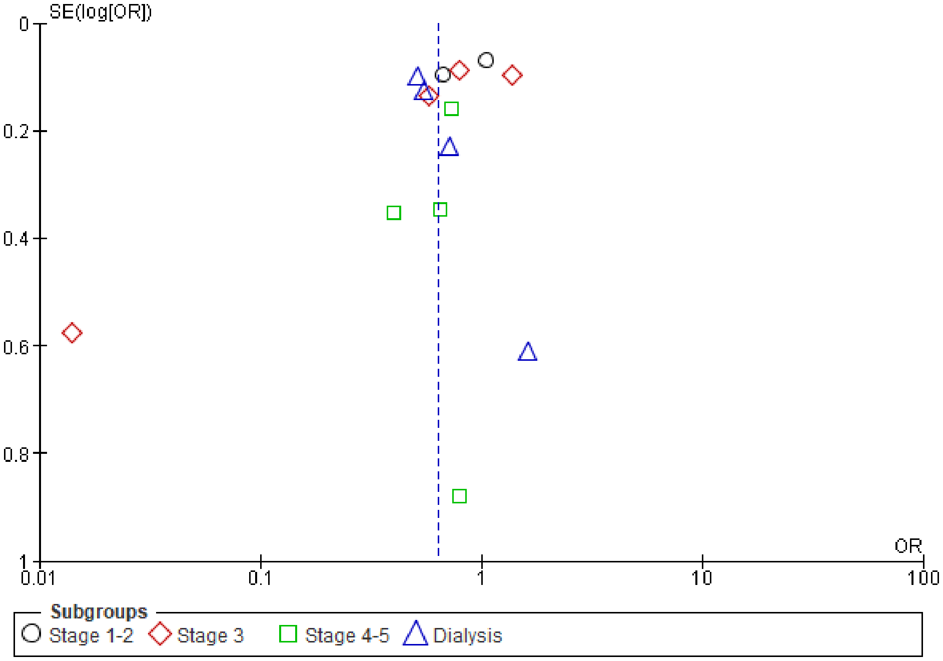

**Supplementary Figure 23**. Funnel plots for the bleeding outcomes with warfarin vs DOAC in patients with concomitant atrial fibrillation and valve disease.

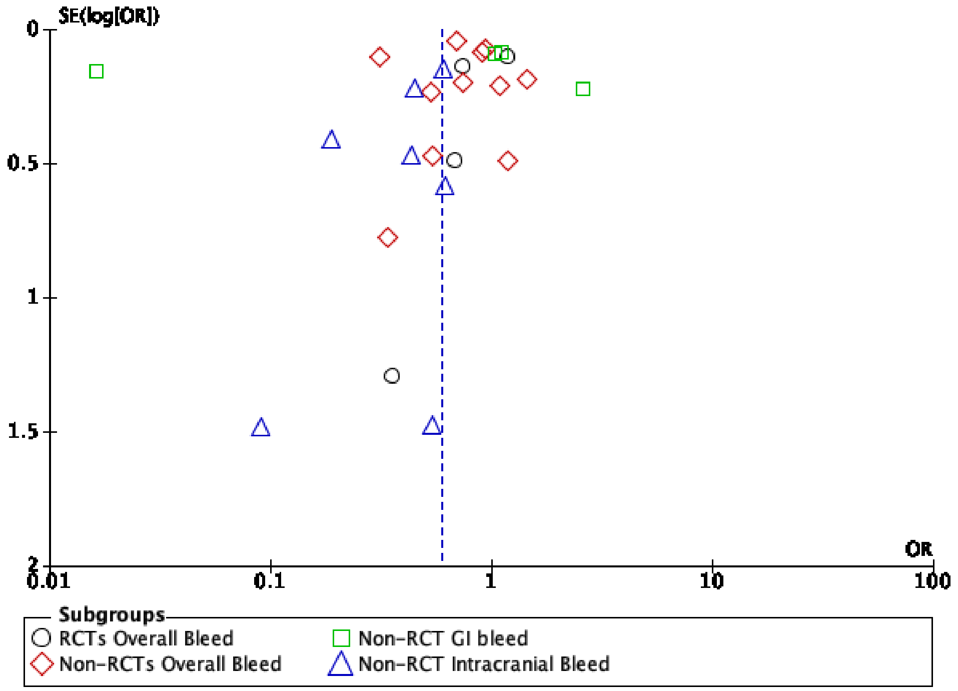

**Supplementary Figure 24**. Funnel plot for the association between stroke and TIA incidences with anticoagulation choice of DOAC versus warfarin in patients with concomitant atrial fibrillation and valve disease.

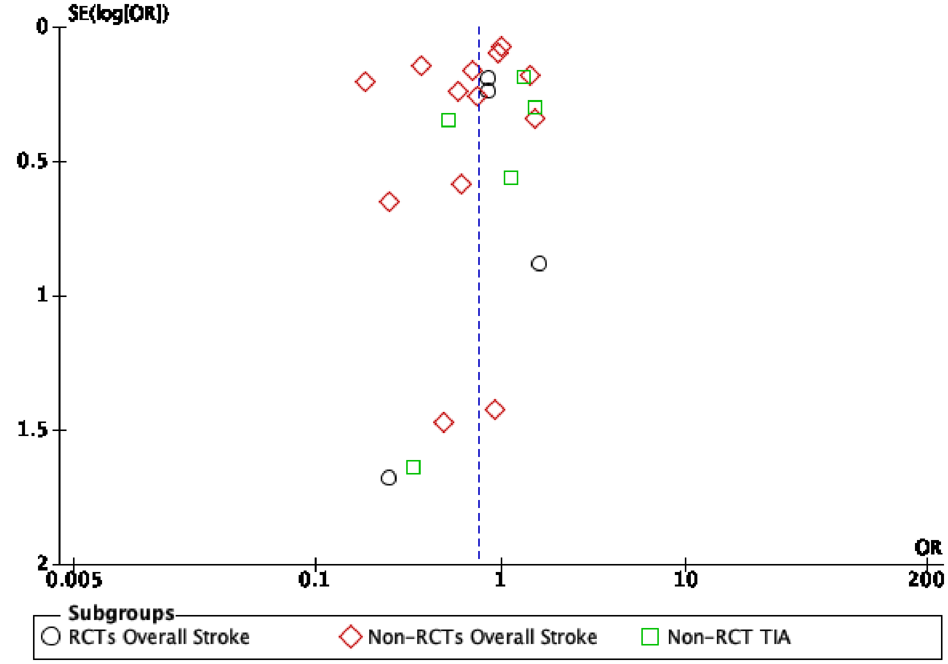
